# Identification of a type 1 diabetes-associated T cell receptor repertoire signature from the human peripheral blood

**DOI:** 10.1101/2024.12.10.24318751

**Authors:** Puneet Rawat, Melanie R. Shapiro, Leeana D. Peters, Michael Widrich, Koshlan Mayer-Blackwell, Keshav Motwani, Milena Pavlović, Ghadi al Hajj, Amanda L. Posgai, Chakravarthi Kanduri, Giulio Isacchini, Maria Chernigovskaya, Lonneke Scheffer, Kartik Motwani, Leandro Octavio Balzano-Nogueira, Camryn M. Pettenger-Willey, Sebastiaan Valkiers, Laura M. Jacobsen, Michael J. Haller, Desmond A. Schatz, Clive H. Wasserfall, Ryan O. Emerson, Andrew J Fiore-Gartland, Mark A. Atkinson, Günter Klambauer, Geir Kjetil Sandve, Victor Greiff, Todd M. Brusko

## Abstract

Type 1 Diabetes (T1D) is a T-cell mediated disease with a strong immunogenetic HLA dependence. HLA allelic influence on the T cell receptor (TCR) repertoire shapes thymic selection and controls activation of diabetogenic clones, yet remains largely unresolved in T1D. We sequenced the circulating TCRβ chain repertoire from 2250 HLA-typed individuals across three cross-sectional cohorts, including T1D patients, and healthy related and unrelated controls. We found that HLA risk alleles show higher restriction of TCR repertoires in T1D individuals. We leveraged deep learning to identify T1D-associated TCR subsequence motifs that were also observed in independent TCR cohorts residing in pancreas-draining lymph nodes of T1D individuals. Collectively, our data demonstrate T1D-related TCR motif enrichment based on genetic risk, offering a potential metric for autoreactivity and basis for TCR-based diagnostics and therapeutics.

## Introduction

Antigen-specific recognition of both foreign and auto-antigens is enabled by T-cell receptors (TCR). Elucidating the TCR sequences implicated in type 1 diabetes (T1D) is crucial for advancing our understanding of disease pathogenesis and facilitating the development of reliable biomarkers. Human leukocyte antigen (HLA) loci shape the TCR repertoire (DeWitt et al., 2018; Ortega et al., 2024; Zahid et al., 2024), and HLA risk alleles have been shown to influence TCR clonal sequences in various autoimmune diseases, including rheumatoid arthritis (RA), T1D, and celiac disease (CD) (Ishigaki et al., 2022; Nagafuchi et al., 2022). In T1D, HLA class II loci account for most genetic risk (Lambert et al., 2004; Noble et al., 1996), with DR3 and DR4 alleles conferring the highest risk (Noble & Valdes, 2011). Seropositivity for islet autoantibodies in combination with genetic risk can provide population-level estimates of the rate of T1D progression, with longitudinal studies illustrating the influence of HLA on type of initial seroconversion (e.g., insulin autoantibody [IAA] vs GAD autoantibody [GADA] first) and progression to multiple autoantibodies (Krischer et al., 2022). The development of T-cell biomarkers could enhance the monitoring of disease progression and improve current predictive methods for pre-symptomatic disease detection (Greissl et al., 2021). However, efforts to develop T1D biomarkers have been constrained by a reliance on pre-existing knowledge of autoimmune targets and cellular mechanisms in T1D (Ross et al., 2021). There remains a critical need for genetic studies that explore the link between TCRs and HLA in the context of T1D and for the identification of TCR-based biomarkers (Hanna et al., 2024).

Analyses of the adaptive immune receptor repertoire (AIRR) in infectious and autoimmune disease contexts have demonstrated that the TCR repertoire may be used to develop novel diagnostics (Arnaout et al., 2021; Emerson et al., 2017; Greiff et al., 2020; X. Liu et al., 2019; Mhanna et al., 2024; O’Donnell et al., 2024; Snyder et al., 2020; Yu et al., 2024). Longitudinal studies of healthy adult cohorts showed repertoire stability, indicating the robustness of TCR-based signatures for identifying sustained disease repertoire perturbations (Chu et al., 2019). However, autoimmunity-associated immune receptor signals, in contrast to those associated with infection and cancer, were initially found to be small or nearly undetectable when considering global repertoire metrics (Christophersen et al., 2014, 2019; Dahal-Koirala et al., 2022; Greiff et al., 2020; C. R. Weber et al., 2022). Therefore, novel analytical techniques, such as machine learning (ML) platforms, have been used to detect even very small shifts in signal within adaptive immune repertoires. For example, repertoire alterations have been documented in response to cytomegalovirus (CMV) (De Neuter et al., 2019; Emerson et al., 2017), cancer (e.g., lymphoma, tumor-infiltrating lymphocytes) (Ostmeyer et al., 2019; Schmidt-Barbo et al., 2024), therapeutic responses to checkpoint inhibitors in melanoma (Sidhom et al., 2022), as well as systemic autoimmune diseases including multiple sclerosis (MS) (Schneider-Hohendorf et al., 2024), systemic lupus erythematosus (SLE) (Nagafuchi et al., 2025), and RA (Komech et al., 2018; X. Liu et al., 2019) using statistical analyses and ML approaches on AIRR sequencing data with reasonable accuracy (area under the receiver operating curve [AUROC] > 0.75). The ML methods have demonstrated the capability to classify immune repertoires for a given clinical status and to recover immune signals associated with specific clinical conditions (Sidhom et al., 2021; Slabodkin et al., 2023; Widrich et al., 2020).

While previous reports indicate that islet autoantigen reactive cells are present at similar frequencies in the peripheral blood of control and T1D study participants (Culina et al., 2018; Eugster et al., 2015; Gomez-Tourino et al., 2017; Mitchell et al., 2022; Seay et al., 2016), others have observed increased expansion of antigen-specific clonotypes in T1D, indicating a disease signature in the periphery (Cerosaletti et al., 2017). The majority of islet-antigen specific complementarity determining region 3 beta chain (CDR3β) sequences were found to be private (Nakayama & Michels, 2021), or observed at the individual level, demonstrating a need for short consensus motif-based biomarkers.

We sought to identify a T1D-associated T cell repertoire signature from bulk peripheral blood mononuclear cells (PBMC) to provide a translationally relevant biomarker. To this end, we sequenced 2250 circulating TCRꞵ repertoires across the natural history of T1D to: (i) investigate the existence of HLA-restricted, T1D-associated CDR3β sequences and motifs, (ii) assess the feasibility to classify individuals as having T1D based on the TCR repertoire, and (iii) identify T1D-associated TCR signatures within the TCR repertoires. Our study reveals that the TCR repertoire lacks shared public clones across clinical groups. However, a significant enrichment of TCR repertoires at the subsequence level was observed, both with and without accounting for genetic risk factors. These findings suggest that TCR repertoire analysis may offer valuable insights into autoreactivity, potentially serving as a basis for developing TCR-based diagnostic tools and therapeutic strategies.

## Results

### Overview of the dataset and reproducibility assessment

To study T1D-associated TCR repertoire alterations, we immunosequenced the rearranged CDR3 TCRβ region in bulk PBMCs of three different cross-sectional cohorts (Fig. 1). Cohort 1 contains 1393 repertoires (103176 ± 26850 unique CDR3β sequences) distributed across the natural history of T1D from the University of Florida Diabetes Institute (UFDI) biobank. These include individuals diagnosed with T1D (n=426); first-degree relatives of T1D patients (FDR, n=625); second-degree relatives of T1D patients (SDR, n=59); unrelated healthy control individuals (CTRL, n=188), and islet autoantibody-positive individuals without diabetes who have increased risk of developing T1D (AAb+, n=95). Although statistically significant differences in unique CDR3β sequences were observed between clinical groups (<0.05), the difference in mean, median, and standard deviation was minimal (Fig. S1). Cohorts 2 and 3 respectively contain 679 T1D repertoires (112637 ± 55091 unique CDR3β sequences) and 178 deep sequenced CTRL repertoires (366271 ± 143197 unique CDR3β sequences), the latter sequenced with a different protocol than cohort 1 (see **Methods**). We obtained high reproducibility and adequate sequencing depth to capture the clonal diversity of TCRs using technical replicates (Fig. S2).

**Fig. 1.**
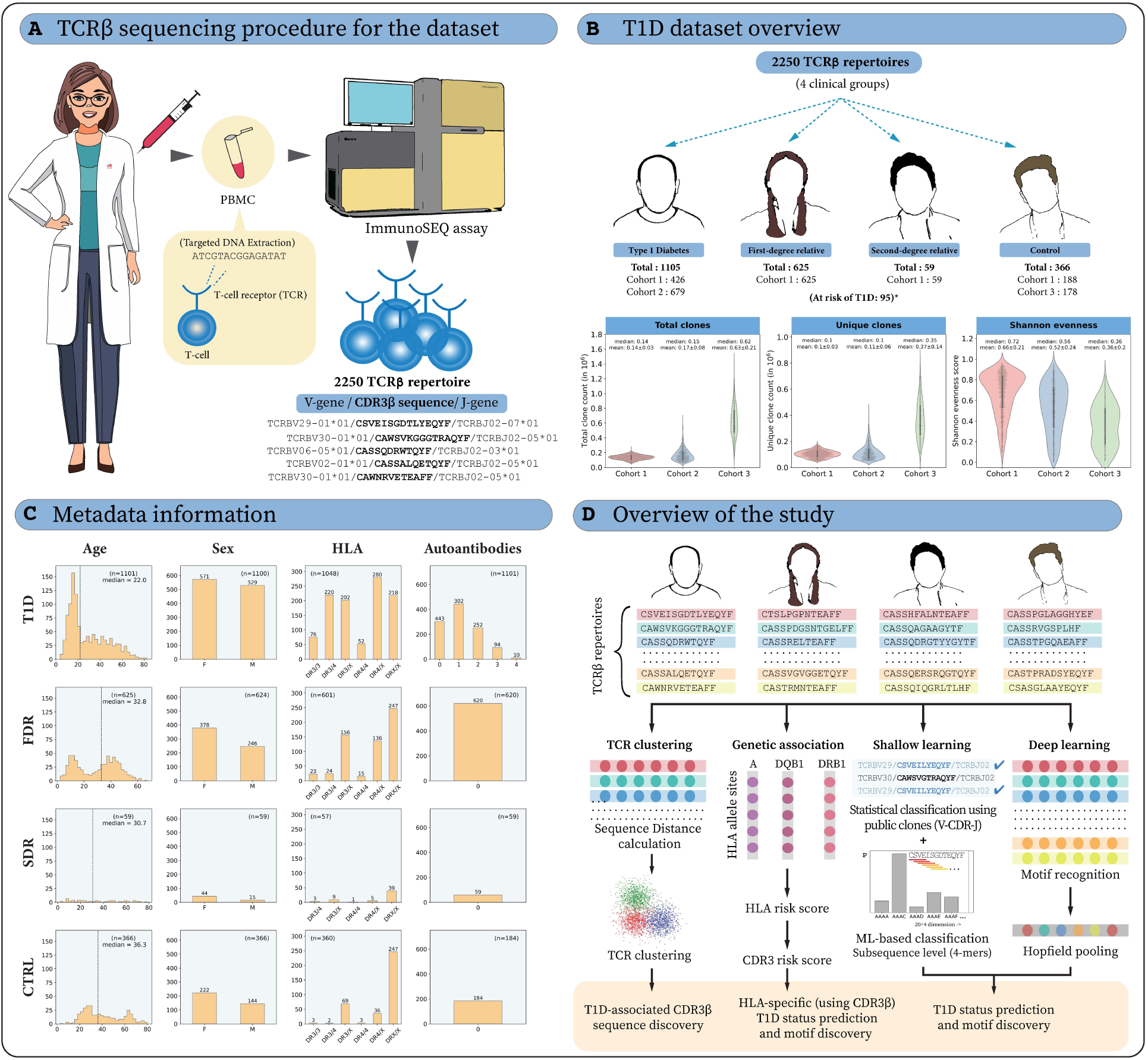
Overview of T1D TCRβ cohorts and the study. **(A)** DNA was isolated from peripheral blood mononuclear cells (PBMC) to conduct TCRβ chain sequencing. **(B)** The dataset of 2250 TCRβ repertoires contains four clinical groups: T1D (T1D), first-degree relatives (FDR), second-degree relatives (SDR), and non-related controls (CTRL) (*excluding 95 individuals without diabetes who have an increased risk of developing T1D, i.e. AAb+), which were sequenced in three different cohorts (see **Methods** section for cohort-specific sequencing details). The unique number of clones, total number of clones (unique clones and their clonal frequency) and Shannon evenness are shown for each cohort. Mean and median values are presented above each violin plot. **(C)** The distribution of age (dotted line shows median value), sex distribution, high-risk human leukocyte antigen (HLA) and number of autoantibodies (IAA, GADA, ZnT8A, IA-2A) present (for cohort 1 and 2) are shown for the four clinical groups across all three cohorts. **(D)** In this study, we leveraged available HLA information to conduct HLA and V-gene-restricted TCR clustering and to elucidate the extent to which high-risk HLAs restrict CDR3β sequences. The influence of HLA-mediated T1D risk was further translated into TCR repertoire-based T1D risk and identification of CDR3β motifs. Furthermore, we applied machine learning and deep learning (ML/DL) techniques to classify T1D and healthy related and unrelated controls. These computational approaches were also employed to identify T1D-associated TCRβ sequences and/or sequence motifs. **Supplementary Figures:** Fig. S1, Fig. S2

In addition to TCRβ sequencing, the dataset has extensive metadata in the form of whole genome (>978K single nucleotide polymorphism [SNP]) data derived from the UFDIchip, a custom Affymetrix array (Perry et al., 2023), clinical data (HbA1c, C-peptide, gender, age, T1D duration), and serological data (islet AAb number and specificity [IAA, GADA, zinc transporter 8 (ZnT8A), insulinoma associated protein-2 (IA-2A)] (M. D. Williams et al., 2021)) (Fig. S1). We employed precision genotyping (Perry et al., 2023) and HLA imputation (Jia et al., 2013) techniques to obtain 4-digit classical alleles for the major histocompatibility complex (MHC) class I and II genes. Specifically, we obtained 4-digit HLA for 1332 (95.6%) individuals including all AAb+ repertoires in cohort 1, 645 (95%) individuals in cohort 2, and 178 (100%) individuals in cohort 3. The majority of the primary analyses in this work were based on cohort 1 because it is the largest cohort among the three that also contains all clinical statuses. In contrast, cohorts 2 and 3 served as test cohorts for ML analysis unless specified otherwise.

### Repertoire-level similarity and diversity do not differ between T1D clinical groups

Repertoire-level similarity and diversity analyses are widely used for cross-patient comparisons of AIRRs (Chiffelle et al., 2020; Greiff et al., 2015; Mhanna et al., 2024; Miho et al., 2018; Vujović et al., 2023; C. R. Weber et al., 2022). We assessed immune repertoire diversity and similarity across clinical status within cohort 1 and restricted our analysis to the 779 individuals under age 30 to mitigate potential age-related confounding factors (Britanova et al., 2014). We assessed *TRBV*-gene usage (Fig. 2A) as previous studies in other autoimmune diseases (e.g., SLE, RA) revealed distinct V-gene usage patterns that may be associated with different disease states (Britanova et al., 2023; X. Liu et al., 2019). We observed a small number of statistically significant differences in V-gene usage, but absolute differences were biologically insubstantial. For example, we observed a significant difference in TCRBV19 (p-value: 1.14 * 10^-04^) gene usage across clinical groups. Yet, the difference in average frequency of V-genes in T1D (5.49 ± 0.58) and CTRL (5.75 ± 0.66) repertoires was minimal, in line with previous reports on other T1D cohorts (Gomez-Tourino et al., 2017).

**Fig. 2.**
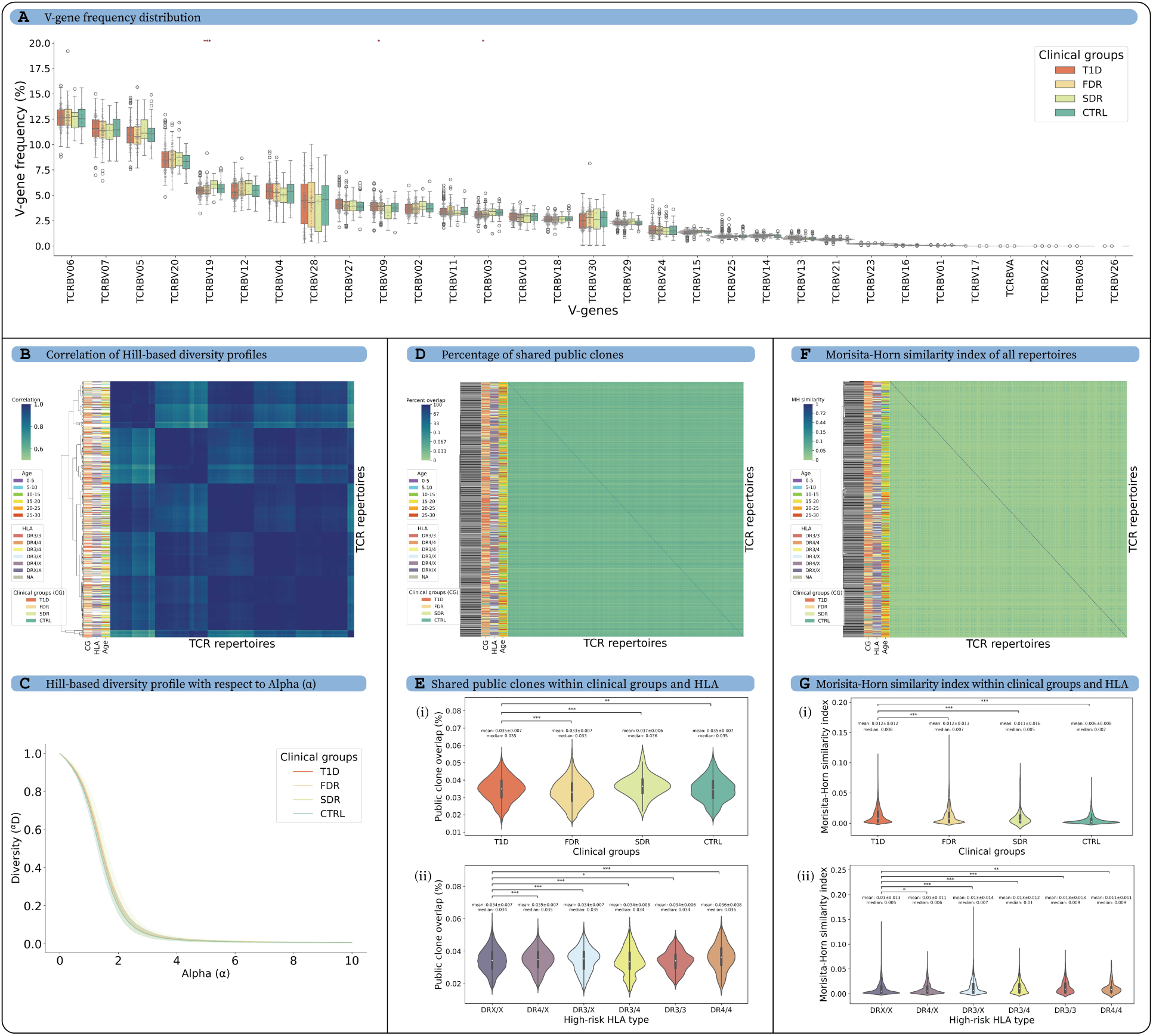
The V-gene distribution, diversity and clonal overlap of the TCRβ repertoires were similar across T1D, FDR, SDR, and CTRL groups. The analysis was performed on individuals in cohort 1 younger than 30 years of age, to minimize age-based confounding factors. **(A)** The V-gene distribution was similar across TCRβ repertoires of different clinical groups. The frequency of each V-gene was averaged for all repertoires in each clinical group. The difference in V-gene distribution was tested for all clinical groups using Kruskal–Wallis test, and p-values were adjusted for multiple testing using the Benjamini-Hochberg procedure. **(B)** Hill-based diversity profiles were calculated for alpha (α) value range [0,10] and step size 0.2, and Shannon evenness was used for the undefined value of α=1 for each repertoire. The heatmap shows the Pearson correlation of Hill-based diversity profiles for each pair of repertoires (see **Methods**). **(C)** The average Hill-based diversity profile for each α value of TCRβ repertoires calculated for different clinical groups. **(D)** The calculation of the percentage overlap of public clones involved determining the number of shared clones between two repertoires and dividing it by the total number of clones present in the smaller repertoire. The resulting values were used to create a heatmap that represents the percentage of public clones shared between each pair of repertoires. There was very low overlap among pairs of repertoires (max Public clones overlap = 0.08%). **(E)** The percentage of public clone overlap (i) among the clinical groups and (ii) among the high-risk HLA types (DR3/DR4). **(F)** The Morisita-Horn (MH) similarity index calculated for each pair of repertoire shows low overlap among TCRβ repertoires (Max MH similarity index = 0.34) and no clustering was observed based on age, clinical group, or high-risk HLA. **(G)** The MH similarity index (i) within the clinical groups and (ii) within the high-risk HLA types. All cluster heatmaps were generated using the UPGMA clustering method and Euclidean distance matrix. A universal color scheme was used to show T1D, FDR, SDR and CTRL in red, yellow, light green and dark green, respectively. The significance of difference in distribution was calculated using two tailed Mann–Whitney U tests with respect to T1D or non-high-risk HLA individuals (DRX/X), in respective plots. The p-values were adjusted using Benjamini–Hochberg method and described as * for [0.01,0.05], ** for [0.001,0.01] and *** for <0.001. Only significant p-values were displayed. The number of individuals in each group were as follows: (i) clinical groups: T1D: 367, FDR: 282, SDR: 29, CTRL: 101 and (ii) high-risk HLA types: DRX/X: 293, DR4/X: 150, DR3/X: 157, DR3/4: 77, DR3/3: 45, DR4/4: 23. In figure **D** and **F**, the linear color scale of the heat map was divided into 2 parts, where midpoint segregates the scale into two separate scales. The lower half encompasses the most common range of values (contains >90% data points) and the upper half accommodates values reaching up to the maximum.

To compare repertoires on the sequence level, we defined public clones as identical CDR3β sequences shared among two repertoires, ignoring the V- and J-gene information. To compare clonal expansion, we calculated the diversity of the immune repertoires using Hill-based diversity profiles (Greiff et al., 2015). The 𝛼-parameterized Diversity profile consolidates many previously established diversity indices (SR, ^𝛼=0^𝐷 (Gotelli & Colwell, 2001; Greiff et al., 2014); Shannon, ^𝛼=1^𝐷 (Estorninho et al., 2013; Shannon, 1948); Simpson’s, ^𝛼=2^𝐷 (Bashford-Rogers et al., 2013; Simpson, 1949); Berger- Parker, ^𝛼=∞^𝐷 (Berger & Parker, 1970; Greiff et al., 2014)). We calculated the correlation between the diversity profile of each repertoire and clustered them according to clinical groups, conventional high-risk HLA types (DR3, DR4), and age (Fig. 2B, C). However, the diversity profile did not cluster based on these parameters.

Furthermore, we calculated the similarity between repertories based on two metrics: (i) **percentage of shared public clones** for all repertoire pairs with respect to the total number of clones present in the smaller repertoire (based on clonal overlap) (Greiff et al., 2017) and (ii) **Morisita-Horn (MH) index** (based on clonal overlap and clonal frequency). Neither the public clones percentage nor MH index clustered based on clinical status, age, or high-risk HLA types (DR3, DR4) (Fig. 2D, F). Although the observations were statistically significant across clinical groups, the differences in median values were minor and lacked biological relevance for percentage of public clones (median values range from 0.033% to 0.036%) and MH index (median values range from 0.002 to 0.008). Similarly, conventional high-risk HLA alleles were not associated with biologically relevant differences in percentages of public clones (median values range from 0.034 to 0.036) and MH index (median values range from 0.005 to 0.01) (Fig. 2E, G). In summary, repertoire diversity and similarity analyses did not reveal biologically meaningful distinctions across clinical groups or HLA genetic risk factors.

### Inflammatory-disease-associated CDR3β sequences are overrepresented in T1D repertoires

Since no discernible repertoire-level similarity or diversity differences were observed across clinical groups without TCR-level antigen information, we next investigated if incorporating publicly available information on T1D and other disease-associated CDR3β sequences from the McPAS and VDJdb databases (<u>Table S1</u>) would show differences between clinical groups (Amoriello et al., 2021; Bagaev et al., 2020; Tickotsky et al., 2017). Using McPAS data, we found that T1D repertoires had a slightly higher percentage of T1D-associated CDR3β sequences (0.096 ± 0.047) compared to other clinical groups (FDR: 0.093 ± 0.073; SDR: 0.09 ± 0.061; CTRL: 0.094 ± 0.043 with Kruskal–Wallis test p-value of 0.0036; Extended Data Fig. 1A, <u>Table S2</u>). However, these T1D-associated CDR3β sequences were not sufficient to cluster the repertoires based on clinical groups, high-risk HLA, or age (Extended Data Fig. 1B). Similarly, we observed a significant overrepresentation of CD and influenza-associated CDR3β sequences in T1D repertoires (Cohn et al., 2014; Mitchell et al., 2023; Volta et al., 2011) (Extended Data Fig. 1A, <u>Table S2</u>). T1D and CD have shared genetic drivers, the most notable being shared high-risk HLA (DQ2.5/DQ8) (Gutierrez-Achury et al., 2015). Clinical guidelines emphasize vaccination for individuals with T1D, and the overrepresentation of influenza-associated CDR3β sequences might reflect increased vaccination compliance at the population level. Using VDJdb data, we observed a significant enrichment in hepatitis C virus (HCV), human immunodeficiency virus (HIV), and severe acute respiratory syndrome coronavirus 2 (SARS-CoV-2) associated sequences in T1D repertoires compared to other clinical groups (Extended Data Fig. 1C and <u>Table S2</u>). Despite the absence of a direct genetic link, several case reports have shown potential associations of T1D with HCV (L.-K. Chen et al., 2005; Masuda et al., 2007), HIV (Min-ChunYeh et al., 2023; Taguchi et al., 2023), and SARS-CoV-2 (Kendall et al., 2022). The SARS-CoV-2-specific CDR3β sequences observed in our cohort likely represent potential cross-reactive CDR3β sequences with other human coronaviruses, as all patient samples were collected before the COVID-19 pandemic.

The association of CMV with T1D is still unclear, with prior reports suggesting a lower (Ekman et al., 2019) and higher prevalence (Pak et al., 1988) of T1D in CMV patients. 2118 CMV-associated CDR3β sequences in McPAS observed no significant association with T1D status in our study. We further utilized a collection of 25,508 unique CDR3β sequences associated with CMV exposure curated from the literature (May et al., 2024) and observed a significant reduction in the presence of CMV-associated CDR3β sequences in the T1D repertories compared to other groups in cohort 1 (Extended Data Fig. 1D). Our observation supports findings of a recent study performed on a larger cohort (Ekman et al., 2019) suggesting early childhood CMV infection decelerates the progression to clinical T1D. In summary, we validated that CDR3β sequences previously linked with T1D are associated with clinical groups within our cohort. However, the strength of these associations was minimal, highlighting the need for a more comprehensive analysis using advanced methodologies. Furthermore, we identified potential links between T1D progression and several other diseases, including CD, influenza, HCV, HIV, SARS-CoV-2, and CMV.

### HLA risk alleles demonstrate higher restriction of TCR repertoire diversity in T1D individuals

#### Limited T1D association of HLA-associated public TCRs

Several autoimmune diseases, including T1D, RA, CD, and SLE, have a substantial component of genetic risk driven by HLA class II genotype (García et al., 2021, 2022; Hu et al., 2015; Ilonen et al., 2002; Noble & Valdes, 2011; Pociot & Lernmark, 2016; Smith et al., 2024). Given the potential implications of high-risk HLA alleles in modulating thymic selection of autoreactive clones (Ashby & Hogquist, 2024; ElAbd et al., 2025) or in promoting expansion of these clones in the periphery (Pugliese, 2017) (Fig. 3A), several computational models and statistical methods were recently developed to predict the association between the TCRs and HLA (S. Liu et al., 2023a; Ortega et al., 2024; Zahid et al., 2024). Thus, in an attempt to control for HLA differences among clinical groups and probe for T1D-specific associations of TCRβ receptor features, we first identified strongly HLA-associated TCRβ receptor features within cohort 2 with the intent of testing those features prevalence and enrichment with T1D status in cohort 1 (see Q1, Fig. 3B).

**Fig. 3.**
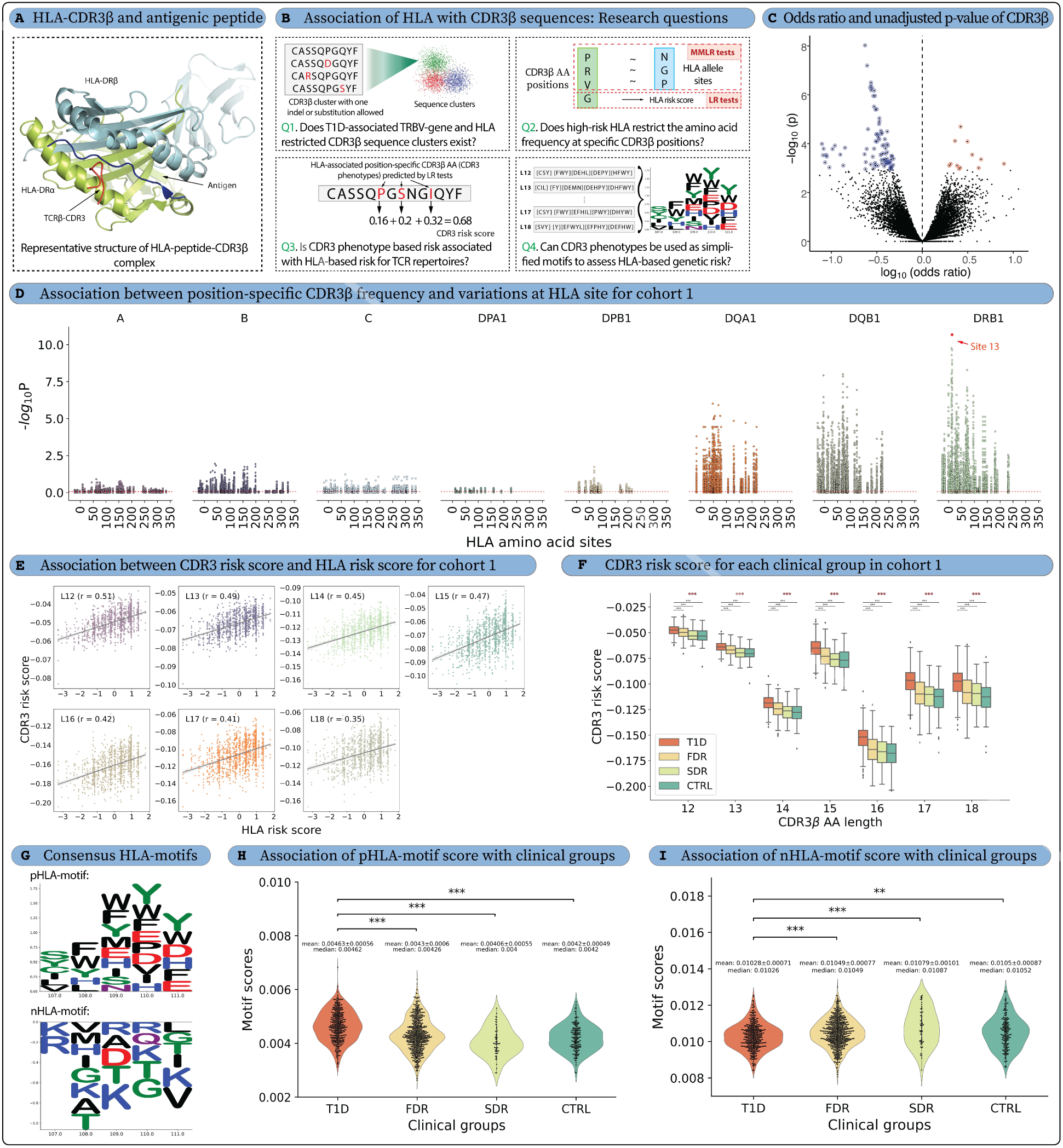
High-risk HLA alleles restrict the AA frequency of CDR3β sequences. **(A)** A representative complex structure of the human CDR3β region in close proximity to Influenza HA Antigen Peptide and MHC Class II Molecule, HLA-DR4 (PDB id: 1J8H), showcasing the potential impact of HLA on CDR3β sequences **(B)** This study addressed four key questions related to HLA restriction: **Q1** finds the T1D-associated TRBV-gene and HLA-restricted full length CDR3β sequences that are overrepresented or depleted in the TCR repertoires, **Q2** investigates the restriction of CDR3β AA frequencies by HLA leading to identification of HLA-associated position-specific CDR3β AAs (CDR3 phenotypes), **Q3** calculates the risk score for each repertoire based on CDR3 phenotypes to compares it with HLA-based risk score and clinical groups, and **Q4** identifies the positively- and negatively-associated HLA-motifs from CDR3 phenotypes and assesses their association with clinical groups, classical high-risk HLA alleles (DR3/DR4), autoantibody presence and T1D duration. The first three questions were addressed for all TCR repertoires in cohort 1 with available HLA information (n=1242) and individually for each clinical group, including T1D repertoires (n=402), FDR repertoires (n=601), and CTRL repertoires (n=182) to assess the generalisability and robustness of the analysis. The number of SDR (n=57) repertoires was insufficient to obtain significant CDR3 phenotype and, therefore, SDR were excluded from this analysis. **(C)** We observed few TRBV-gene family- and HLA-associated CDR3β sequences depleted (blue) or overrepresented (red) in the T1D repertoires vs FDR, SDR, CTRL in Cohort 1, when considering nearly-identical CDR3 with one indel or substitution allowed (Q1). The plot shows odds ratio on x-axis and unadjusted p-values of y-axis. **(D)** The MANOVA test p-values from the MMLR analysis for the 1242 repertoires (cohort 1) showed association between CDR3 positions and HLA sites, where variation in CDR3β AA frequency at each CDR3β position was plotted for each HLA site containing mutation(s) (Q2). An LR model was also developed with HLA risk score and CDR3β AA frequency to identify CDR3 phenotypes. These CDR3 phenotypes were utilized to compute CDR3 risk score and HLA-motifs from TCR repertoire. **(E)** A significant correlation was observed between T1D-associated HLA risk scores and CDR3 risk scores calculated for each repertoire in cohort 1 (Q3). **(F)** CDR3 risk score was also higher for the T1D clinical group and observed the expected trend where T1D>FDR>SDR>CTRL (Q3) **(G)** The positively- (pHLA-motif) and negatively- (nHLA-motif) associated HLA-motif obtained from the CDR3 phenotypes, where **(H)** pHLA-motif showed higher presence in T1D repertoires and **(I)** nHLA-motif showed expected opposite trend with higher presence in CTRL repertoires. In the figure, Multiple testing was performed using Kruskal–Wallis test (denoted with red stars), and p-values were adjusted between different CDR3β lengths. p-values for pairwise testing were calculated using two tailed Mann-Whitney U tests, and p-values were adjusted between different clinical groups. All p-values were adjusted for multiple testing using the Benjamini–Hochberg method. p-values were described as * for [0.01,0.05], ** for [0.001,0.01], *** for <0.001, and no stars plotted for non-significant values. **Supplementary Figures:** Extended Data Fig. 2, Extended Data Fig. 3, Extended Data Fig. 4, Extended Data Fig. 5, Fig. S4, Fig. S3, Fig. S5, Fig. S6, Fig. S7, Fig. S8, Fig. S9

We tabulated the publicity of all unique cohort 2 TCRs across the cohort 2 repertoires, defining detection as an exact or near-exact match (1 CDR3 AA variation), and computed the odds ratio (OR) of detection in participants with and without each common HLA allele. A TCR feature was assigned to the HLA allele with the lowest p-value (Fisher’s exact test). To limit the number of hypotheses tested in the next stage, we filtered TCR features with p-values < 1e-8 and detected in at least 5% of HLA-matched individuals and not more than 10% of HLA-mismatched individuals. This yielded 20,037 unique V-gene family-CDR3β centroids assigned to either MHC class I allele (n = 7,255) or MHC class II (n = 12,782) allele(s) (Fig. S4, <u>Table S3</u>). To determine whether these features were associated with T1D status, we then searched for these features in cohort 1 among participants with the hypothesized restricting HLA allele (Fig. S3). After correcting for multiple hypothesis testing, 39 HLA-DRB1*03 associated TCRs were significantly underrepresented (False discovery rate < 0.2) among the T1D group (Fig. 3C, Extended Data Fig. 2A, Extended Data Fig. 2C). Twelve strongly HLA-associated (B*40:01, B*15:01, DQA1*01:02, DQA1*05:01, DQB1*02:01) TCR features were also overrepresented (False discovery rate < 0.2) in the HLA-matched T1D repertoires versus controls (Extended Data Fig. 2B).

The TCRβ feature most underrepresented in cohort 1 T1D repertoires was TRBV7 CASSLSLAGSNNEQFF, with it or its near-exact neighbor detected in 31.5% (80/254) control and 9.3% (18/192) T1D repertoires of persons expressing the DRB1*03:01 and/or DQA*05:01:DQB*02:01. Many of the other TCR features, strongly underrepresented in T1D repertoires (blue circles in Fig. 3C), also had a similar CDR3β sequence (1-3 mutation distance from TRBV7 CASSLSLAGSNNEQFF) as shown in motifs (Extended Data Fig. 2C). Two of these sequences, V07,CASSLSLAGTYNEQFF and V07,CASSLSLAGAYNEQFF, were previously identified as TCRβ sequences associated with DRB1*03:01 status by statistical testing in a healthy population (DeWitt et al., 2018). These features were observed in repertoires across all age groups among controls; however, because the population without diabetes (FDR/SDR/CTRL) is older than the T1D diagnosed group, we cannot exclude the possibility that the TCR features most depleted in cohort 1 T1D repertoires may reflect age-dependent acquisition of T cell memory due to recurrent exposures and vaccinations. Overall, despite finding many TCRs associated with known T1D-associated HLA risk alleles, we noted little definitive signal of HLA-restricted TCRs enriched in repertoires based on T1D clinical status after conditioning to persons with the relevant allele.

#### Amino acid (AA) variation in HLA risk alleles restricts the position-wise AA frequency of the TCR repertoire

##### A statistical framework to assess HLA-based TCR repertoire restriction

Investigations into the genetic basis of type 1 diabetes (T1D) have increasingly highlighted the HLA region as pivotal in shaping immune tolerance and susceptibility (ElAbd et al., 2025; Nakayama et al., 2005). A previous study on 18,832 T1D case-control samples identified three AA sites in HLA-DQβ1 (site 57) and HLA-DRβ1 (sites 13 and 71) as the main drivers of T1D risk (Hu et al., 2015). These three positions together explained 90% of the T1D-specific phenotypic variance in the HLA-DRB1–HLA-DQA1–HLA- DQB1 locus and 80% of the variance explained by the entire MHC region. Ishigaki et al. (Ishigaki et al., 2022) further observed that the polymorphisms in HLA alleles alter the T-cell repertoire. The variation of AA in the above three HLA sites (in DQβ1 and DRβ1) and associated log odd scores were used to calculate the genetic risk of T1D from HLA in our cohorts, termed the HLA risk score (Hu et al., 2015) (see **Methods**). A TCR sequence-based disease-associated risk score (termed CDR3 risk score) was also established, which quantifies the association of AA variation in HLA alleles and positional AA variation in CDR3β sequences (Ishigaki et al., 2022).

We computed CDR3 risk scores for all TCR sequences in our most comprehensive cohort 1 containing all clinical groups and then, tested our observations in cohorts 2 and 3 comprised of T1D and CTRL individuals, respectively: (i) To examine the association of CDR3β sequences with HLA allele-based genetic risk, (ii) to identify the high-risk HLA-associated CDR3β AA, utilize them to calculate a repertoire-level CDR3 risk score, and assess whether this score exhibits an association with clinical groups, and (iii) to identify CDR3β sequence motifs that encapsulate HLA-based T1D risk within TCR repertoires (see Q2- 4; Fig. 3B).

In keeping with previous work (Ishigaki et al., 2022), we refer to AA locations in HLA as ‘sites’ and AA locations within CDR3β as ‘positions’. Across eight HLA genes, we identified 398 HLA sites that exhibited AA variation (Extended Data Fig. 3A). The observed variations in the HLA sequences were distributed almost equally between HLA class I genes (218 variations, 54.8%) and HLA class II genes (180 variations, 45.2%). For the computation of a CDR3 risk score, only the CDR3β lengths 12 to 18 (denoted as L12 to L18) were considered for all HLA-CDR3β association analyses as they were the most frequent CDR3β lengths in all repertoires (Extended Data Fig. 3F). To compute the CDR3 risk score, the AA frequencies were calculated for each position of the CDR3β sequences, where 7 AA positions were present for L12 with increments of one position up to L18, due to removal of TRBV- and J-gene encoded regions, leading to a total of 70 CDR3β positions (Extended Data Fig. 3B). We divided the cohort 1 repertoires into four subsets: (i) total cohort 1 (contains all clinical statuses, 1242 repertoires), (ii) T1D only (402 repertoires), (iii) FDR only (601 repertoires), and (iv) CTRL only (182 repertoires). All the analyses in this section were performed for all four cohort 1 subsets. SDR (57 repertoires) were not analyzed as no significant T1D- associated CDR3β AAs were observed in the dataset due to low repertoire count.

##### HLA sites restrict the position-specific AA diversity of CDR3β sequences

We first validated that CDR3β sequence diversity was restricted as a function of HLA sites in cohort 1, as observed in the previous study (Ishigaki et al., 2022). We obtained AA variants for each HLA site from HLA genotyping. To quantify the association between each HLA site and CDR3β position, multivariate multiple linear regression (MMLR) was used, where a multidimensional vector of CDR3β position-specific AA frequency (response variable) was used to predict the association with multidimensional vector containing polymorphisms at each HLA site (explanatory variable) (Ishigaki et al., 2022) (Extended Data Fig. 3C). To test interindividual variance in CDR3 AA frequencies explained by the HLA genotype, we subsequently evaluated the MMLR model using a multivariate analysis of variance (MANOVA) (Fig. 3D). A total of 27860 MMLR-MANOVA tests were performed (398 HLA sites x 70 CDR3β positions), and 13313 significant associations (47.8% of total tests; FDR<0.05) were observed after false discovery rate correction using Benjamini-Hochberg adjustment (<u>Table S4</u>). There were 5022 (37.7%) significant associations from class I HLA types and 8291 (62.3%) from class II HLA types. The number of significant associations was more than two times higher than those reported previously (Ishigaki et al., 2022), as our dataset was also almost double in size. The lowest p-value was observed between *HLA-DRB1* site 13 (p-value = 3.1 * 10^-214^), located within the T1D risk-associated peptide binding groove (Hu et al., 2015), and the CDR3β position 111 for L15. Position 109 in L13 (5^th^ position in our analysis with p-value = 4.7 * 10^-180^) was reported in a previous study as having the lowest p-value (Ishigaki et al., 2022). Both CDR3β positions were in the antigen-binding region. Separately, we performed MMLR analysis on each T1D, FDR, and CTRL repertoire subset to observe similar HLA-based CDR3β restriction in each clinical group (Fig. S5, Fig. S6 and **Supplementary Note**). Taken together, (i) we confirmed the observation from Ishigaki et al. (Ishigaki et al., 2022) that AA polymorphisms in HLA restrict the AA positional frequencies of the CDR3β sequences and (ii) extended these observations to T1D, FDR, and CTRL.

##### HLA-risk-allele-linked CDR3 risk score is highest in T1D individuals

Building on observed associations between variation in HLA sites and AA frequencies within the CDR3β sequences, we next investigated the influence of HLA risk score on position-specific CDR3β AAs (hereafter referred to as CDR3 phenotypes) in TCR repertoires. We first calculated the HLA risk score for each repertoire in cohort 1 (see **Methods**, Extended Data Fig. 3D, <u>Table S5</u>). To identify CDR3 phenotypes associated with the HLA risk score, we conducted a total of 1400 (70 CDR3β positions x 20 AA frequencies at each position) linear regression (LR) tests with HLA risk score (Extended Data Fig. 3E, <u>Table S6</u>). There were a total of 529 CDR3 phenotypes with significant association with T1D risk for cohort 1 (p-value ≤0.05) (Fig. S5). The correlation coefficients from these LR tests were treated as the effect sizes for the respective CDR3 phenotypes, which were further used to calculate the CDR3 risk score by summing up the effect sizes of each CDR3 phenotype for each CDR3β sequence and calculating the average value for the whole repertoire. The CDR3 risk score reflects HLA-based T1D risk in the TCR repertoire, and this relationship was validated by correlating it with the HLA risk score across different CDR3β sequence lengths (correlation coefficient, r = 0.35 to 0.51, Fig. 3E). The HLA-associated CDR3 phenotypes (AAs) exhibit both positive and negative effect sizes. CDR3 risk scores based solely on positive or negative effect sizes showed comparable correlations (r = 0.27 to 0.49) to the combined CDR3 risk score (Fig. S5, Fig. S6). Additionally, average CDR3 risk score was highest for the T1D repertoires followed by FDR, SDR, and CTRL repertoires, respectively, across different lengths (Fig. 3F). A similar observation was also exhibited by classical high-risk HLA allele types, where presence of at least one DR3 or DR4 allele had higher CDR3 risk score compared to other non-T1D risk allele types (DRX/X) (Fig. S7). We assessed the robustness and factors influencing the CDR3 risk score on cohort 1 by analyzing the overlap of CDR3 phenotypes across clinical groups and assessing the significance of CDR3 phenotype effect sizes, respectively (see **Supplementary Note** and Fig. S8, <u>Table S7</u>). CDR3 phenotypes obtained from cohort 1 were also tested on cohorts 2 and 3, where the CDR3 risk score was significantly higher for the T1D repertoires present in cohort 2 compared to CTRL repertoires present in cohort 3, across different CDR3β length (Fig. S9). The significant correlation with HLA risk score and association with clinical groups were also validated for CDR3 risk score, across each subset of cohort 1 (Fig. S5, Fig. S6).

To summarize, we identified 529 HLA-associated CDR3 phenotypes that were selectively restricted by high-risk HLA alleles associated with T1D. These CDR3 phenotypes and their effect sizes were used to calculate the CDR3 risk score exclusively based on TCR repertoires. A robust association between CDR3 risk score, HLA risk score, and T1D clinical status validates the role of HLA-mediated restriction in shaping TCR repertoires. Additionally, these CDR3 phenotypes were also validated in cohorts 2 and 3.

##### HLA-associated CDR3β motifs provide a simplified representation of high-risk HLA-based CDR3 restriction

We identified a consistent recurring pattern in charged and aromatic AA in both positively- and negatively-associated CDR3 phenotypes of different CDR3β length (see **Methods**, Fig. S5A, <u>Table S8</u>). The pattern was utilized to derive a simplified representation of the HLA-associated T1D signature within TCR repertoires as positively-associated HLA-motif (pHLA-motif) and negatively-associated HLA-motif (nHLA-motif) (Fig. 3G). We further utilized respective HLA-motifs to calculate an HLA-motif score for each repertoire based on the total number of CDR3β sequences containing these motifs. The pHLA-motif score was significantly higher for T1D (0.00463 ± 0.00056, p-value<0.001) versus FDR, SDR, and CTRL groups (values ranging from 0.00406 to 0.0043) (Fig. 3H, Extended Data Fig. 4). A similar trend was observed for the T1D individuals in cohort 2 (0.00451 ± 0.00057) and CTRL in cohort 3 (0.00404 ± 0.00053). On the other hand, nHLA-motif scores were lower for T1D (0.01028 ± 0.00071) repertoires compared to other clinical groups (values ranging from 0.01049 to 0.01079) (Fig. 3I, Extended Data Fig. 5). Again, a similar trend was observed for the T1D individuals present in cohort 2 (0.0103 ± 0.0008) and CTRL in cohort 3 (0.011 ± 0.0008).

The expected trends for pHLA-motif score (T1D>FDR>SDR>CTRL) and nHLA-motif score (T1D<FDR<SDR<CTRL) were observed, even when at least one HLA allele was non-risk (DRX) and when no islet autoantibody presence was detected (Extended Data Fig. 4, Extended Data Fig. 5). However, the differences observed for the nHLA-motif score were less pronounced. pHLA-motif score showed an expected negative correlation with T1D duration and T1D patient age (Pearson correlation ranging from - 0.3 to -0.44), likely due to a reduction in β-cell antigens (C. L. Williams et al., 2022). However, the nHLA- motif score had a low and inconsistent correlation (Pearson correlation ranging from -0.12 to 0.11) with T1D duration and/or patient age. In conclusion, the distinct pHLA- and nHLA-motif scores reflect the influence of HLA-associated T1D risk on TCR repertoires, suggesting these motifs as potential markers of HLA-driven susceptibility and protection in T1D.

#### The presence of heterozygous HLA alleles restricts TCR diversity

A recent murine study reported that the presence of heterozygous MHC class II alleles constrains the diversity of the TCR repertoire (Brown et al., 2024). To investigate this observation in humans, we tested the hypothesis in our dataset. First, we calculated the generation probability (Pgen) using Igor, a tool that models the V(D)J recombination process to estimate the likelihood of generating a specific TCR or B-cell receptor (BCR) sequence (Marcou et al., 2018). Pgen reflects the probability of a sequence being produced purely by the random recombination of V, D, and J gene segments, along with nucleotide insertions and deletions. Subsequently, we calculated the post-selection probability (Ppost) using SoNNia, which distinguishes sequences that have undergone selection. In our study, Pgen and Ppost were used as proxies to assess TCR repertoire diversity (Isacchini et al., 2021).

Our analysis revealed a weak trend in cohort 1, where increased HLA heterozygosity appeared to be associated with reduced TCR diversity, as indicated by both Pgen and Ppost (Extended Data Fig. 6A, B). This trend was not observed in cohorts 2 and 3, potentially due to HLA biases arising from presence of only T1D repertoires in significantly larger cohort 2 and fewer CTRL repertoires in cohort 3, respectively. Additionally, we observed that frequencies of certain V-genes (e.g., TRBV28, TRBV4-3, TRBV3-2, TRBV3-1) clustered into a distinct group (Extended Data Fig. 6C). We used TRBV28 as a case study and observed V-gene frequencies as low-frequency (f < 0.015), mid-range frequency (0.015 ≤ f ≤ 0.04), and high-frequency (f > 0.04) clusters. Although a direct association between V-gene frequency clusters and specific clinical groups was not identified, a potential link with certain HLA alleles was observed. (Extended Data Fig. 6D). However, these V-gene frequency clusters could be attributed to confounding factors, such as age, ethnicity, infection history, or sequencing artifacts, which fall outside the scope of the current study. These observations also contrast a previous study on 666 individuals, indicating HLA (class I)-heterozygous individuals present a broader immunopeptidome for recognition by cytotoxic T cells (Krishna et al., 2020). Altogether, our findings were consistent with the observations from the murine study (Brown et al., 2024), where HLA heterozygosity was shown to limit TCR repertoire diversity.

### Machine learning enabled classification of T1D status using TCR repertoires

The classification of immune repertoire for disease status prediction is based on the principle that disease signatures (identical or similar) are shared across individuals affected with the same disease (Mhanna et al., 2024). However, there are several approaches to identify disease signatures associated with disease (M. Liu et al., 2022). In our study, we used previously utilized HLA risk scores (Hu et al., 2015) and three different ML/deep learning (DL) approaches, namely, (i) public clone (V-CDR3-J)-based shallow ML, (ii) k-mer based shallow ML, and (iii) attention-based DL, to classify the T1D repertoires. Across all ML approaches, T1D status was considered the positive class and the remaining clinical groups (FDR, SDR, CTRL, together termed “no diabetes” [ND]) were considered non-T1D or negative class for training, tuning, and testing within cohort 1. The combined set of cohorts 2 and 3 (only T1D and only CTRL repertoires, respectively) was used as a held-out test set. Due to class imbalance, we used AUROC as the primary metric for evaluating predictive performance. Moreover, a confounder adjustment procedure was applied for DeepRC and k-mer based logistic regression (LogReg) models through a series of steps aimed at adjusting the impact of age on the classification of subjects based on their T1D status (see **Methods**).

#### HLA-based classification serves as a strong baseline model for predicting T1D status

We first used classical high-risk HLA alleles, DR3 and DR4, to assess T1D status classification. As expected, we found that the presence of high-risk HLA alleles (DR3 and/or DR4) alone is insufficient to accurately predict the clinical status of T1D (see **Supplementary Note,** Extended Data Fig. 7A), reflective of the complex, multifactorial nature of this disease (Noble & Erlich, 2012). However, homozygosity for either DR3 or DR4 is a strong predictor of T1D status (91.4% in cohort 1 and 97.2% in cohort 2). We also utilized HLA risk score (Hu et al., 2015) to classify the T1D status (Fig. 4A). The AUROC curve was 0.73 for cohort 1 and 0.85 for the held-out test set (cohort 2 and 3) (Fig. 4B). However, balanced accuracy was ∼67% for both training and held-out test datasets, due to class imbalance.

**Fig. 4.**
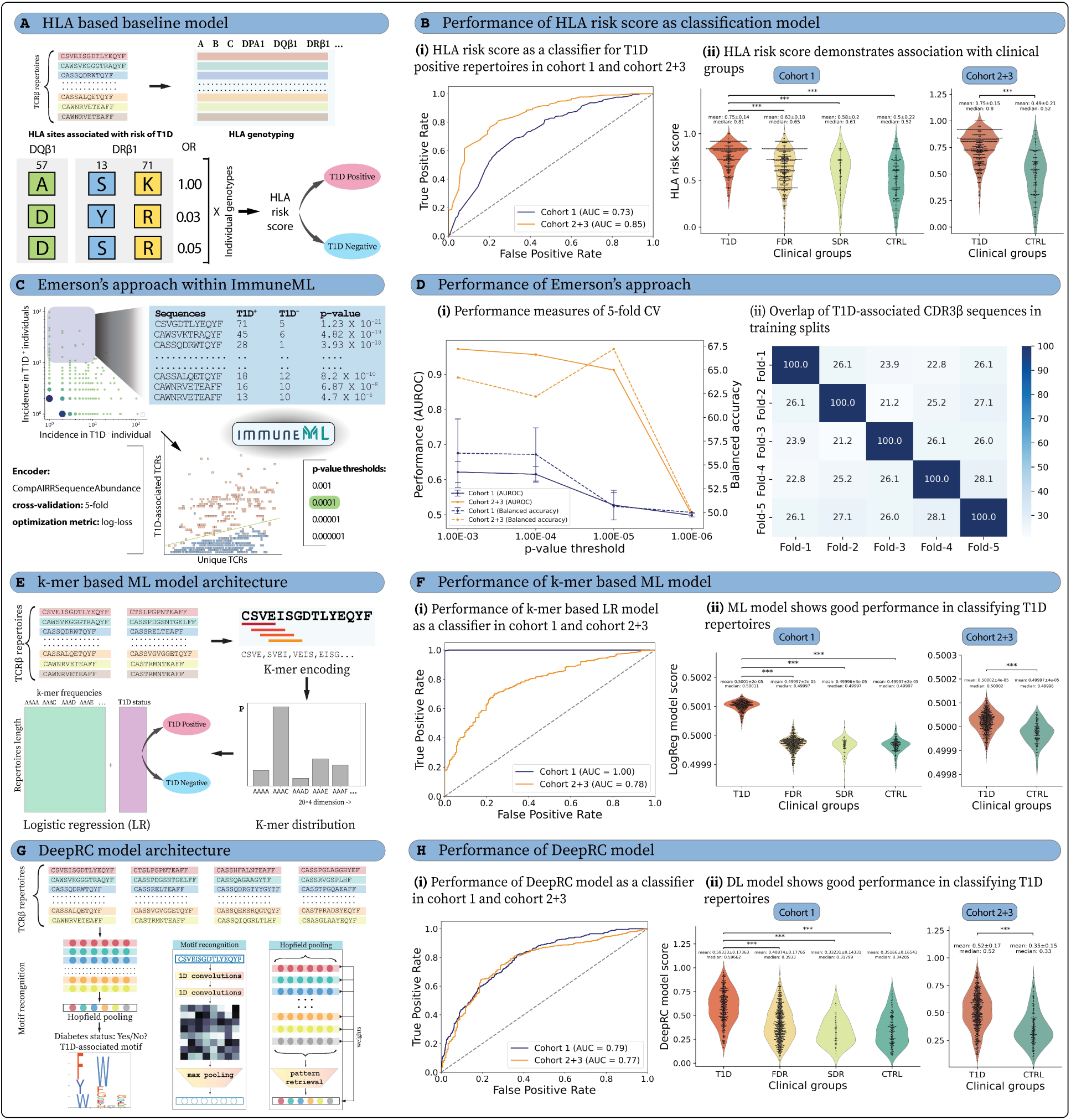
The application of different repertoire classification methods to classify T1D and non-T1D repertoires shows varying levels of prediction performance. **(A)** Schematic representation of T1D status classification using the HLA risk score (baseline model). HLA risk score calculated based on 3 AA positions (HLA-DQβ1: site 57 and HLA-DRβ1: site 13 and 71) was utilized to classify T1D and non-T1D repertoires. **(B)** HLA risk score was used for classification in cohort 1 and cohort 2+3, where (i) AUROC was 0.73 for cohort 1 and 0.85 for cohort 2+3 and (ii) HLA risk score was higher for T1D clinical group with expected trend of T1D>FDR>SDR>CTRL. **(C)** The immuneML platform was utilized to replicate the approach utilized by Emerson et al., (Emerson et al., 2017), where T1D-associated TCRβs were identified and used in a classification model. **(D)** The model was trained on Cohort 1 based on log-loss calculations and 5-fold cross-validation (CV) and tested on cohort 2+3, where (i) AUROC was significantly lower for cohort 1 compared to cohort 2+3 for all p-value thresholds and (ii) overlap of identified T1D-associated sequences across different splits was also relatively low (21% - 28%). **(E)** Schematic representation of repertoire classification using k-mer frequency as a feature in machine learning models. We calculated the frequencies of 4-mers to create a matrix of n*20^4^, where n is the number of repertoires. We further used Logistic regression (LogReg) for the classification of T1D positive and negative repertoires. **(F)** The LogReg prediction showed overfitting in cohort 1, where (i) AUROC of cohort 1 was 1.0 and reduced to 0.775 for cohort 2+3. (ii) Overfitting was also visible when LogReg predictions in cohort 1 were stratified based on clinical groups. T1D score was significantly higher compared to the non-T1D clinical groups and almost no difference was observed between FDR, SDR and CTRL in cohort 1. **(G)** Schematic representation of the Deep learning approach (DeepRC) used for T1D disease status classification and DeepRC-motif identification. TCRβ repertoires were represented as collections of CDR3β sequences, which exhibit variability between individuals. A convolutional neural network (CNN) model was implemented to recognize motif patterns in each of the sequences and map them to sequence-representations. Subsequently, a pooling function aggregated these sequence representations into a repertoire-level representation. **(H)** DeepRC was more reliable compared to the LogReg model. (i) The AUROC values were similar for both cohort 1 (0.79) and cohort 2+3 (0.77) and (ii) DeepRC predictions also observed the expected trend of T1D>FDR>CTRL with SDR having the least score. In all violin plots, p-values for pairwise testing with respect to the T1D clinical group were calculated using two tailed Mann-Whitney U tests and p-values were adjusted between different clinical groups using the Benjamini–Hochberg method. p-values were described as * for [0.01,0.05], ** for [0.001,0.01] and *** for <0.001 and no stars plotted for non-significant values. **Supplementary Figures:** Extended Data Fig. 7, Extended Data Fig. 8, Extended Data Fig. 9, Fig. S10, Fig. S11, Fig. S12

We further investigated the performance of the CDR3 risk score, which was essentially an effect of high-risk HLA on the TCR repertoire. As the CDR3 risk score was calculated for different CDR3β lengths, we obtained AUROC values between 0.65 to 0.75 (balanced accuracy ranging from 50.4% to 65.8%) on cohort 1 and 0.66 to 0.80 (balanced accuracy ranging from 52% to 72.7%) on cohort 2+3 (Fig. S10). The pHLA and nHLA motifs can be considered a simplified output of genetic association study, and therefore, tested for their classification performance. The pHLA motif obtained AUROC of 0.68 (balanced accuracy 64.8%) on cohort 1 and 0.73 (balanced accuracy 68.6%) on cohort 2+3. Whereas, nHLA motifs expectedly had comparatively lower AUROC of 0.59 (balanced accuracy 50.2%) on cohort 1 and 0.73 (balanced accuracy 50%) on cohort 2+3 (Fig. S10). The performance metrics for the aforementioned classifiers are also summarized in the supplementary (<u>Table S9</u>).

#### The presence of public clones is insufficient for the classification of T1D status

Emerson and colleagues (Emerson et al., 2017) developed a statistical classification framework that could diagnose CMV status from peripheral blood TCRβ sequences. Their approach identified the statistically significant enrichment of TCRβ sequences in CMV^+^ compared to CMV^−^ individuals. We applied this approach on the T1D dataset using our open-source ImmuneML (Pavlović et al., 2021) platform (Fig. 4C). The model performance was tested on 5-fold cross-validation (CV), where log-loss value was predicted to be the least at p-value threshold of 0.0001 (log-loss 0.417 for training and 0.4 for test; Fig. S11, <u>Table S10</u>). The balanced accuracy and AUC values of the model at the optimal p-value threshold were low for the training cohort 1 (average AUROC: 0.615 and balanced accuracy: 56.1% across 5 splits) (Fig. 4D). Testing the model on cohort 2+3 showed relatively high AUROC (0.956) and balanced accuracy (62.2%). However, increased performance in the test dataset was likely due to the sequencing depth bias between cohorts 2 and 3 since test cohort precision (100%) was significantly higher than recall (24.4%) (Fig. S11). A total of 140 TCR clones were predicted to be associated with T1D (<u>Table S11</u>). A low overlap, ranging from 21% to 28%, was observed among T1D-associated TCRβ sequences across each CV split (Fig. 4D). In summary, the statistical classification approach based on public clones was insufficient to classify repertoires, due to the low prevalence of public clones.

#### K-mer based logistic regression failed to differentiate clinical groups

In the k-mer based repertoire classification approach, a sliding window of 4-mers was employed for each CDR3β sequence, and the occurrence of k-mers was calculated for each immune repertoire (see **Methods**). The k-mer frequency matrix was utilized as a feature in logistic regression (LogReg)-based ML models. The final model was obtained by averaging the output of 5-folds into a single model after sigmoidal activation (Fig. 4E). There can be several confounding factors related to T1D affecting the TCR repertoires (Conrad et al., 2023). The dataset was age-corrected by applying sample weights to avoid more prominent age-related confounding factors (see **Supplementary Notes**) (Zaslavsky et al., 2024). The LogReg model was fitted on the training dataset (cohort 1) and yielded an AUROC of 0.78 on the held-out test dataset (cohort 2 and 3) (Fig. 4F). The model showed balanced accuracy of 73.25% on the held-out test dataset with a sensitivity of 70.1% and specificity of 76.4% (<u>Table S9</u>). We also observed that the prediction score obtained from the LogReg model did not demonstrate any association with the clinical groups, high-risk HLA types (DR3 and/or DR4), or islet autoantibody status (Fig. S12). Contrary to common observation, LogReg scores exhibited a positive correlation (r = 0.12) with T1D duration. Whereas, patient age had negative correlation with LogReg score across all clinical groups (r = -0.45 to -0.5) except for T1D (r = 0.15). It is important to note that we observed overfitting on the training dataset even after regularising the coefficients (see **Methods**). We subsequently employed a more sophisticated DL model to enhance the classification accuracy of immune repertoires.

#### Interpretable DL-based multiple instance learning achieves comparable performance to HLA-associated TCR features in differentiating clinical groups

Significant developments have been made in applying DL methods for the classification of immune repertoires based on disease status (M. Chen et al., 2023; Emerson et al., 2017; Katayama & Kobayashi, 2022; Widrich et al., 2020). The Deep Repertoire Classification (DeepRC) model is one such modular and customizable method particularly suited for large-scale multiple-instance learning problems including immune repertoire classification (Widrich et al., 2020) (Fig. 4G). Similar to the LogReg approach, the DeepRC model was also trained using a 5-fold CV, where three splits were designated for training, one for tuning, and the remaining one for testing, in a recursive manner. The best-performing model for each CV fold was selected based on the AUROC of the tuning set, resulting in five selected models. The AUROC of these five best-performing models ranged from 0.69 to 0.76 (<u>Table S9</u>) on the remaining testing set in cohort 1. We further trained an ensemble LogReg model to combine the predictions of the 5 best models, which improved the AUROC to 0.79 (balanced accuracy 72.5%). The ensemble model was considered the final DeepRC model and applied to the held-out test set, resulting in the AUROC 0.77 (balanced accuracy 72.9%) (Fig. 4H).

The DeepRC model exhibited performance comparable to the LogReg model on the held-out test set and observed association with clinical groups, high-risk HLA types (DR3/DR4), autoantibody status, and age in cohort 1 (Extended Data Fig. 8). Notably, DeepRC predictions based solely on the TCR repertoire demonstrated improved classification performance compared to HLA-associated TCR features (such as CDR3 risk score, pHLA- and nHLA-motif scores). However, its performance was lower when compared to HLA-based genetic risk (HLA risk score). Interestingly, DeepRC’s predictions also showed a positive correlation with T1D duration, similar to the k-mer-based LogReg model, while exhibiting a negative correlation with the age of T1D patients. Additionally, DeepRC allowed for the extraction of a simplified sequence motif representation from the trained model, facilitating a biologically meaningful interpretation of the DL model, as discussed in the section below.

#### T1D-associated CDR3β motifs simplify the DL model with minimal performance tradeoff while maintaining key trends

Interpretability of the above presented DL model is a challenging task; however, DeepRC (Widrich et al., 2020) supports different methods of interpretability, via the attention values and contribution analysis method known as Integrated Gradients (Sundararajan et al., 2017). We extracted a low-complexity DeepRC-motif to identify a biological signature of T1D (see **Methods**) and calculated the DeepRC-motif score by normalizing the number of motif-containing sequences with the total number of CDR3β sequences. The DeepRC-motif achieved an AUROC of around 0.7 on both training dataset (balanced accuracy 65.67%) and held-out test dataset (balanced accuracy 66.48%) (<u>Table S9</u>). The DeepRC-motif showed association with clinical groups, high-risk HLA types (DR3 and/or DR4), autoantibody status, and age in cohort 1, similar to the DeepRC model. However, T1D duration showed a negative correlation (r = -0.13) with the DeepRC-motif score (Extended Data Fig. 9). Taken together, DeepRC provided a low-complexity alternative in the form of a motif, which demonstrated an association with clinical groups, despite being derived solely from TCR repertoires and without HLA information. It is noteworthy that the DeepRC-motif is a gapped motif with two optional positions out of total five positions, which may exhibit a higher rate of false positives in identifying T1D-associated CDR3β sequences, as ∼17% of the CDR3β sequences in a repertoire contained a DeepRC motif, compared to only ∼0.4% for the pHLA motif.

The generation of pHLA-motifs by aggregating position-specific AAs from CDR3β sequences of varying lengths presents considerable challenges. Nevertheless, only ∼0.4% of the repertoires contained these motifs. To enhance specificity, we obtained refined consensus HLA-associated CDR3β sequences by identifying the CDR3β sequences that contain both pHLA- and DeepRC-motif, termed “consensus-motif”. This approach reduced the proportion of HLA-associated CDR3β sequences to approximately 0.25%, without reducing the performance compared to pHLA-motifs. Additionally, AUROC of consensus-motif (0.74) was marginally improved compared to pHLA-motif (0.73) (Fig. S13). Expectedly, the consensus-motif score was higher in T1D donors and increased alongside the number of autoantibodies and risk HLA (Fig. S13). It also showed negative correlation with both patient age and T1D duration (Fig. S13).

### T1D-associated TCR motifs as predictive markers in independent cohorts

#### T1D genetic risk loci are associated with TCR motifs in peripheral blood and pancreatic lymph nodes

Despite significant progress in defining genetic loci that contribute to T1D risk via Genome-wide association studies (GWAS), our understanding of the impact for such variants on immune function remains limited. We have shown that high-risk HLA alleles restrict the TCR repertoires, allowing for potentially autoreactive TCR presence in genetically-predisposed individuals. Thus, we hypothesized that T1D risk variants, particularly in the HLA region, may be associated with increased frequency of the pHLA-motif and DeepRC-motif. To address this, we performed microarray-based precision medicine genotyping (Perry et al., 2023) of 716 unrelated living participants from cohort 1 (ND, n=489; T1D, n=227).

To understand which T1D risk variants contributed to the association with the enriched TCRβ motifs, we performed quantitative trait locus (QTL) analysis. *HLA-DQA1*\*05:01-*DQB1*\*02:01 (p=3.91e-10), *HLA-DRB1*\*0301 (p=5.83e-10), *HLA-DQA1*\*01:02-*DQB1*\*06:02 (p=3.47e-09), *HLA-DRB1*\*15:01 (p=2.38e-07), *HLA-DQA1*\*03:0X-*DQB1*\*03:01 (p=8.5e-05), and *HLA-DQA1*\*05:05-*DQB1*\*03:01 (p=3.23e-4) haplotypes were significantly associated with DeepRC-motif frequency independent of disease status such that risk haplotype correlated with an increased motif score (Fig. 5A, B). The risk alleles of a T1D- associated variant tagging the *XL9* super enhancer (p=3.91e-10), known to regulate *HLA-DRB1* and *HLA- DQA1* expression (Majumder et al., 2020), and an intergenic *HLA-DRA1-DRB1* variant (p=1.85e-3) were likewise associated with increased peripheral blood DeepRC-motif score (Fig. 5A, B). An intergenic deletion nearby *CTLA4* carrying risk for T1D that is in linkage with reduced *CTLA4* expression quantitative trait loci (eQTL) (Machiela & Chanock, 2015; Võsa et al., 2021) was also weakly associated with increased DeepRC-motif score (p=0.015) (Fig. 5A, B). Analysis of the pHLA-motif revealed enrichment in risk allele-carrying subjects for *HLA-DRB1*\*0301 (p=0.018), *HLA-DQA1*\*05:01-*DQB1*\*02:01 (p=0.018), and *HLA-DQA1*\*05:05-*DQB1*\*03:01 (p=0.021) (Extended Data Fig. 10A, B) similar to that observed for the DeepRC-motif. The nHLA-motif frequency was decreased in those with the T1D risk haplotypes *HLA- DQA1*\*02:01-*DQB1*\*02:02 (p=9.14e-08), *HLA-DQA1*\*03:0X-*DQB1*\*03:02 (p=9.85e-06), and HLA-DR4 (p=1.1e-05) (Fig. S14A, B). Thus, variants affecting HLA class II type in addition to HLA class II and *CTLA4* expression levels may influence the frequency of T1D-enriched TCRβ motifs in blood.

**Fig. 5.**
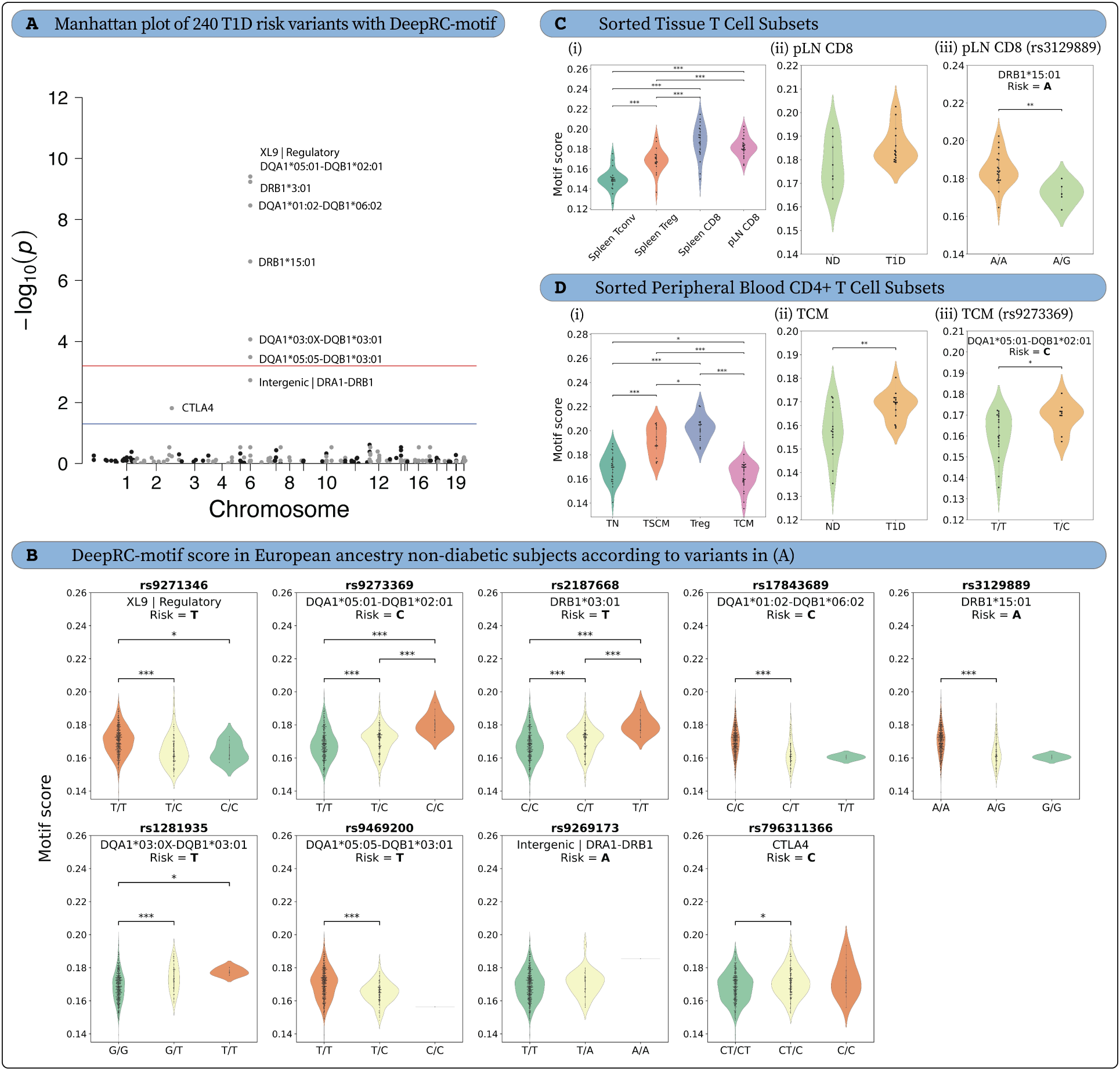
DeepRC motif frequency enriched in carriers of T1D risk genetics in bulk and sorted central memory CD4^+^ T cells from peripheral blood as well as pancreatic lymph node CD8^+^ T cells. **(A)** Manhattan plot of 240 T1D risk variants versus DeepRC-motif score in bulk peripheral blood. Linear regression assuming additive genotypic effect with age, sex, T1D status, predicted probability of CMV infection, and 10 multidimensional scaling components as covariates. Benjamini-Hochberg false discovery rate threshold at p=5.73e-4 (red line) and a conventional threshold of p = 0.05 (blue line). **(B)** Violin plots of DeepRC-motif score in European ancestry non-diabetic (ND) subjects, according to variants in **(A)**. Gene or tagged HLA genotype and risk allele annotated above each plot. **(C)** DeepRC-motif score across sorted T cell subsets in spleen and pancreatic lymph node (pLN). **(i)** Mixed-effects analysis with Tukey’s multiple comparisons test. **(ii)** pLN CD8^+^ T cell DeepRC-motif score according to T1D status. **(iii)** pLN CD8^+^ T cell DeepRC-motif score according to DRB1*15:01 tag SNP. Linear regression with age, sex, diabetes status, and 10 multidimensional scaling components as covariates. **(D)** DeepRC-motif score in sorted peripheral blood CD4^+^ T cell subsets. **(i)** Mixed-effects analysis with Tukey’s multiple comparisons test. **(ii)** Central memory CD4^+^ T cell DeepRC-motif score according to T1D status. **(iii)** Central memory CD4^+^ T cell DeepRC-motif score according to DQA1*05:01-DQB1*02:01 tag SNP. Linear regression with age, sex, and T1D status as covariates. In all violin plots, p-values for pairwise testing were calculated using two tailed Mann-Whitney U tests and p-values were adjusted between different clinical groups using the Benjamini–Hochberg method. p-values were described as * for [0.01,0.05], ** for [0.001,0.01] and *** for <0.001 and no stars plotted for non-significant values. **Supplementary Figures:** Extended Data Fig. 10, Fig. S14

Observations in peripheral blood were tested in TCRβ-sequenced sorted conventional CD4^+^CD127^+^ T cells (Tconv), CD4^+^CD127^−^CD25^+^ regulatory T cells (Treg), and CD8^+^ T cells from pancreatic lymph nodes (pLN) as well as spleen from 27 Network for Pancreatic Organ donors with Diabetes (nPOD) donors (data previously published (Seay et al., 2016): ND, n=7; T1D, n=15; T2D, n=3; other diabetes, n=2) in order to understand matters of cell type and tissue specificity. DeepRC- and pHLA-motif scores were significantly increased in CD8^+^ T cells in the spleen and pLN as compared to Tconv and Treg in the spleen (Fig. 5C, Extended Data Fig. 10C). Treg DeepRC- and pHLA-motif scores were also higher than those of Tconv in the spleen (Fig. 5C, Extended Data Fig. 10C). nHLA-motif score was enriched in splenic Tconv and Treg as compared to spleen and pLN CD8^+^ T cells (Fig. S14C). T1D subjects showed a trend for increased DeepRC (p=0.095, Fig. 5C) and pHLA (p=0.067, Extended Data Fig. 10C) motif scores and increased nHLA (p=0.025, Fig. S14C) motif score in pLN CD8^+^ T cells as compared to those with ND. Of the peripheral blood findings, *HLA-DRB1*\*15:01 (p=0.004) was also significantly associated with decreased DeepRC TCR motif frequency in pLN CD8^+^ T cells. nHLA motif score was decreased in *HLA- DQA1*\*02:01-*DQB1*\*02:02 pLN CD8^+^ T cells (p=0.226, Fig. S14C), in contrast to the increased frequency in peripheral blood (Fig. S14A). These data support the notion that HLA class II-mediated protection from T1D may limit the frequency of potentially autoreactive TCRβ motifs at the site of autoimmunity.

Similar analysis was performed in TCRβ-sequenced sorted CD45RO^-^CD27^+^CCR7^+^CD95^-^ naive (TN), CD45RO^-^CD27^+^CCR7^+^CD95^+^ stem cell memory (TSCM), CD25^hi^CD127^lo/–^Treg, and CD45RO^+^CD27^+^ central memory (TCM) CD4^+^ T cell subsets from peripheral blood of 28 living study participants (data previously published (Gomez-Tourino et al., 2017): ND, n=14; T1D, n=14). DeepRC- and pHLA-motif scores were significantly higher in TSCM and Treg than TN and TCM (Fig. 5D, Extended Data Fig. 10D). nHLA motif score was increased in Treg as compared to TN and TCM, while TSCM also showed higher frequency than TCM (Fig. S14D). Despite showing lower DeepRC-motif score compared to other cell subsets, TCM DeepRC-motif score was significantly enriched in T1D as compared to ND subjects (p=0.0047, Fig. 5D), which can partially be accounted for by increased motif score in *HLA-DQA1*\*05:01- *DQB1*\*02:01-carrying subjects (p=0.028, Fig. 5D). *HLA-DQA1*\*05:01-*DQB1*\*02:01 was also associated with enrichment of the pHLA-motif score in Treg (p=0.031) and TCM (p=0.027) (Extended Data Fig. 10D). Together, these data suggest that T1D HLA risk genetics may confer selection pressure for T1D-enriched TCRβ motifs in peripheral blood, particularly in CD4^+^ TCM and Treg.

## Discussion

The adaptive immune repertoire is shaped by a myriad of factors, including aging, environmental exposures, and genetics. T1D natural history studies identified islet autoantibody seropositivity as early autoimmune signals in genetically predisposed populations (Achenbach et al., 2005; Atkinson & Eisenbarth, 2001). Yet there is a need to improve disease prediction and monitoring through the validation of additional cellular biomarkers. It has been proposed that the TCR repertoire, as a record of response to foreign and self-antigens, could potentially distinguish a T1D-specific signal from repertoire shifts caused by pathogen exposures and vaccinations. In this study, we have comprehensively assessed the HLA-based TCR repertoire restriction, TCR repertoire classification, and immune signal identification on a cross-sectional dataset of 2250 TCR repertoires spanning various stages through the natural history of T1D. We initially observed that repertoire-level diversity, similarity, and V-gene usage do not differentiate T1D donors from their relatives without diabetes or unrelated controls. However, mapping known T1D antigen-specific clones (Tickotsky et al., 2017) in our dataset revealed an enrichment of T1D-associated sequences in T1D repertoires. This mirrors recent data that demonstrated stable enrichment of CDR3β sequences matching known preproinsulin reactive clones during disease progression in a genetically at-risk cohort, thus providing support for T1D reactive sequences as a biomarker in T1D (Mitchell et al., 2022). Despite this, we also found T1D-associated sequences frequent in non-T1D donors. This is in accordance with previous reports suggesting phenotypic rather than frequency differences in circulating autoreactive cells in T1D (Skowera et al., 2015), and may also be impacted by an incomplete understanding of antigen reactivities and receptor sequences in T1D, altogether complicating the utility of *a priori* sequences and necessitating a more comprehensive analyses of our bulk repertoire data.

We next examined how high-risk HLA molecules, which account for most of the genetic risk in T1D (Hu et al., 2015; Noble & Valdes, 2011), influence T1D-associated TCR features. Interestingly, clustering of public TCRs yielded more significantly underrepresented hits associated with HLA risk allele *HLA-DRB1*\*0301 in T1D, while we identified overrepresented TCR features associated with risk HLA-DQ molecules *DQA1*\*0501 and *DQB1*\*0201 in T1D. The observation of a majority of significant underrepresented features is intriguing given previous data suggesting persistent low grade viral infections are associated with autoimmunity development (Krogvold et al., 2015), or that certain infections may be protective (Ekman et al., 2019; Strom, 2009). Alternatively, our observations could reflect deficits in regulatory subsets in T1D; however, while differences in diversity have been shown for other circulating T cell subsets in T1D, this is not the case for Tregs (Gomez-Tourino et al., 2017). Thus, further investigation of the identity and phenotypes of T cells expressing these differentially abundant sequences is warranted.

We further analyzed the HLA-DQβ1 and HLA-DRβ1 variants associated with high risk for T1D (Hu et al., 2015) to examine their effect on TCR repertoire restriction (Ishigaki et al., 2022). We identified several TCR features, including a repertoire-level CDR3 risk score and subsequence-level CDR3 phenotypes, which indicate HLA-based T1D-susceptibility imparted solely by TCR repertoires. Additionally, we identified HLA-associated CDR3β motifs by aggregating CDR3 phenotypes, which were positively (pHLA) and negatively (nHLA) associated with T1D. Importantly pHLA-motifs were enriched in T1D donors and exhibited an increased frequency of aromatic, negatively charged and hydrophobic residues, particularly in the CDR3β middle and C terminal region, a feature of self-reactivity (García et al., 2022; Lagattuta et al., 2022; Textor et al., 2023). Interestingly, pHLA-motif scores exhibited a negative correlation with disease duration and patient age, potentially indicative of a waning autoimmune response associated with concomitant loss of 𝛽 cell antigens (C. L. Williams et al., 2022). To test the potential for HLA alleles to influence the diversity of the TCR repertoire, we modeled T cell selection by calculating the probability of generation and selection of TCR sequences. In accordance with a recent murine study (Brown et al., 2024), we observed, although weakly, that heterozygosity limits repertoire diversity. This runs contrary to some observations that the highest risk HLA DQ2/DQ8 genotype allows for transdimer formation, thus potentially allowing for presentation of a more diverse pool of antigens (van Lummel et al., 2012) or permitting cross-reactivity (Chow et al., 2019). Moreover, recent work suggests that HLA-DM mediated editing is reduced for HLA-DQ2 and HLA-DQ8 pMHC molecules (Zhou et al., 2016). While it is thought that lack of DM editing would favor presentation of peptides with more rapid dissociation rates (D. A. Weber et al., 1996) and thus, allow for broader peptide presentation, it is still unclear how lower versus higher stability of pMHC complexes affect the diversity of the repertoire (Denzin, 2013). These analyses exemplify the multifaceted influence of HLA and the complexity of immune regulation in T1D, while providing further insights into the interplay between genetic risk and TCR repertoire diversity.

Considering that most prior analyses have focused on HLA as a primary determinant of T1D risk, we applied several ML strategies to identify features indicative of T1D status independent of HLA information. Although the performance of HLA-dependent TCR features (pHLA and nHLA motifs; AUROC: ∼0.73) was lower than the HLA risk score (AUROC: 0.85), the classification of T1D status without explicitly utilizing HLA allele information achieved notable performance with the K-mer-based LogReg strategy (AUROC: 0.78) and DeepRC (AUROC: 0.77). While the K-mer-based LogReg model exhibited overfitting on the training dataset, the DeepRC model proved more interpretable and capable of identifying the input sequence with the highest contribution to the T1D prediction per repertoire. These identified sequences were subsequently analyzed to derive a T1D-associated motif. We found motif scores to be increased in T1D donors and to be increased alongside the number of islet autoantibodies and risk HLA, altogether providing support for this receptor motif as being involved in T1D pathogenesis. Similar to our pHLA- motif, we once again found a negative association of DeepRC-motif score with T1D duration. A caveat for our analysis is the potential for false positives and non-specific sequences included in our ML motif. The frequency of CDR3β sequences containing the DeepRC motif was approximately 17%, whereas the estimated precursor frequency of T1D antigen specific T cells is around 1-10 per million cells (Naik et al., 2004), closer to the frequency of sequences possessing the pHLA-motif. It is possible that the high frequency of DeepRC-motif-containing sequences could be reflective of incorporation of sequences that are not restricted by high-risk HLA alleles, cross-reactivity (Tran et al., 2024), or bystander activation, any of which could be important in T1D progression. Thus, further investigation is needed to examine the epitope reactivity of sequences containing this motif. Moreover, we expect incorporation of additional information such as paired TCRαβ sequences and cell phenotype features (e.g., activated effector memory states) to further improve the classification accuracy.

We found T1D risk alleles, primarily within the HLA region, to control the frequency of the DeepRC-motif. Notably, risk genotypes at *HLA-DQ* and *HLA-DR* loci, as well as regulatory element *XL9*, were associated with a higher motif score. We recently reported that high-risk HLA alleles contributed to increased HLA expression on circulating monocytes (Shapiro et al., 2023). Our motif score eQTL (Võsa et al., 2021) results provide further support for the notion that enhanced HLA class II expression modulated by risk alleles could promote the activation and expansion of lower affinity autoreactive clones in T1D. Interestingly, the only eQTL hit outside of the HLA region passing the significance threshold was located within *CTLA4*, and was previously linked with reduced CTLA4 expression. Notably, other T1D risk-conferring variants within *CTLA4* are known to negatively impact *CTLA4* expression and function (Robertson et al., 2021) and thus, are thought to result in reduced TCR activation threshold or impaired regulatory signaling.

HLA-DQ restricted proinsulin and HLA-DR restricted GAD-specific T cells have been identified within the insulitic lesion (Pathiraja et al., 2015) or peripheral blood (Eugster et al., 2015) as well as the pLN and spleen of T1D donors (Seay et al., 2016). To explore the tissue relevance of the identified DeepRC-motif and the cell type localization of motif bearing sequences, we interrogated the motif in our previously published sorted bulk-sequenced nPOD tissue dataset (Seay et al., 2016). We found that motif bearing sequences were increased in CD8^+^ T cells and Tregs in the spleen and pLN, with increased motif score in pLN CD8^+^ T cells of T1D donors, which was modulated by genotype at protective HLA class II loci. The observation of HLA class II loci modulating CD8^+^ T cell phenotype likely indicates an indirect effect whereby risk alleles promote inflammatory CD4^+^ T cell phenotype or alter Treg phenotype to permit this enrichment (Stadinski et al., 2023), though the mechanisms controlling this need further defining. Accordingly, in a separate validation cohort (Gomez-Tourino et al., 2017) consisting of a sorted CD4^+^ T cell subset from the PBMC of T1D and CTRL individuals, we identified an enrichment in motif score among CD4^+^ TCM cells in T1D donors, which was impacted by HLA-DQ risk genotype. CD4^+^ TCM cells have the capacity to produce cytokines such as IL-2 and IL-21, which drive CD8^+^ T cell proliferation and effector phenotype (Zander et al., 2022). Notably, alterations in the peripheral blood CD4^+^ TCM compartment, specifically involving increases in T follicular helper (TFH) cells, have been reported in T1D and prior to overt disease (Bettelli & Campbell, 2020; Ferreira et al., 2015). In fact, an altered TFH-like signature is detectable as early as infancy in genetically at-risk children who later progressed to T1D, implicating genetics as a driving factor in enhancing helper and effector T cell function in T1D. Thus, we have identified a replicable TCRβ motif which encodes T1D status, is relevant in circulation as well as disease relevant tissue and is controlled by T1D risk loci.

One key aspect of our study is that HLA-based TCR restriction analysis did not account for clinical group information, while FDR and SDR repertoires were classified as ND in the ML models. Nevertheless, most analyses, including the DeepRC model, HLA and CDR3 risk scores, and DeepRC- and pHLA-motif scores, revealed a trend where the scores were highest for T1D repertoires and lowest for CTRL, following the expected pattern: T1D > FDR > CTRL. Furthermore, both the DeepRC-motif and pHLA-motif were overrepresented in individuals with genetic risk for T1D across several independent cohorts, including bulk and sorted CD4^+^ TCM cells from peripheral blood as well as pLN CD8^+^ T cells. Notably, we observed a limited presence of public TCRs in our dataset compared to previous studies focused on other autoimmune diseases (Emerson et al., 2017; Mitchell et al., 2022; Mitchell & Michels, 2020). This discrepancy may be attributed to high variability in confounding factors, including age, T1D duration, genetic background, and ethnicity of the individuals. A large longitudinal HLA-stratified cohort could potentially enhance the identification of T1D-specific public TCRs. In this study, we integrated the assessments: (i) the influence of genetic risk on TCR repertoires with HLA-based restriction analysis, and (ii) influences of T1D- associated genetic and environmental factors (García et al., 2019) on TCR repertoires through ML-based methodologies. This integration underscores the critical role of HLA in T1D risk and highlights adaptive immune motifs associated with T1D risk.

### Limitations of this study

As with all AIRR studies, sequencing depth remains a critical concern, particularly for autoimmune diseases such as T1D, where immune signals in peripheral blood may be relatively low (Christophersen et al., 2014; C. R. Weber et al., 2022). The detection of an immune signature is further hindered by limited information on cell populations and antigen specificity. While the repertoires in the current study maintain an average of unique CDR3β sequence count > 100,000 (cohorts 1 and 2), deep sequencing in cohort 3 yielded nearly a 3.5-fold increase in unique CDR3β sequences. Although analysis of replicates demonstrated a high MH index and strong Pearson correlation between deep and shallow sequencing data, the loss of additional unique CDR3β sequences at lower sequencing depth may impact the identification of rare, T1D-specific signals. Furthermore, repetition in data collection centers introduced duplicate repertoires with different donor identification numbers into the dataset. While we applied an MH similarity threshold to exclude duplicates, there remains a possibility that some duplicate repertoires were retained in the dataset.

Furthermore, repetition in data collection centers resulted in the inclusion of duplicate repertoires associated with different donor identification numbers in the dataset. To address this, we applied an MH index similarity threshold of 0.4 to exclude duplicates. Nevertheless, there remains a possibility that some duplicate repertoires were not excluded from the dataset.

The HLA risk score is derived from three HLA positions with the strongest association with T1D risk. A more comprehensive HLA risk score incorporating additional HLA positions could improve T1D risk prediction. HLA risk score is fixed and thus, cannot serve as a biomarker for T1D disease progression, diagnosis, or assessing the effects of T1D treatment.

Islet autoantibody seroconversion peaks early in childhood, at around two years of age (Ziegler et al., 2012). As this represents a critical period of exposure to pathogens and vaccinations, it will be extremely important to understand the longitudinal stability of the TCR repertoires in children to effectively identify TCR-based signatures and monitor their shifts over time in diseases such as T1D. A more extensive longitudinal sampling could also provide insights into how other diseases or infections influence the TCR repertoire at specific time points. Additionally, a larger cohort of HLA-matched individuals would be ideal for identifying HLA-specific signals associated with T1D.

It is also essential to acknowledge the limitations associated with derivation of HLA-motifs. HLA-motifs were constructed by aggregating position-specific AA of different CDR3β lengths, which complicates the interpretation of positional dependence. Furthermore, motifs derived from aggregated positional information do not permit the retrieval of AA-specific positional dependencies and are primarily suited to produce ungapped motifs. The DeepRC-derived motifs had unexpectedly higher proportions within each repertoire, indicating a substantial rate of false positive CDR3β sequences and emphasizing the need for more effective motif extraction methods. A comprehensive study is also warranted to examine the role of underrepresented protective motifs within the T1D cohort, potentially enhancing T1D risk prediction.

A future strategy toward identifying disease-relevant biomarkers may benefit from combining PBMC-based sorting with tissue-based, spatially resolved, and cell-population-specific sampling. That being said, we showed that the T1D-associated TCR signals identified were also present in entirely different datasets (Culina et al., 2018; Mhanna et al., 2021), tracking with HLA-risk status. Additionally, T1D antigen-specific TCRs were also observed in CTRL individuals, warranting further studies on low-affinity, cross-reactive signals and physiological autoreactive processes (Eugster et al., 2024). Biomarker specificity could be further enhanced by incorporating paired alpha and beta chain information (Pogorelyy et al., 2024). Identifying epitopes reactive with TCRs exhibiting overrepresentation or depletion in T1D repertoires could provide valuable insights into underlying disease mechanisms and help refine associated motifs. To circumvent large-scale experimental screening for antigen specificity (which is currently unfeasible), there is a need for AI-based deorphanization of TCR epitopes (Joglekar & Li, 2021; Ma et al., 2021; T et al., 2024; A. Weber et al., 2024; Zdinak et al., 2024).

## Conclusions

There is growing interest in harnessing the immune history encoded within TCR repertoires to develop TCR-based diagnostics and therapeutics for future clinical applications, including immunization (Glanville et al., 2017; Minervina et al., 2019), viral infections (Emerson et al., 2017; Shomuradova et al., 2020), and autoimmune diseases (Jacobsen et al., 2017; Nakayama & Michels, 2021). We conducted a large-scale, comprehensive analysis of T1D-associated TCR repertoire alterations by immunosequencing the CDR3β regions and genotyping HLA using sampling amenable to young donor samples for screening efforts. The HLA-associated T1D risk was leveraged to extract robust HLA-mediated TCR features, including repertoire-level CDR3 risk score and CDR3β subsequence-level HLA-motifs, to predict T1D risk. Additionally, we predicted T1D status directly from TCR repertoires using ML and DL approaches. DeepRC performed comparably to HLA-mediated TCR features and identified a simplified DeepRC-motif from TCR repertoires alone, without needing HLA information. These potential motif-based biomarkers, derived from cross-sectional samples, reflect the natural history and pathogenesis of T1D, and show associations with HLA, disease duration, and islet autoantibody status. These findings not only support future longitudinal studies but also, introduce disease-associated motifs that can be tracked in response to clinical interventions aimed at halting disease progression, such as T cell-targeting therapies like teplizumab (anti-CD3, Tzield), anti-thymocyte globulin (ATG), Alefacept (LFA3-Ig fusion protein), and Abatacept (CTLA4-Ig fusion protein) (Gitelman & Bluestone, 2016). As studies expand in organ donor tissues from individuals with T1D, this work is expected to provide a valuable reference dataset for identifying antigen reactivities and convergent or public receptor sequences enriched in the pLN and pancreas.

## Methods

### Study participants and sample collection

Study participants or their legal guardians provided written informed consent with pediatric and adolescent participants also providing assent prior to enrollment. Cohort 1 was comprised of individuals from outpatient clinics at the University of Florida (UF; Gainesville, FL), Nemours Children’s Hospital (Orlando, FL), and Emory University in accordance with Institutional Review Board (IRB) approved protocols at each site. Peripheral blood samples were collected into sodium heparin-coated and serum separator vacutainer tubes by venipuncture from non-fasted individuals (i.e., unknown prandial state or time of day), then shipped or rested overnight prior to processing at the UF Diabetes Institute (UFDI). At the time of blood draw, participants were generally healthy with no reported malignancy or infection, and sample collection occurred between 2010-2018 prior to the coronavirus disease 2019 (COVID-19) pandemic. All samples and their associated data and metadata were deidentified in accordance with UF IRB201400703. Cohort 2 included subjects with T1D from the T1D Exchange Clinical Network (Beck et al., 2012; Davis et al., 2015) and cohort 3 included controls sequenced by Adaptive Biotechnologies.

### DNA isolation and TCRβ sequencing

Genomic DNA (gDNA) was isolated from peripheral blood mononuclear cells (PBMCs) of 2250 subjects. *TRB* (TCRβ) CDR3 region sequencing was performed via Adaptive Biotechnologies immunosequencing assay. Briefly, gDNA was amplified using bias-controlled multiplex PCR prior to sequencing (Carlson et al., 2013; Robins et al., 2009). Cohort 1, containing 1393 repertoires, comprising 188 CTRL, 59 SDR, 625 FDR, 426 T1D repertoires, was sequenced using hsTCRB_v4 Service at shallow sequencing depth (Carlson et al., 2013; Robins et al., 2009). There were 95 additional autoantibody positive non-T1D repertoires (AAb+) in cohort 1, which were excluded to avoid potential bias in the study. Cohort 2, containing 679 T1D repertoires, was sequenced using hsTCRB_v4b Service at shallow depth. Cohort 3, containing 178 CTRL repertoires, was sequenced using hsTCRB_v4b Service at deep sequencing depth. The shallow and deep technical replicates of cohort 1 were studied separately and were not part of the above mentioned cohort.

### Genotyping

Subjects from cohorts 1 (n=1242), 2 (n=645), and 3 (n=178) were genotyped using the UFDIchip custom microarray, with processing on an Affymetrix GeneTitan instrument and a BioMek FX dual arm robotic workstation (Perry et al., 2023). The UFDIchip includes >9,000 markers from the Axiom^TM^ Precision Medicine Research Array (Thermo Fisher Scientific) covering the HLA region. Raw data were converted to genotype calls using Axiom™ Analysis Suite software (v3.0, Thermo Fisher Scientific) “Best Practices Workflow” with “Human.legacy.v5” settings. Four-digit HLA genotypes were imputed using Axiom^TM^ HLA Analysis software (v1.2.0.38) (Dilthey et al., 2013). Imputation results were used for analysis if probability scores > 0.7. HLA class II haplotypes considered to carry T1D risk were defined as DR3 *(HLA-DRB1*\*03:01–*HLA-DQA1*\*05:01–*HLA-DQB1*\*02:01) and DR4 (*HLA-DRB1*\*04:01/02/04/05/08–*HLA-DQA1*\*03:01–*HLA-DQB1*\*03:02/04) (Noble & Valdes, 2011). Subjects were grouped based on HLA class II risk diplotypes, with non-risk haplotypes designated as “DRX”, resulting in the following groups: DRX/X, DR3/X, DR3/3, DR4/X, DR4/4, or DR3/4. SNP2HLA (v1.0.3) was used for HLA amino acid polymorphism imputation from genotyping data (Jia et al., 2013).

### Islet autoantibody measurement

GADA, IA-2A, and ZnT8A were measured from serum using commercial ELISA kits, which consistently demonstrate high sensitivity and specificity in the Islet Autoantibody Standardization Program (IASP) workshops (Wasserfall et al., 2016). IAA was measured from serum by chemiluminescence assay, as recently reported and evaluated by the IASP (Marzinotto et al., 2023).

### Dataset preprocessing

Firstly, we filtered the Out-of-frame sequences from each repertoire and selected repertoire, which contains a minimum of 50,000 distinct sequences. In this study, we defined a clone or clonotype as a unique CDR3β sequence, unless specified otherwise. The clone counts for duplicate CDR3β amino acid sequences were merged. Additionally, V/D/J-gene information from different columns was consolidated to address unresolved V/D/J-gene annotations. Repertoires with a potential common donor origin were excluded by applying a Morisita-Horn (MH) similarity threshold of less than 0.40, based on thresholds established in previous studies on replicate repertoires (Chang et al., 2020; Niu et al., 2015).

### Statistical analysis

All cluster heatmaps were generated using the UPGMA clustering method and Euclidean distance matrix (Sokal & Michener, 1958). The p-values for the multiple testing were performed using Kruskal–Wallis test. Similarly, p-values for pairwise testing were calculated using two tailed Mann-Whitney U tests. p-values were described as * for [0.01,0.05], ** for [0.001,0.01] and *** for <0.001 and no stars plotted for non-significant values. All p-values were adjusted for multiple testing using the Benjamini–Hochberg method.

### Repertoire-level similarity and diversity analysis

#### V-gene usage analysis

The V-gene distribution within each immune repertoire was determined by assessing the frequency of occurrence for each V-gene. This frequency was obtained by dividing the count of a specific V-gene by the total number of unique clones present in the repertoire. The V-gene frequencies were calculated without considering the clonal frequency.

#### Hill-based diversity profile analysis

The diversity of the TCR repertoires were calculated using Hill-based diversity profile, which is based on Rényi’s definition of generalized entropy (Hill, 1973; Jost, 2006; Rényi, 1961). It is defined as:

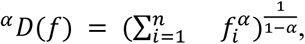

Where 𝑓 is the clonal frequency distribution with 𝑓_𝑖_ being the frequency of each clone and 𝑛 the total number of clones. The 𝛼-values represent weights, which means as 𝛼 increases, higher frequency clones are weighted more. The 𝛼-parameterized Diversity generates a diversity index profile for a given array of 𝛼 values. The diversity profile is not defined for 𝛼 = 1. However, diversity tends towards Shannon entropy when 𝛼 tends to 1 (based on L’Hospital’s rule). We calculated the diversity profile of each repertoire based on 𝛼 value ranging [1,10] with step size of 0.2. The above analyses were performed on individuals with age less than 30 in cohort 1 to avoid confounding factors due to age (Britanova et al., 2014).

#### Public clone analysis

In order to evaluate the similarity between TCR repertoires, we defined public clones as identical CDR3β sequences shared among two repertoires. The percentage of shared public clones is defined as:

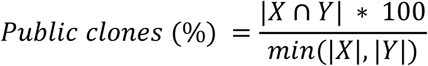

Where, |𝑋| and |𝑌| are the repertoire sizes (number of unique clones) of the repertoire X and Y. The number of shared clones between both repertoires are |𝑋 ∩ 𝑌|.

#### Morisita-Horn similarity index analysis

We employed the MH similarity index (Horn, 1966) to assess the degree of similarity among TCR repertoires, taking into account the clonal frequency attributed to each unique clonotype. The MH index between a pair of repertoires is defined as:

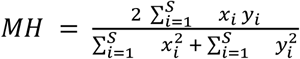

Where, S is the number of unique clones, and x and y denote the frequency of i^th^ clone in either repertoire. The MH index ranges between 0 (no overlap) and 1 (complete clonal overlap and identical clonal frequencies).

### Overrepresentation analysis of different disease-associated CDR3β sequences

We downloaded the McPAS (Tickotsky et al., 2017) and VDJdb (Bagaev et al., 2020) datasets from the respective websites (April, 2024). The unavailable, redundant and non-human sequences were removed from the dataset. Only pathology (in McPAS)/antigen species (in VDJdb) with more than 30 sequences were selected for analysis. The final statistics of the datasets is given in <u>Table S1</u>. We obtained the overlapping CDR3β sequences between cohort 1 and McPAS/VDJdb datasets using CompAIRR (Rognes et al., 2022). We considered the clonal frequency of the overlapping sequences in each repertoire in cohort 1 and normalized the count using the following formula and grouped them by the clinical groups.

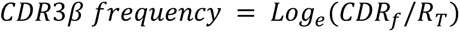

Where, 𝐶𝐷𝑅_𝑓_ is the clonal frequency of the overlapping CDR3β clone in a repertoire in cohort 1 and 𝑅_𝑇_ is the sum of clonal frequencies in the same repertoire.

### HLA-conditional T1D-association testing in single AA variation neighborhood

We first considered each unique TCRβ sequence within T1D positive repertoires from Cohort 2. The sequences were partitioned by the IMGT V-gene family and all receptor sequences from 669 repertoires were concatenated to map each sequence to its exact and near-exact (single AA variation distance) neighbors. To avoid O(n^2^) comparisons on such a large set of sequences, we identified nearly identical sequences using a fuzzy clustering technique. This method bins highly similar sequences into shared memory locations using keys with a single masked position, inspired by prior work on TCR clustering (Chotisorayuth & Tiffeau-Mayer, 2024; Glanville et al., 2017; S. Liu et al., 2023b; Rognes et al., 2022; Valkiers et al., 2021). Briefly, each CDR3 was represented as the set of possible sequences with a single masked wildcard position for either a substitution or insertion in a Python dictionary. Each unique key was then assigned an integer index and two arrays link CDR sequence indices to masked CDR3 key indices. Group-by key and permutation operations were then applied for rapid identification of all links between identical and near-identical CDR3 sequences. After identifying single AA variation sequence neighbors of each unique CDR3 centroid, only CDR3s with at least five total neighbors in Cohort 2 were retained for further analysis. Next, we considered the donors associated with each CDR3 sequence index and computed the odds ratio (OR) of detecting a CDR3 or its near-exact single AA variation neighbor across donors with class I or class II HLA alleles, computing a p-value by a Fisher’s exact test.

A TCR feature centroid was assigned to the most statistically significant HLA allele if it had an OR > 1 with a p-value < 1e-8, was detected in > 5% of HLA-matched individuals and not in more than 10% of HLA-mismatched individuals. From cohort 2, this procedure yielded 20,037 unique V-gene family-CDR3β centroids assigned to either a MHC class I allele (n = 7,255) or MHC class II allele(s) (n = 12,782). Next, for each strongly HLA-associated feature, we tabulated detection in Cohort 1 repertoires of participants with the hypothesized restricting HLA allele. (We considered a detection if the query TCR had an exact or near exact match to a TCR with a with-in repertoires frequency greater than 1 in 500,000 templates). Based on detections in HLA-matched repertoires we computed the odds ratio of observing the centroid or its neighbor in participants based on T1D clinical status and computed a p-value based on Fisher’s exact test (with the Python package *fishersapi* based on *fast-fisher*), and computed and adjusted p-value based on the Benjamini-Hochberg procedure (Benjamini & Hochberg, 1995) to control false discovery rate using the Python package *statsmodels*. To visualize potentially T1D underrepresented and overrepresented sequence motifs derived from sequences with a false discovery rate < 0.2 we clustered these sequences into graphs using the Python package *tcrdist3 (Mayer-Blackwell et al., 2021)* and *networkx (Hagberg et al., 2008)* at an edge threshold of 24 TCRdist units and plotted motifs based on aligned sequences within each graph connected component.

### Quantification of HLA-based restriction of CDR3β sequence diversity

We studied the association of the HLA and CDR3β sequences using published methods (Ishigaki et al., 2022) and observed the impact of T1D-risk HLA alleles on selective restriction of CDR3β sequences. The analysis was primarily performed on cohort 1 and observations were validated on cohort 2+3. To assess the robustness of the findings, we also performed the analogous analysis on T1D, FDR and CTRL repertoires separately. SDRs (59 repertoires) were not included in this analysis due to insufficient sample number. We considered A, B, C, DPA1, DPB1, DQA1, DQB1, DRB1 HLA haplotypes in the study and variations in the HLA alleles were obtained from genotyping experiments. The rare or common alleles for each HLA haplotype were removed from the analysis based on their frequency in cohort 1 (frequency ≥ 99% or <1%). Furthermore, four-digit classical alleles were partitioned based on AA polymorphism at the HLA site (Extended Data Fig. 3A).

#### Position-specific CDR3β AA frequency analysis

Firstly, CDR3β sequences were segregated based on the length. We considered CDR3β lengths ranging from 12 to 18 (described as L12–L18 in the manuscript) as most of the CDR3β sequences (94.17% of total sequences) were within this range (Extended Data Fig. 3B). All analyses focused on the international ImMunoGeneTics information system (IMGT) (Lefranc et al., 2009) CDR3 positions 107 to 116 that directly contact antigens (Glanville et al., 2017) (Extended Data Fig. 3B, F). The flanking positions in CDR3β sequences defined by germline-encoded V- or J-genes were identified using IMGT/HighV-QUEST for each length group and removed from the CDR3β sequences. The frequency of each AA was computed at every position within the CDR3β sequence for length L12 to L18. This computation involved dividing the total count of a particular AA by the count of all AAs at a specific position of CDR3β sequence (Extended Data Fig. 3B).

#### Multivariate multiple linear regression model (MMLR)-multivariate analysis of variance (MANOVA) analysis

In simplest terms, the MMLR analysis is performed using n-dimensional independent variables to predict the n-dimensional dependent variables. In our study, the frequency of all AAs at a specific position of the CDR3β sequence was used to calculate the association with counts of all AA polymorphism at an HLA site (Extended Data Fig. 3C) (Sakaue et al., 2023). We also included the top three principal components of all HLA genotypes in this analysis. Therefore, the full MMLR model was the following equation.

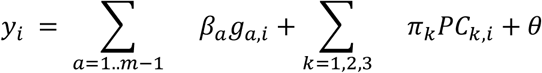

Here, 𝑦_𝑖_ is n-dimensional CDR3β amino acid frequency vector of 𝑖^𝑡ℎ^ repertoire, 𝛽_𝑎_ is an n-dimensional parameter that represents the additive effect per group of classical alleles containing the same AA at specific HLA site (denoted by 𝑎 in equation), 𝑔_𝑎,𝑖_ is the allele count of allele group 𝑎 in 𝑖^𝑡ℎ^ repertoire. We included m − 1 group of classical alleles, casting 1 group as the reference. 𝜋_𝑘_ is an n-dimensional parameter that represents the effect of the 𝑘^𝑡ℎ^ principal component and 𝑃𝐶_𝑘,𝑖_ is the value of the 𝑘^𝑡ℎ^ principal component of 𝑖^𝑡ℎ^ repertoire. θ is an n-dimensional parameter that represents the intercept. The null model only had the terms for covariates without allelic effects.

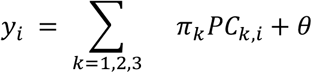

The improvement in the model fit between the full and null model was estimated using MANOVA and the significance of the improvement was assessed using Pillai’s trace. Codes from a previous study (https://github.com/immunogenomics/cdr3-QTL) were modified and reused accordingly in the study using Python and R languages (Ishigaki et al., 2022).

#### HLA risk score

The mutations in HLA-DQB1 at site 57 and HLA-DRB1 at site 13 and 71 have previously been associated with T1D disease progression based on 18,832 case-control samples of T1D (Hu et al., 2015). In this analysis, we grouped the classical four-digit HLA-DQB1 and HLA-DRB1 alleles based on the AAs present in these three sites along with the T1D risk in terms of OR (<u>Table S5</u>). The HLA risk score was calculated based on the sum of the individual OR scores multiplied by the number of alleles containing a specific AA polymorphism (Extended Data Fig. 3D). In case multiple combinations were possible for the three AAs, then we consider the combination with the highest OR score.

#### CDR3 risk score

The association of CDR3β amino acids with HLA risk score is referred to as CDR3 risk score. We developed linear regression (LR) models between the HLA risk score and CDR3β frequency of each AA at specific CDR3β position (for all lengths running from L12-L18) as described previously (Ishigaki et al., 2022; Sakaue et al., 2023). The p-values obtained from the LR model were further adjusted for multiple testing using the Benjamini–Hochberg method. The correlation coefficients of the significant association between the AA frequency and HLA risk score were denoted as effect size of that particular T1D-associated CDR3β AA or CDR3 phenotype (Extended Data Fig. 3E). The number of significant associations were dependent on the number of repertoires in the study. Therefore, we observed a higher number of associations for cohort 1.

The cumulative effect sizes of CDR3 phenotypes were calculated for the whole CDR3β sequence, and CDR3 risk score for the entire repertoire was derived by averaging the cumulative effect sizes of all CDR3β sequences (Extended Data Fig. 3E). Further, we calculated the correlation between HLA risk score and CDR3 risk score for each length and also compared CDR3 risk scores across clinical groups.

#### High-risk HLA-associated motif identification from CDR3 phenotypes

The CDR3 phenotypes, derived from the HLA risk score, were transformed into sequence motifs to evaluate their association with clinical groups. The AAs were grouped based on the IMGT positions, and a cutoff of effect sizes ≥0.1 was employed to select significant associations. Positive and negative effect sizes were analyzed separately to identify positively-associated (pHLA-motif) and negatively-associated (nHLA-motif) motifs with HLA-based T1D risk. The analysis was focused on CDR lengths L13 to L16, which had a higher occurrence in the repertoires (see Table S8). Furthermore, IMGT position aligned AAs showed the repetitive behavior of AAs on the C-terminal end, leading to selection of the conserved regions (IMGT positions 107-111). The effect sizes of consensus motifs in the logo plot represent the average effect sizes of CDR3 phenotypes for length L13 to L16.

#### Comparative analysis of CDR3 risk score across T1D clinical groups

We calculated CDR3 phenotypes and their effect sizes across repertoires from different clinical groups, including T1D, FDR and CTRL. Notably, an overlap was found in CDR3 phenotypes across the clinical groups. To appropriately test for association between CDR3 risk score and clinical groups, the CDR3 phenotypes and their corresponding effect sizes, derived from a specific clinical group, were applied to calculate the CDR3 risk score across all clinical groups. It is crucial to emphasize that the CDR3 phenotypes for each clinical group were computed independently of other clinical groups. As a control, we shuffled the effect sizes of the CDR3 phenotypes obtained from cohort 1 to highlight the importance of effect sizes and position-specific nature of the CDR3 phenotypes.

#### Analysis of heterozygous HLA allele based restriction of TCR diversity

We employed IGoR, a probabilistic model of V(D)J recombination, to model the recombination processes of unproductive TCR sequences. The sequences were aligned, and the recombination parameters were inferred through iterative learning. Multiple rounds of inference were performed to ensure robust parameter estimation. The inferred models were saved, and gene usage patterns were visualized for each sample. Note that raw sequences without any filtering criteria were used in the IGoR model (Marcou et al., 2018). Further, the productive TCR sequences were analyzed using SoNNia (Isacchini et al., 2021), a selection model that builds on the recombination probabilities learned by IGoR. Sonia inferred selection pressures by modeling the observed TCR sequences and generating a large number of synthetic sequences for comparison. The model was trained over 80 epochs to optimize selection parameters, and its performance was evaluated by comparing the generative (Pgen) and posterior (Ppost) sequence probabilities. Model performance was assessed by calculating the entropy of the sequence distributions both pre- and post-selection, which provided insights into the diversity and selection pressures within the TCR repertoire.

The HLA genes A, B, C, DPA1, DPB1, DQA1, DQB1, DRB1 were considered to assess the heterozygosity of HLA alleles. We calculated the total count of homozygous alleles in HLA genes (ranging from 0 being heterozygous and 8 being homozygous), and assuming the equal contribution of the HLA genes in the TCR diversity.

To assess the clustering of the v-gene frequencies, TRBV28 was used as a case study, where it was segregated as low-frequency (f < 0.015), mid-range frequency (0.015 ≤ f ≤ 0.04), and high-frequency (f > 0.04). For each subset, bootstrapped samples were generated by resampling 30% of the data multiple times, allowing for robust estimation of frequency distributions with respect to HLA alleles. The mean and standard deviation of allele frequencies were computed across the resampled datasets to capture the variability in the data.

### Machine learning approaches for repertoire classification and motif identification

#### ML analysis data preprocessing and general ML workflow

We employed a combination of machine learning and deep learning approaches to classify T1D status and identify T1D-associated motifs within immune repertoires. Cohort 1 served as the training set for all methods, while repertoires from Cohort 2 and 3 were used as a held-out test set. The T1D class was considered a positive class and remaining clinical statuses, FDR, SDR and CTRL, were considered non-T1D or negative class.

In the k-mer based logistic regression (LogReg) and DeepRC model, the same 5-fold cross-validation (CV) dataset was used to train both model parameters and hyper-parameters in cohort 1. The data was split into 5 random subsets, with T1D and control samples drawn separately without replacement for each fold. Each fold contained an approximately equal number of samples and maintained a consistent ratio of control to T1D samples. Moreover, each repertoire was assigned a weight based on the prevalence of their age group and the distribution of disease states across these groups, thereby normalizing the influence of age on the classification model (see **Supplementary Notes** for more detail). The first and last four AAs in each CDR3β sequence were also excluded to avoid the effect of V- and J-genes in ML/DL methods.

#### Statistical classification framework for T1D repertoires using public clones

We utilized the ImmuneML platform (Pavlović et al., 2021) to implement the statistical classification framework (Emerson et al., 2017) on the T1D dataset, which utilizes unique V-gene, CDR3 and J-gene information. The YAML file was prepared following the immuneML documentation (https://docs.immuneml.uio.no/latest/usecases/emerson_reproduction.html). In summary, we employed the ‘CompAIRRSequenceAbundance’ encoding based on the CompAIRR tool (Rognes et al., 2022) to enable faster repertoire level comparisons. Model performance was assessed across varying significance thresholds (p-values = [0.001, 0.0001, 0.00001, 0.000001]). The optimal model was selected based on optimization of log-loss values in 5-fold cross-validation. Log-loss measures the divergence between predicted probabilities and the actual class value, with lower log-loss indicating better performance. We further calculated the balanced accuracy and area under the ROC curve (AUROC) to analyze the performance of the model. The optimal model was further tested on cohort 2 and 3 to assess its predictive performance in unseen datasets.

#### K-mer based classification model using Logistic regression

The linear subsequence information at the 4-mer level was employed as a feature set to classify T1D status. A 4-mer frequency matrix of 160,000 (20^4^; number of possible 4-mers) by 1298 (number of repertoires in cohort 1) was generated. To populate this matrix, we used a sliding window approach, where a window of 4 consecutive AA moved across each CDR3β sequence. The frequency of occurrence of each unique 4-mer was recorded in the matrix for its respective repertoire. This 4-mer frequency matrix was then used as a feature set in a LogReg to classify the T1D statuses of immune repertoires. For model training, we applied a 5-fold CV on Cohort 1, where 3 folds were used for training, 1 fold for tuning, and 1 fold for testing, with the process repeated recursively. The outputs of the 5 models, one from each fold, were then assembled into a single model by averaging their predictions after applying the sigmoidal activation function. This ensembled model was subsequently tested on a held-out dataset consisting of Cohorts 2 and 3. The following hyperparameter ranges were used for the training of LogReg model:

|  |  |  |
| --- | --- | --- |
| Penalty | : | L1, L2, Elastic-Net penalty or no penalty |
| C | : | random value in [0.1, 10) |
| Solver algorithms | : | Newton-CG, LBFGS, Liblinear, SAG, SAGA |
| Number of iterations | : | {100, 500, 1000, 5000} |

#### Deep learning-based multiple instance learning model for classification of T1D status

##### Development of DeepRC model

We utilized DeepRC (Widrich et al., 2020), a DL method based on continuous modern Hopfield networks (MHN) (Ramsauer et al., 2020), within a multiple instance learning framework to classify immune repertoires based on T1D status. In this method, each TCR repertoire was treated as a sample, classified as either T1D or CTRL, while individual CDR3β sequences within the repertoire were viewed as instances. Thus, each sample is represented as a "bag" of CDR3β sequences with a single class label.

The DeepRC architecture used a convolutional neural network (CNN) as an encoder for processing individual sequences. Input vectors were structured with dimensions corresponding to sequence_length × n_amino_acids, where the sequence length may vary across sequences. A learned kernel of shape (kernel_size x n_amino_acids) was convolved along the AA positions of each CDR3β sequence, and feature-wise max-pooling was applied to obtain a fixed-sized feature vector of shape (n_sequences x kernel_size), where n_sequences may differ between repertoires. A learned MHN attention pooling mechanism, as described in (Widrich et al., 2020), was used to pool the instances in a repertoire of shape (n_sequences x kernel_size) into a single feature vector of shape (kernel_size). This results in a single feature vector of shape (kernel_size) as representation for a single repertoire. Finally, the feature vector was passed to a fully connected output layer, consisting of one output unit, to predict the class label of the repertoire.

All trainable parameters in the DeepRC architecture were optimized end-to-end using the Adam optimizer in PyTorch, with a binary cross-entropy loss function. Similar to LogReg, we assigned 3 folds for training, 1 fold as tuning, and the remaining 1 fold as test set, recursively, in a 5 fold CV setup. We train the DeepRC model on the training set while using the tuning set loss as early stopping criteria. The best-performing model for each CV fold was selected based on the area under the receiver operating characteristic curve (AUROC) of the tuning set, resulting in 5 distinct "best" models from the CV process. Subsequently, a LogReg model was trained to aggregate the predictions from these 5 “best” models, forming the final DeepRC model, which was then evaluated on the held-out test set. Following parameters were used for the training of DeepRC model:

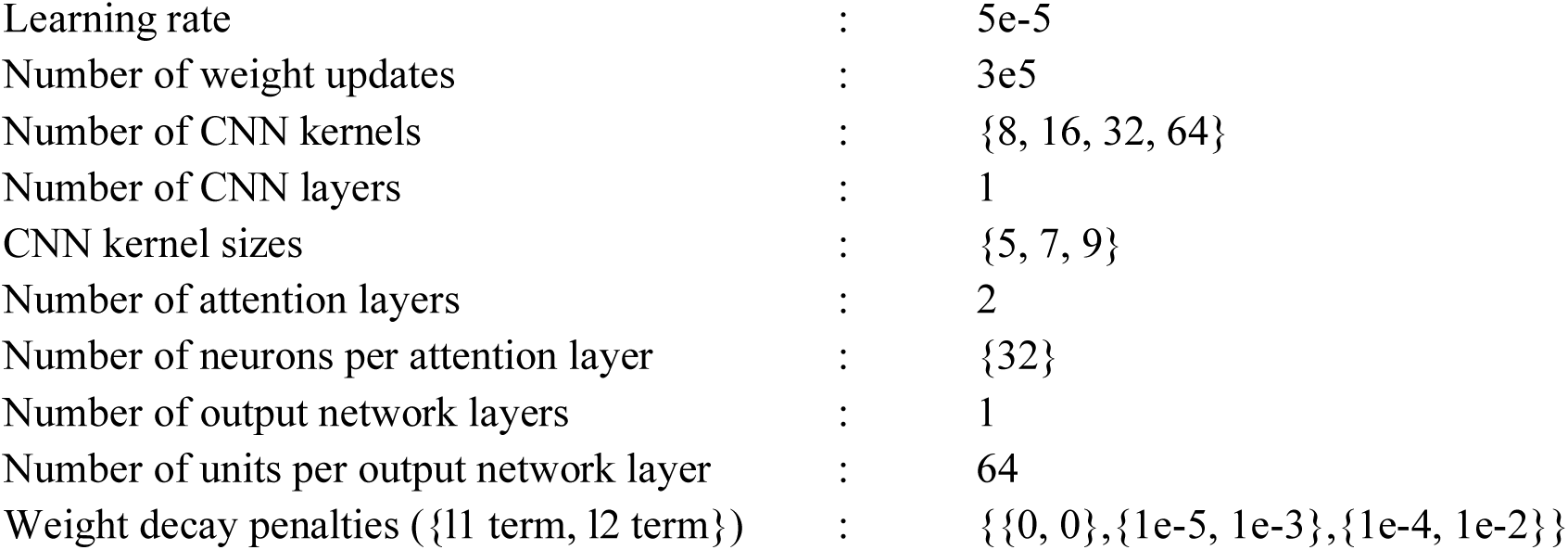

|  |  |  |
| --- | --- | --- |
| Learning rate | : | 5e-5 |
| Number of weight updates | : | 3e5 |
| Number of CNN kernels | : | {8, 16, 32, 64} |
| Number of CNN layers | : | 1 |
| CNN kernel sizes | : | {5, 7, 9} |
| Number of attention layers | : | 2 |
| Number of neurons per attention layer | : | {32} |
| Number of output network layers | : | 1 |
| Number of units per output network layer | : | 64 |
| Weight decay penalties ({l1 term, l2 term}) | : | {{0, 0},{1e-5, 1e-3},{1e-4, 1e-2}} |

##### T1D-associated motif identification from DeepRC

DeepRC supports different ways of interpretability, via the attention values and via the contribution analysis method Integrated Gradients (IG) (Sundararajan et al., 2017). We applied the IG method to the trained DeepRC model from each cross-validation fold as described in the original paper (Widrich et al., 2020). It allows visualization of the contribution of inputs and weights to the prediction of the DeepRC models, which could then be used to manually extract motifs. For each DeepRC model in the cross-validation, we compute the Integrated Gradients (IG) such that we obtain the contribution of the input sequences to the DeepRC model output. In this step, we compute IG on the T1D positive samples of the tuning set samples that were also used for early stopping. The training split samples were not used here as they have already been overfitted by the DeepRC model. Subsequently, for each DeepRC model, we identified the input sequence with the highest contribution to the T1D prediction per repertoire. The DeepRC model with null coefficient in the ensemble LogReg model was excluded from the motif calculation. The identified sequences from each DeepRC model, corresponding to T1D-positive repertoires in the tuning set, were collected into a FASTA file and subsequently analyzed with GLAM2 (Frith et al., 2008) to compute motifs through sequence alignment.

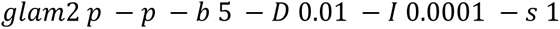

We chose a motif width of -b 5, as it corresponds to the smallest motif width matching the CNN kernel size. The first and last 4 AA of every input sequence were cropped before alignment. The final selected motif [FWY],([AFHILMPQRSTVY]){0,1},[EFGHSWY],([AFQT]){0,1},[ACDEGHIKNPQRSTVY] contains two deletions at position 2 and 4 in the motif (Extended Data Fig. 9).

Consensus sequences containing both the pHLA-motif and DeepRC-motif were obtained by sequentially filtering the T1D dataset: first by the presence of DeepRC-motif, followed by further filtering based on presence of pHLA-motif. The resulting intersection of the pHLA-motif and DeepRC-motif is termed the “consensus-motif”. Similar to other motifs, the consensus-motif score was calculated by normalizing the number of motif-containing sequences with the total CDR3β sequence count in the respective repertoire.

### Quantitative Trait Locus Analysis

QC measures were performed to remove data from individuals that were sex discordant, related, and/or showed unusual levels of heterozygosity, as previously described (Shapiro et al., 2023). Genetic ancestry was inferred using Admixture software (Alexander et al., 2009) for projection analysis on the 1000 Genomes cohort(1000 Genomes Project Consortium et al., 2015). Quantitative trait loci (QTL) analysis was performed to detect associations between TCR motif frequency versus 240 T1D risk variants (Shapiro et al., 2023) that were directly genotyped or imputed, as previously described. For PBMC data reported in this publication, linear regression was performed assuming additive genotypic effect with age, sex, T1D status, predicted probability of CMV infection (Shapiro et al., 2023), and 10 multidimensional scaling components as covariates. For nPOD tissue data, age, sex, diabetes status, and 10 multidimensional scaling components were included and for sorted peripheral blood data, age, sex, and T1D status were included as covariates.

## Data availability

Study data is available for download from the AIRR Data Commons (Christley et al., 2020) using the iReceptor Gateway (Corrie et al., 2018) using Study ID IR-T1D-000004. This data was curated as part of the AIRR T1D Consortium initiative (Hanna et al., 2024). Sequence data is also publicly available through Adaptive Biotechnologies ImmuneACCESS site. The data will be published upon completion of the review process. The code and data used in this study are available at the following GitHub repository: https://github.com/csi-greifflab/T1D-TCR.

## Data Availability

All data produced in the present study will be available upon publication in the journal

## Acknowledgement

We thank the study participants for the gift of blood sample donation, without which this work would not have been possible.

## Author contributions

PR: Software, Formal Analysis, Investigation, Writing-Original Draft, Writing-Review & Editing, Visualization

MRS: Software, Formal Analysis, Writing-Original Draft, Writing-Review & Editing, Visualization

LDP: Software, Formal Analysis, Writing-Original Draft, Writing-Review & Editing, Visualization

MW: Methodology, Software, Formal Analysis, Writing-Review & Editing

KMB: Software, Formal Analysis, Writing-Original Draft, Writing-Review & Editing

KeM: Investigation, Data Curation, Writing-Review & Editing

MP: Software, Writing-Review & Editing

GAH: Software

ALP: Writing-Review & Editing, Project Administration

CK: Software, Writing-Review & Editing

GI: Software, Writing-Review & Editing

MC: Methodology, Writing-Review & Editing

LS: Software

KaM: Methodology, Writing-Review & Editing

LOB-N: Software, Writing-Review & Editing

CPW: Software, Writing-Review & Editing

SV: Software, Writing-Review & Editing

LMJ: Methodology, Writing-Review & Editing

MJH: Resources, Writing-Review & Editing

DAS: Resources, Writing-Review & Editing

CHW: Writing-Review & Editing

ROE: Data Curation, Supervision

AJFG: Supervision, Writing-Review & Editing

MAA: Writing-Review & Editing, Funding Acquisition

GK: Writing-Review & Editing, Supervision

GKS: Conceptualization, Writing-Review & Editing, Supervision

VG: Conceptualization, Writing-Review & Editing, Supervision, Funding Acquisition

TMB: Conceptualization, Resources, Writing-Review & Editing, Supervision, Funding Acquisition

## Extended Data Figures

Extended Data Fig. 1. **The publicly available inflammatory disease and T1D-associated CDR3β sequences demonstrate association with clinical groups in cohort 1.** The unique clone count of CDR3β sequences in cohort 1, which were also present in the McPAS/VDJdb database for given pathology (as detailed in <u>Table S1</u>), were averaged for each repertoire and normalized by the total clone count. These normalized values were further clustered based on the clinical groups and visualized in log scale. (**A**) Sequence matches were calculated for different pathologies in McPAS database. (**B**) The distribution of T1D pathology-associated CDR3β sequences in each repertoire. individual T1D-associated CDR3β sequences revealed a small subset of potential false positives in the McPAS database, which were observed in almost all repertoires. Manually curated databases are subject to bias based on a priori knowledge and experimental design. Thus, our results provide further support for utilization of machine learning and other unbiased methodologies to classify T1D repertoires. (**C**) Sequence matches were calculated for different antigen species in the VDJdb database. **(D)** Association of 25,508 unique CDR3β sequences associated with cytomegalovirus (CMV) exposure with clinical groups. In the figure, multiple testing was performed using Kruskal–Wallis test (denoted with red stars) and p-values were adjusted between pathology/antigen species. p-values for pairwise testing were calculated using two tailed Mann-Whitney U test and p-values were adjusted between different clinical groups. All p-values were adjusted for multiple testing using the Benjamini–Hochberg method. p-values were described as * for [0.01,0.05], ** for [0.001,0.01] and *** for <0.001 and no stars plotted for non-significant values. Cluster plot was generated using the UPGMA clustering method and Euclidean distance matrix. A universal color scheme was used to show type 1 diabetes (T1D), first degree relatives (FDR), second degree relatives (SDR) and control (CTRL) in red, yellow, light green and dark green, respectively.

Extended Data Fig. 2. **T1D-association of TCR features observed based on assessment of HLA-associated, TRBV-gene family constrained and single amino acid mutation distance neighbor CDR3β sequences.** (**A**) TRBV-gene family and HLA-associated CDR3β sequences depleted (blue) or overrepresented (red) in the T1D repertoires (false discovery rate < 0.2), when considering detection of a CDR3 or a nearly-identical CDR3 with only one indel or substitution. (**B**) Gene usage and CDR3β(AA) motifs formed from top sequence clusters overrepresented in T1D repertoires. **(C**) Gene usage and CDR3β(AA) motifs formed from top sequence clusters underrepresented in T1D repertoires. HLA association determined by Fisher Exact Test p-value < 1E-8, HLA-prevalence > 0.05, non-HLA-prevalence < 0.1 in Cohort2. Relates to Fig. 3.

Extended Data Fig. 3. **The methodology used to assess the association between HLA and CDR3β. (A)** The information related to amino acid (AA) polymorphism present at the HLA site was obtained from the HLA genotyping experiments. Here, HLA-DRB1 is given as an example, which has 4 AA variants (K, A, E, R) at site 71. All possible AA variant counts were calculated for each repertoire. **(B)** The histogram (left) shows the distribution of the CDR3β length in cohort 1 (for lengths containing more than 30,000 CDR3β sequences). The position-specific AA frequency was calculated for each CDR3β sequence with length ranging from L12 to L18 after removing the V- and J-germline-encoded flanking regions (right). In the example, we have removed the N-terminal sequences based on specific V-genes (“CASSE” for TRBV02-01 and “CASSQ” for TRBV04-01). The V and J germline-encoded sequences were removed using IMGT/HighV-QUEST tool. Further, the frequency of all AAs was calculated for each position and for each length (L12-L18) of CDR3β sequences. **(C)** The multivariate multiple linear regression model (MMLR) was calculated for each HLA site polymorphism, where count of AA variants at the site was explanatory variable and AA frequency of specific CDR3β position was response variable. This was done for each IMGT position of CDR3β sequence for all lengths (L12-L18), recursively. **(D)** The HLA risk score was calculated based on the 3 AA positions: HLA-DQβ1 (site 57) and HLA-DRβ1 (site 13 and 71). The odds ratio (OR) for possible combinations of three AAs was provided in a previous study (Hu et al., 2015), which was multiplied by the individual’s genotype. In case multiple combinations were possible for the three AAs, then we consider the combination with the highest OR score. **(E)** In the first step to calculate CDR3 risk score, we considered HLA risk score and position-specific frequency of one AA to develop several linear regression models (LR) for each CDR3β position and length, recursively. The p-values were further adjusted for multiple testing using the Benjamini–Hochberg method. The AAs which showed significant correlations with HLA risk score (p-value≤0.05) were considered “T1D-associated position-specific CDR3β AA” or “CDR3 phenotypes” and respective correlation coefficients were considered the “effect sizes” associated with T1D. Within a repertoire, the effect sizes of each CDR3 phenotype were summed up for the whole CDR3β sequence, and averaged for the whole repertoire to calculate CDR3 risk score. **(F)** In our study, the CDR3β positions for CDR3β length L12 to L18 were defined using the IMGT numbering scheme, which are highlighted in blue color in the figure. The examples in the figures are taken directly or inspired from Iskigaki et al (Ishigaki et al., 2022). Relates to Fig. 3.

Extended Data Fig. 4. **The assessment of the pHLA-motif, derived from the HLA risk score, shows positive association with the clinical groups, high-risk HLA alleles (DR3/DR4) and autoantibody presence. (A)** A sequence logo representation of the pHLA-motif. The consensus motif was derived from the CDR3β lengths L13-16 and IMGT positions 107-111. **(B)** The motif score was higher for T1D repertoires in cohort 1, following the expected trend of T1D > FDR > SDR > CTRL, with SDR being an exception. A similar pattern was also observed in cohorts 2 and 3. The pHLA-motif score represents the number of CDR3β sequences containing the pHLA-motifs normalized by total number of CDR3β sequences per repertoire, where higher pHLA-motif score signifies higher risk of T1D. **(C)** The correlation between T1D duration and motif score shows a pearson correlation of -0.18 and spearman correlation of -0.16. **(D)** There was no trend observed in V-gene usage across TCRβ repertoires of different clinical groups, when considering the CDR3β sequences containing pHLA-motifs. **(E)** The pHLA-motif score was significantly higher in the high-risk HLA type (DR3 or DR4) individuals than other HLA types (DRX/X). The p-values in the figure were calculated with respect to the DRX/X HLA type. **(F)** The pHLA-motif score was plotted for each high-risk HLA type and grouped by the clinical groups.The pHLA-motif score observed the expected trend T1D>FDR>CTRL in all cases irrespective of high-risk HLA allele type. Here, pairwise testing was performed with respect to the T1D clinical group. **(G)** The pHLA-motif score also showed positive association with the number of autoantibodies present. The p-values were calculated with respect to no presence of autoantibodies. **(H)** The pHLA-motif score was high for autoantibody count >0. However, in case of no presence of autoantibody, motif score followed the expected trend where T1D>FDR>CTRL. **(I)** The pHLA-motif score was plotted against age of the patient for different clinical groups (for cohort 1). Each T1D status showed negative correlation with the age of the patient. In the figure, the p-values for the multiple testing were performed using Kruskal–Wallis test (denoted with red stars). Similarly, p-values for pairwise testing were calculated using two tailed Mann-Whitney U tests. p-values were described as * for [0.01,0.05], ** for [0.001,0.01] and *** for <0.001 and no stars plotted for non-significant values. All p-values were adjusted for multiple testing using the Benjamini– Hochberg method. Relates to Fig. 3.

Extended Data Fig. 5. **The assessment of the nHLA-motif (protective motif), derived from the HLA risk score, shows negative association with the clinical groups, high-risk HLA alleles (DR3/DR4) and autoantibody presence. (A)** A sequence logo representation of the nHLA-motif. The consensus motif was derived from the CDR3β lengths L13-16 and IMGT positions 107-111. **(B)** The motif score was higher for non-T1D repertoires in cohort 1, following the expected trend of T1D < FDR < SDR < CTRL, with SDR being an exception. A similar pattern was also observed in cohorts 2 and 3. The nHLA-motif score represents the number of CDR3β sequences containing the nHLA-motifs normalized by total number of CDR3β sequences per repertoire, where higher nHLA-motif score signifies lower risk of T1D. **(C)** The nHLA-motif score from the protective motif showed no correlation with the T1D duration (pearson correlation of 0.05 and spearman correlation of 0.07). **(D)** The V-genes with higher frequency had relatively lower usage in the T1D repertoires and had an observable trend of T1D<FDR<CTRL, when considering the CDR3β sequences containing the nHLA-motif. **(E)** The repertoires containing at least one DR3 allele exhibited nHLA-motif scores comparable to non-risk repertoires, indicating that DR3 alleles may possess more protective motifs. **(F)** The nHLA-motif score was plotted for each high-risk HLA type and grouped by the clinical groups. The nHLA-motif score was significantly higher for CTRL repertoires in cases both HLA alleles were not associated with high-risk of T1D (DRX/X) and observed the expected trend T1D<FDR<CTRL. Here, pairwise testing was performed with respect to the T1D clinical group. **(G)** The nHLA-motif score was higher for the repertoires with no or just one autoantibody present. The p-values were calculated with respect to no autoantibody presence. **(H)** The nHLA-motif score observed association with clinical groups in repertoires with no autoantibody presence with an expected trend where T1D<FDR<CTRL. **(I)** The nHLA-motif score was plotted against age of the patient for different clinical groups (for cohort 1). No correlation was observed for the nHLA-motif score with age of the patient. In the figure, the p-values for the multiple testing were performed using Kruskal–Wallis test (denoted with red stars). Similarly, p-values for pairwise testing were calculated using two tailed Mann-Whitney U tests. p-values were described as * for [0.01,0.05], ** for [0.001,0.01] and *** for <0.001 and no stars plotted for non-significant values. All p-values were adjusted for multiple testing using the Benjamini– Hochberg method. Relates to Fig. 3.

Extended Data Fig. 6. **Heterozygous HLA alleles showed restriction of the TCR repertoire diversity in cohort 1. (A)** The pre-selection TCR repertoire diversity (Pgen, IGoR) and **(B)** post-selection repertoire diversity (Ppost, SoNNia) calculated with respect to count of homozygous HLA alleles for 8 HLA genes (A, B, C, DPA1, DPB1, DQA1, DQB1, DRB1) showed a trend where repertoires with more heterozygous HLA alleles had lower diversity in both pre and post selection. **(C)** V-gene usage frequency calculated from the IGoR model. Some of the V-genes including TRBV28, TRBV4-3, TRBV3-2, TRBV3-1 showed clustering of v-gene frequency. **(D)** as a case study, the three v-gene frequency clusters of TRBV28 were assessed for their potential link with the HLA genes. The HLA alleles with frequency >0.01 were considered in the plot. The error bars were plotted by bootstrapping samples, where 30% of the data resampled multiple times. In figure **A** and **B**, p-values for pairwise testing were calculated using two tailed Mann-Whitney U tests with respect to most heterozygous HLA individuals (number of homozygous alleles = 0). p-values were described as * for [0.01,0.05], ** for [0.001,0.01] and *** for <0.001 and no stars plotted for non-significant values. All p-values were adjusted for multiple testing using the Benjamini–Hochberg method.

Extended Data Fig. 7. **Assessment of HLA risk score demonstrates that HLA alleles are robust predictors of T1D status (A)** Confusion matrices were generated using high-risk HLA allele types (DR3 and DR4) as predictor of T1D status under two scenarios: (i) considering the presence of at least one high-risk HLA allele as indication of T1D status, and (ii) considering the presence of both high-risk HLA allele as indication of T1D status. The confusion matrices were generated separately for cohort 1 and cohort 2+3 (see **Supplementary Note** for more detail). **(B)** The HLA risk score was calculated based on the 3 AA positions: HLA-DQβ1 (site 57) and HLA-DRβ1 (site 13 and 71). Further, HLA risk score was used for binary classification of T1D positive and negative repertoires. **(C)** Expectedly, T1D duration does not show correlation with HLA risk score (pearson correlation: 0.11 and spearman correlation: 0.14). **(D)** HLA risk score in the repertoires with high-risk HLA alleles type (DR3 or DR4) was significantly higher compared to other HLA types (DRX/X). The p-values in the figure were calculated with respect to the DRX/X HLA type. **(E)** The HLA risk score was plotted for each high-risk HLA type and grouped by the clinical groups. The HLA risk score was significantly high in all clinical groups containing both alleles associated with high-risk (DR3 and DR4). However, it observed the expected trend T1D>FDR>SDR>CTRL in cases where at least one HLA allele was not associated with high-risk of T1D. Here, pairwise testing was performed with respect to the T1D clinical group. **(F)** The HLA risk score also observed positive association with the number of autoantibodies present. The p-values were calculated with respect to no presence of autoantibodies. **(G)** HLA risk score was high for all individuals when autoantibody count was >0. However, in case of no presence of autoantibody, HLA risk score followed the expected trend where T1D>FDR>SDR>CTRL. **(H)** The HLA risk score was plotted against age of the patient for different clinical groups (for cohort 1) and as expected, it did not show any correlation with the age of the patient. In the figure, the p-values for the multiple testing were performed using Kruskal–Wallis test (denoted with red stars). Similarly, p-values for pairwise testing were calculated using two tailed Mann-Whitney U tests. p-values were described as * for [0.01,0.05], ** for [0.001,0.01] and *** for <0.001 and no stars plotted for non-significant values. All p-values were adjusted for multiple testing using the Benjamini–Hochberg method. Relates to Fig. 4.

Extended Data Fig. 8. **Assessment of the DeepRC model performance demonstrates that TCR repertoires can be robust predictors of T1D risk. (A)** DeepRC model score in the repertoires with high-risk HLA alleles type (DR3 or DR4) was significantly higher compared to other HLA types (DRX/X). The p-values in the figure were calculated with respect to the DRX/X HLA type. **(B)** The DeepRC model score was plotted for each high-risk HLA type and grouped by the clinical groups. The DeepRC model score was significantly high in all clinical groups containing both alleles associated with high-risk (DR3 and DR4). However, it observed the expected trend T1D>FDR>CTRL in cases where at least one HLA allele was not associated with high-risk of T1D. Here, pairwise testing was performed with respect to the T1D clinical group. **(C)** The DeepRC model score also observed positive association with the number of autoantibodies present. The p-values were calculated with respect to no presence of autoantibodies. **(D)** DeepRC model score was high for all individuals when autoantibody count was >0. However, in case of no presence of autoantibody, DeepRC model score followed the expected trend where T1D>FDR>CTRL. **(E)** The DeepRC model score was plotted against age of the patient for different clinical groups (for cohort 1) and a negative correlation was observed for all clinical groups. **(F)** Interestingly, T1D duration observed a positive correlation with DeepRC model score (pearson correlation: 0.19 and spearman correlation: 0.09). **(G)** The final ensemble logistic regression (LR) model, built using the outputs of each DeepRC model, assigned weights to each fold. These ensemble weights, along with the performance (AUROC curve) of the DeepRC model for each fold, were also presented. In the figure, the p-values for the multiple testing were performed using Kruskal–Wallis test (denoted with red stars). Similarly, p-values for pairwise testing were calculated using two tailed Mann-Whitney U tests. p-values were described as * for [0.01,0.05], ** for [0.001,0.01] and *** for <0.001 and no stars plotted for non-significant values. All p-values were adjusted for multiple testing using the Benjamini–Hochberg method. Relates to Fig. 4.

Extended Data Fig. 9. **Assessment of the TCR repertoires-derived, interpretable version of the DeepRC model, referred to as DeepRC-motif, demonstrates a robust association with T1D status.** The DeepRC-motif score represents the number of CDR3β sequences containing the DeepRC-motif normalized by total number of CDR3β sequences per repertoire, where higher DeepRC-motif score signifies higher risk of T1D. **(A)** The sequence logo representation of the DeepRC-motif. **(B)** DeepRC-motif score was significantly higher for T1D repertoires and observed the expected trend T1D>FDR>SDR>CTRL. **(C)** T1D duration observed a negative correlation with DeepRC-motif score (pearson correlation: -0.13 and spearman correlation: -0.12). **(D)** DeepRC-motif score in the repertoires with high-risk HLA alleles type (DR3 or DR4) was significantly higher compared to other HLA types (DRX/X). The p-values in the figure were calculated with respect to the DRX/X HLA type. **(E)** The DeepRC-motif score was plotted for each high-risk HLA type and grouped by the clinical groups. The DeepRC-motif score was significantly high in all clinical groups containing both alleles associated with high-risk (DR3 and DR4). However, it observed the expected trend T1D>FDR>CTRL in cases where at least one HLA allele was not associated with high-risk of T1D. Here, pairwise testing was performed with respect to the T1D clinical group. **(F)** The DeepRC-motif score also observed positive association with the number of autoantibodies present. The p-values were calculated with respect to no presence of autoantibodies. **(G)** DeepRC-motif score was high for all individuals when autoantibody count was >0. However, in case of no presence of autoantibody, DeepRC-motif score followed the expected trend where T1D>FDR>CTRL. **(H)** The DeepRC-motif score was plotted against age of the patient for different clinical groups (for cohort 1) and a negative correlation was observed for all clinical groups. In the figure, the p-values for the multiple testing were performed using Kruskal–Wallis test (denoted with red stars). Similarly, p-values for pairwise testing were calculated using two tailed Mann-Whitney U tests. p-values were described as * for [0.01,0.05], ** for [0.001,0.01] and *** for <0.001 and no stars plotted for non-significant values. All p-values were adjusted for multiple testing using the Benjamini– Hochberg method. Relates to Fig. 4.

Extended Data Fig. 10. **Positively-associated HLA-motif (pHLA-motif) enriched in carriers of T1D risk genetics in bulk and sorted CD4^+^ Treg and TCM from peripheral blood as well as pancreatic lymph node CD8^+^ T cells.** (**A**) Manhattan plot of 240 T1D risk variants versus pHLA-motif score in bulk peripheral blood. Linear regression assuming additive genotypic effect with age, sex, T1D status, predicted probability of CMV infection, and 10 multidimensional scaling components as covariates. Benjamini-Hochberg false discovery rate threshold at p=2.58e-4 (red line) and a conventional threshold of p = 0.05 (blue line). **(B)** Violin plots of pHLA-motif score in European ancestry non-diabetic (ND) subjects, according to significantly associated variants in **(A)**. Tagged HLA genotype and risk allele annotated above each plot. (**C**) pHLA-motif score across sorted T cell subsets in spleen and pancreatic lymph node (pLN). **(i)** Mixed-effects analysis with Tukey’s multiple comparisons test. (**ii**) pLN CD8^+^ T cell pHLA-motif score according to T1D status. (**D**) pHLA-motif score in sorted peripheral blood CD4^+^ T cell subsets. **(i)** Mixed-effects analysis with Tukey’s multiple comparisons test. (**ii**) Treg and (**iii**) CD4^+^ TCM pHLA-motif score according to DQA1*05:01-DQB1*02:01 tag SNP. Linear regression with age, sex, and T1D status as covariates. In all violin plots, p-values for pairwise testing were calculated using two tailed Mann-Whitney U tests and p-values were adjusted between different clinical groups using the Benjamini–Hochberg method. p-values were described as * for [0.01,0.05], ** for [0.001,0.01] and *** for <0.001 and no stars plotted for non-significant values. Relates to Fig. 5.

## References

1000 Genomes Project Consortium, Auton, A., Brooks, L. D., Durbin, R. M., Garrison, E. P., Kang, H. M., Korbel, J. O., Marchini, J. L., McCarthy, S., McVean, G. A., & Abecasis, G. R. (2015). A global reference for human genetic variation. Nature, *526*(7571), 68–74.

Achenbach, P., Bonifacio, E., Koczwara, K., & Ziegler, A.-G. (2005). Natural history of type 1 diabetes. Diabetes, 54 Suppl 2(suppl_2), S25–S31.

Alexander, D. H., Novembre, J., & Lange, K. (2009). Fast model-based estimation of ancestry in unrelated individuals. Genome Research, 19(9), 1655–1664.

Aly, T. A., Ide, A., Jahromi, M. M., Barker, J. M., Fernando, M. S., Babu, S. R., Yu, L., Miao, D., Erlich, H. A., Fain, P. R., Barriga, K. J., Norris, J. M., Rewers, M. J., & Eisenbarth, G. S. (2006). Extreme genetic risk for type 1A diabetes. Proceedings of the National Academy of Sciences of the United States of America, 103(38), 14074–14079.

Amoriello, R., Chernigovskaya, M., Greiff, V., Carnasciali, A., Massacesi, L., Barilaro, A., Repice, A. M., Biagioli, T., Aldinucci, A., Muraro, P. A., Laplaud, D. A., Lossius, A., & Ballerini, C. (2021). TCR repertoire diversity in Multiple Sclerosis: High-dimensional bioinformatics analysis of sequences from brain, cerebrospinal fluid and peripheral blood. EBioMedicine, 68, 103429.

Arnaout, R. A., Prak, E. T. L., Schwab, N., Rubelt, F., & Adaptive Immune Receptor Repertoire Community. (2021). The Future of Blood Testing Is the Immunome. Frontiers in Immunology, *12*, 626793.

Ashby, K. M., & Hogquist, K. A. (2024). A guide to thymic selection of T cells. Nature Reviews. Immunology, 24(2), 103–117.

Atkinson, M. A., & Eisenbarth, G. S. (2001). Type 1 diabetes: new perspectives on disease pathogenesis and treatment. Lancet, 358(9277), 221–229.

Bagaev, D. V., Vroomans, R. M. A., Samir, J., Stervbo, U., Rius, C., Dolton, G., Greenshields-Watson, A., Attaf, M., Egorov, E. S., Zvyagin, I. V., Babel, N., Cole, D. K., Godkin, A. J., Sewell, A. K., Kesmir, C., Chudakov, D. M., Luciani, F., & Shugay, M. (2020). VDJdb in 2019: database extension, new analysis infrastructure and a T-cell receptor motif compendium. Nucleic Acids Research, 48(D1), D1057–D1062.

Bashford-Rogers, R. J. M., Palser, A. L., Huntly, B. J., Rance, R., Vassiliou, G. S., Follows, G. A., & Kellam, P. (2013). Network properties derived from deep sequencing of human B-cell receptor repertoires delineate B-cell populations. Genome Research, 23(11), 1874–1884.

Beck, R. W., Tamborlane, W. V., Bergenstal, R. M., Miller, K. M., DuBose, S. N., Hall, C. A., & T1D Exchange Clinic Network. (2012). The T1D Exchange clinic registry. The Journal of Clinical Endocrinology and Metabolism, *97*(12), 4383–4389.

Benjamini, Y., & Hochberg, Y. (1995). Controlling the false discovery rate: A practical and powerful approach to multiple testing. Journal of the Royal Statistical Society, 57(1), 289–300.

Berger, W. H., & Parker, F. L. (1970). Diversity of planktonic foraminifera in deep-sea sediments. Science, 168(3937), 1345–1347.

Bettelli, E., & Campbell, D. J. (2020). Circulating TFH cells as a marker for early therapeutic intervention in T1D. Nature Immunology, 21(10), 1141–1142.

Boughter, C. T., & Meier-Schellersheim, M. (2023). Conserved biophysical compatibility among the highly variable germline-encoded regions shapes TCR-MHC interactions. eLife, 12. 10.7554/eLife.90681

Britanova, O. V., Lupyr, K. R., Staroverov, D. B., Shagina, I. A., Aleksandrov, A. A., Ustyugov, Y. Y., Somov, D. V., Klimenko, A., Shostak, N. A., Zvyagin, I. V., Stepanov, A. V., Merzlyak, E. M., Davydov, A. N., Izraelson, M., Egorov, E. S., Bogdanova, E. A., Vladimirova, A. K., Iakovlev, P. A., Fedorenko, D. A., … Chudakov, D. M. (2023). Targeted depletion of TRBV9+ T cells as immunotherapy in a patient with ankylosing spondylitis. Nature Medicine, 29(11), 2731–2736.

Britanova, O. V., Putintseva, E. V., Shugay, M., Merzlyak, E. M., Turchaninova, M. A., Staroverov, D. B., Bolotin, D. A., Lukyanov, S., Bogdanova, E. A., Mamedov, I. Z., Lebedev, Y. B., & Chudakov, D. M. (2014). Age-related decrease in TCR repertoire diversity measured with deep and normalized sequence profiling. Journal of Immunology , 192(6), 2689–2698.

Brown, A. J., White, J., Shaw, L., Gross, J., Slabodkin, A., Kushner, E., Greiff, V., Matsuda, J., Gapin, L., Scott-Browne, J., Kappler, J., & Marrack, P. (2024). MHC heterozygosity limits T cell receptor variability in CD4 T cells. Science Immunology, 9(97), eado5295.

Carlson, C. S., Emerson, R. O., Sherwood, A. M., Desmarais, C., Chung, M.-W., Parsons, J. M., Steen, M. S., LaMadrid-Herrmannsfeldt, M. A., Williamson, D. W., Livingston, R. J., Wu, D., Wood, B. L., Rieder, M. J., & Robins, H. (2013). Using synthetic templates to design an unbiased multiplex PCR assay. Nature Communications, 4, 2680.

Cerosaletti, K., Barahmand-Pour-Whitman, F., Yang, J., DeBerg, H. A., Dufort, M. J., Murray, S. A., Israelsson, E., Speake, C., Gersuk, V. H., Eddy, J. A., Reijonen, H., Greenbaum, C. J., Kwok, W. W., Wambre, E., Prlic, M., Gottardo, R., Nepom, G. T., & Linsley, P. S. (2017). Single-Cell RNA Sequencing Reveals Expanded Clones of Islet Antigen-Reactive CD4+ T Cells in Peripheral Blood of Subjects with Type 1 Diabetes. Journal of Immunology , 199(1), 323–335.

Chang, Y. M., Wieland, A., Li, Z.-R., Im, S. J., McGuire, D. J., Kissick, H. T., Antia, R., & Ahmed, R. (2020). T Cell Receptor Diversity and Lineage Relationship between Virus-Specific CD8 T Cell Subsets during Chronic Lymphocytic Choriomeningitis Virus Infection. Journal of Virology, 94(20). 10.1128/JVI.00935-20

Chen, L.-K., Chou, Y.-C., Tsai, S.-T., Hwang, S.-J., & Lee, S.-D. (2005). Hepatitis C virus infection-related Type 1 diabetes mellitus. Diabetic Medicine: A Journal of the British Diabetic Association, 22(3), 340–343.

Chen, M., Zhao, Y., Wang, Z., He, B., & Yao, J. (2023). A Noisy-Label-Learning Formulation for Immune Repertoire Classification and Disease-Associated Immune Receptor Sequence Identification. In arXiv [cs.LG]. arXiv. http://arxiv.org/abs/2307.15934

Chiffelle, J., Genolet, R., Perez, M. A., Coukos, G., Zoete, V., & Harari, A. (2020). T-cell repertoire analysis and metrics of diversity and clonality. Current Opinion in Biotechnology, 65, 284–295.

Chotisorayuth, T., & Tiffeau-Mayer, A. (2024). Lightning-fast adaptive immune receptor similarity search by symmetric deletion lookup. In arXiv [q-bio.QM]. arXiv. http://arxiv.org/abs/2403.09010

Chow, I.-T., Gates, T. J., Papadopoulos, G. K., Moustakas, A. K., Kolawole, E. M., Notturno, R. J., McGinty, J. W., Torres-Chinn, N., James, E. A., Greenbaum, C., Nepom, G. T., Evavold, B. D., & Kwok, W. W. (2019). Discriminative T cell recognition of cross-reactive islet-antigens is associated with HLA-DQ8 transdimer-mediated autoimmune diabetes. Science Advances, 5(8), eaaw9336.

Christley, S., Aguiar, A., Blanck, G., Breden, F., Bukhari, S. A. C., Busse, C. E., Jaglale, J., Harikrishnan, S. L., Laserson, U., Peters, B., Rocha, A., Schramm, C. A., Taylor, S., Vander Heiden, J. A., Zimonja, B., Watson, C. T., Corrie, B., & Cowell, L. G. (2020). The ADC API: A web API for the programmatic query of the AIRR Data Commons. Frontiers in Big Data, 3, 22.

Christophersen, A., Lund, E. G., Snir, O., Solà, E., Kanduri, C., Dahal-Koirala, S., Zühlke, S., Molberg, Ø., Utz, P. J., Rohani-Pichavant, M., Simard, J. F., Dekker, C. L., Lundin, K. E. A., Sollid, L. M., & Davis, M. M. (2019). Distinct phenotype of CD4+ T cells driving celiac disease identified in multiple autoimmune conditions. Nature Medicine, 25(5), 734–737.

Christophersen, A., Ráki, M., Bergseng, E., Lundin, K. E., Jahnsen, J., Sollid, L. M., & Qiao, S.-W. (2014). Tetramer-visualized gluten-specific CD4+ T cells in blood as a potential diagnostic marker for coeliac disease without oral gluten challenge. United European Gastroenterology Journal, 2(4), 268–278.

Chu, N. D., Bi, H. S., Emerson, R. O., Sherwood, A. M., Birnbaum, M. E., Robins, H. S., & Alm, E. J. (2019). Longitudinal immunosequencing in healthy people reveals persistent T cell receptors rich in highly public receptors. BMC Immunology, 20(1), 19.

Cohn, A., Sofia, A. M., & Kupfer, S. S. (2014). Type 1 diabetes and celiac disease: clinical overlap and new insights into disease pathogenesis. Current Diabetes Reports, 14(8), 517.

Conrad, N., Misra, S., Verbakel, J. Y., Verbeke, G., Molenberghs, G., Taylor, P. N., Mason, J., Sattar, N., McMurray, J. J. V., McInnes, I. B., Khunti, K., & Cambridge, G. (2023). Incidence, prevalence, and co-occurrence of autoimmune disorders over time and by age, sex, and socioeconomic status: a population-based cohort study of 22 million individuals in the UK. The Lancet, 401(10391), 1878– 1890.

Corrie, B. D., Marthandan, N., Zimonja, B., Jaglale, J., Zhou, Y., Barr, E., Knoetze, N., Breden, F. M. W., Christley, S., Scott, J. K., Cowell, L. G., & Breden, F. (2018). iReceptor: A platform for querying and analyzing antibody/B-cell and T-cell receptor repertoire data across federated repositories. Immunological Reviews, 284(1), 24–41.

Culina, S., Lalanne, A. I., Afonso, G., Cerosaletti, K., Pinto, S., Sebastiani, G., Kuranda, K., Nigi, L., Eugster, A., Østerbye, T., Maugein, A., McLaren, J. E., Ladell, K., Larger, E., Beressi, J.-P., Lissina, A., Appay, V., Davidson, H. W., Buus, S., … ImMaDiab Study Group. (2018). Islet-reactive CD8^+^ T cell frequencies in the pancreas, but not in blood, distinguish type 1 diabetic patients from healthy donors. Science Immunology, *3*(20). 10.1126/sciimmunol.aao4013

Dahal-Koirala, S., Balaban, G., Neumann, R. S., Scheffer, L., Lundin, K. E. A., Greiff, V., Sollid, L. M., Qiao, S.-W., & Sandve, G. K. (2022). TCRpower: quantifying the detection power of T-cell receptor sequencing with a novel computational pipeline calibrated by spike-in sequences. Briefings in Bioinformatics, 23(2), bbab566.

Davis, A. K., DuBose, S. N., Haller, M. J., Miller, K. M., DiMeglio, L. A., Bethin, K. E., Goland, R. S., Greenberg, E. M., Liljenquist, D. R., Ahmann, A. J., Marcovina, S. M., Peters, A. L., Beck, R. W., Greenbaum, C. J., & T1D Exchange Clinic Network. (2015). Prevalence of detectable C-Peptide according to age at diagnosis and duration of type 1 diabetes. Diabetes Care, *38*(3), 476–481.

De Neuter, N., Bartholomeus, E., Elias, G., Keersmaekers, N., Suls, A., Jansens, H., Smits, E., Hens, N., Beutels, P., Van Damme, P., Mortier, G., Van Tendeloo, V., Laukens, K., Meysman, P., & Ogunjimi, B. (2019). Memory CD4+ T cell receptor repertoire data mining as a tool for identifying cytomegalovirus serostatus. Genes and Immunity, 20(3), 255–260.

Denzin, L. K. (2013). Inhibition of HLA-DM mediated MHC class II peptide loading by HLA-DO promotes self tolerance. Frontiers in Immunology, 4, 465.

DeWitt, W. S., 3rd, Smith, A., Schoch, G., Hansen, J. A., Matsen, F. A., 4th, & Bradley, P. (2018). Human T cell receptor occurrence patterns encode immune history, genetic background, and receptor specificity. eLife, *7*. 10.7554/eLife.38358

Dilthey, A., Leslie, S., Moutsianas, L., Shen, J., Cox, C., Nelson, M. R., & McVean, G. (2013). Multi-population classical HLA type imputation. PLoS Computational Biology, 9(2), e1002877.

Ekman, I., Vuorinen, T., Knip, M., Veijola, R., Toppari, J., Hyöty, H., Kinnunen, T., Ilonen, J., & Lempainen, J. (2019). Early childhood CMV infection may decelerate the progression to clinical type 1 diabetes. Pediatric Diabetes, 20(1), 73–77.

ElAbd, H., Mahdy, A., Wacker, E. M., Gretsova, M., Ellinghaus, D., & Franke, A. (2025). Decoding the restriction of T cell receptors to human leukocyte antigen alleles using statistical learning. In bioRxiv (p. 2025.02.06.636910). 10.1101/2025.02.06.636910

Emerson, R. O., DeWitt, W. S., Vignali, M., Gravley, J., Hu, J. K., Osborne, E. J., Desmarais, C., Klinger, M., Carlson, C. S., Hansen, J. A., Rieder, M., & Robins, H. S. (2017). Immunosequencing identifies signatures of cytomegalovirus exposure history and HLA-mediated effects on the T cell repertoire. Nature Genetics, 49, 659–665.

Estorninho, M., Gibson, V. B., Kronenberg-Versteeg, D., Liu, Y.-F., Ni, C., Cerosaletti, K., & Peakman, M. (2013). A novel approach to tracking antigen-experienced CD4 T cells into functional compartments via tandem deep and shallow TCR clonotyping. Journal of Immunology , 191(11), 5430–5440.

Eugster, A., Lindner, A., Catani, M., Heninger, A.-K., Dahl, A., Klemroth, S., Kühn, D., Dietz, S., Bickle, M., Ziegler, A.-G., & Bonifacio, E. (2015). High diversity in the TCR repertoire of GAD65 autoantigen-specific human CD4+ T cells. The Journal of Immunology, 194(6), 2531–2538.

Eugster, A., Lorenc, A., Kotrulev, M., Kamra, Y., Goel, M., Steinberg-Bains, K., Sabbah, S., Dietz, S., Bonifacio, E., Peakman, M., & Gomez-Tourino, I. (2024). Physiological and pathogenic T cell autoreactivity converge in type 1 diabetes. Nature Communications, 15(1), 9204.

Ferreira, R. C., Simons, H. Z., Thompson, W. S., Cutler, A. J., Dopico, X. C., Smyth, D. J., Mashar, M., Schuilenburg, H., Walker, N. M., Dunger, D. B., Wallace, C., Todd, J. A., Wicker, L. S., & Pekalski, M. L. (2015). IL-21 production by CD4+ effector T cells and frequency of circulating follicular helper T cells are increased in type 1 diabetes patients. Diabetologia, 58(4), 781–790.

Frith, M. C., Saunders, N. F. W., Kobe, B., & Bailey, T. L. (2008). Discovering sequence motifs with arbitrary insertions and deletions. PLoS Computational Biology, 4(4), e1000071.

García, A. R., Paterou, A., Lee, M., Sławiński, H., Ferreira, R., Landry, L. G., Trzupek, D., Teyton, L., Szypowska, A., Wicker, L. S., Nakayama, M., Todd, J. A., & Pękalski, M. Ł. (2021). HLA class II mediates type 1 diabetes risk by anti-insulin repertoire selection. In bioRxiv (p. 2021.09.06.458974). 10.1101/2021.09.06.458974

García, A. R., Paterou, A., Lee, M., Sławiński, H., Wicker, L. S., Todd, J. A., & Pękalski, M. Ł. (2019). Peripheral tolerance to insulin is encoded by mimicry in the microbiome. In bioRxiv (p. 2019.12.18.881433). bioRxiv. 10.1101/2019.12.18.881433

García, A. R., Paterou, A., Powell Doherty, R. D., Landry, L. G., Lee, M., Anderson, A. M., Scudder, C. L., Slawinski, H., Ferreira, R. C., Trzupek, D., Szypowska, A., Teyton, L., Ternette, N., Nakayama, M., Wicker, L. S., Todd, J. A., & Pekalski, M. L. (2022). Autoimmune interactions between the HLA-DQβ1_57_polymorphism, T cell receptors, and microbial mimics of insulin in type 1 diabetes. In medRxiv (p. 2022.05.11.22274678). 10.1101/2022.05.11.22274678

Gitelman, S. E., & Bluestone, J. A. (2016). Regulatory T cell therapy for type 1 diabetes: May the force be with you. Journal of Autoimmunity, 71, 78–87.

Glanville, J., Huang, H., Nau, A., Hatton, O., Wagar, L. E., Rubelt, F., Ji, X., Han, A., Krams, S. M., Pettus, C., Haas, N., Arlehamn, C. S. L., Sette, A., Boyd, S. D., Scriba, T. J., Martinez, O. M., & Davis, M. M. (2017). Identifying specificity groups in the T cell receptor repertoire. Nature, 547(7661), 94–98.

Gomes, K. F. B., Santos, A. S., Semzezem, C., Correia, M. R., Brito, L. A., Ruiz, M. O., Fukui, R. T., Matioli, S. R., Passos-Bueno, M. R., & Silva, M. E. R. da. (2017). The influence of population stratification on genetic markers associated with type 1 diabetes. Scientific Reports, 7, 43513.

Gomez-Tourino, I., Kamra, Y., Baptista, R., Lorenc, A., & Peakman, M. (2017). T cell receptor β-chains display abnormal shortening and repertoire sharing in type 1 diabetes. Nature Communications, 8(1), 1792.

Gotelli, N. J., & Colwell, R. K. (2001). Quantifying biodiversity: procedures and pitfalls in the measurement and comparison of species richness. Ecology Letters, 4(4), 379–391.

Greiff, V., Bhat, P., Cook, S. C., Menzel, U., Kang, W., & Reddy, S. T. (2015). A bioinformatic framework for immune repertoire diversity profiling enables detection of immunological status. Genome Medicine, 7(1), 49.

Greiff, V., Menzel, U., Haessler, U., Cook, S. C., Friedensohn, S., Khan, T. A., Pogson, M., Hellmann, I., & Reddy, S. T. (2014). Quantitative assessment of the robustness of next-generation sequencing of antibody variable gene repertoires from immunized mice. BMC Immunology, 15, 40.

Greiff, V., Weber, C. R., Palme, J., Bodenhofer, U., Miho, E., Menzel, U., & Reddy, S. T. (2017). Learning the high-dimensional immunogenomic features that predict public and private antibody repertoires. The Journal of Immunology, 199(8), 2985–2997.

Greiff, V., Yaari, G., & Cowell, L. (2020). Mining adaptive immune receptor repertoires for biological and clinical information using machine learning. Current Opinion in Systems Biology. 10.1016/j.coisb.2020.10.010

Greissl, J., Pesesky, M., Dalai, S. C., Rebman, A. W., Soloski, M. J., Horn, E. J., Dines, J. N., Gittelman, R. M., Snyder, T. M., Emerson, R. O., Meeds, E., Manley, T., Kaplan, I. M., Baldo, L., Carlson, J. M., Robins, H. S., & Aucott, J. N. (2021). Immunosequencing of the T-cell receptor repertoire reveals signatures specific for diagnosis and characterization of early Lyme disease. In bioRxiv. medRxiv. 10.1101/2021.07.30.21261353

Gutierrez-Achury, J., Romanos, J., Bakker, S. F., Kumar, V., de Haas, E. C., Trynka, G., Ricaño-Ponce, I., Steck, A., Type 1 Diabetes Genetics Consortium, Chen, W.-M., Onengut-Gumuscu, S., Simsek, S., Diabeter, Rewers, M., Mulder, C. J., Liu, E., Rich, S. S., & Wijmenga, C. (2015). Contrasting the Genetic Background of Type 1 Diabetes and Celiac Disease Autoimmunity. Diabetes Care, *38 Suppl 2*(Suppl 2), S37–S44.

Hagberg, A., Schult, D., Swart, P., & Hagberg, J. M. (2008). Exploring Network Structure, Dynamics, and Function using NetworkX. https://www.semanticscholar.org/paper/06214a0cf38875da38586e81539890f7ad8aeb1c

Hanna, S. J., Bonami, R. H., Corrie, B., Westley, M., Posgai, A. L., Luning Prak, E. T., Breden, F., Michels, A. W., Brusko, T. M., & Type 1 Diabetes AIRR Consortium. (2024). The Type 1 Diabetes T Cell Receptor and B Cell Receptor Repository in the AIRR Data Commons: a practical guide for access, use and contributions through the Type 1 Diabetes AIRR Consortium. Diabetologia, 1–17.

Hill, M. O. (1973). Diversity and evenness: A unifying notation and its consequences. Ecology, 54(2), 427–432.

Horn, H. S. (1966). Measurement of “overlap” in comparative ecological studies. The American Naturalist, 100(914), 419–424.

Hu, X., Deutsch, A. J., Lenz, T. L., Onengut-Gumuscu, S., Han, B., Chen, W.-M., Howson, J. M. M., Todd, J. A., de Bakker, P. I. W., Rich, S. S., & Raychaudhuri, S. (2015). Additive and interaction effects at three amino acid positions in HLA-DQ and HLA-DR molecules drive type 1 diabetes risk. Nature Genetics, 47(8), 898–905.

Ilonen, J., Sjöroos, M., Knip, M., Veijola, R., Simell, O., Akerblom, H. K., Paschou, P., Bozas, E., Havarani, B., Malamitsi-Puchner, A., Thymelli, J., Vazeou, A., & Bartsocas, C. S. (2002). Estimation of genetic risk for type 1 diabetes. American Journal of Medical Genetics, 115(1), 30–36.

Isacchini, G., Walczak, A. M., Mora, T., & Nourmohammad, A. (2021). Deep generative selection models of T and B cell receptor repertoires with soNNia. Proceedings of the National Academy of Sciences of the United States of America, 118(14), e2023141118.

Ishigaki, K., Lagattuta, K. A., Luo, Y., James, E. A., Buckner, J. H., & Raychaudhuri, S. (2022). HLA autoimmune risk alleles restrict the hypervariable region of T cell receptors. Nature Genetics, 54(4), 393–402.

Jacobsen, L. M., Posgai, A., Seay, H. R., Haller, M. J., & Brusko, T. M. (2017). T cell receptor profiling in type 1 diabetes. Current Diabetes Reports, 17(11), 118.

Jia, X., Han, B., Onengut-Gumuscu, S., Chen, W.-M., Concannon, P. J., Rich, S. S., Raychaudhuri, S., & de Bakker, P. I. W. (2013). Imputing amino acid polymorphisms in human leukocyte antigens. PloS One, 8(6), e64683.

Joglekar, A. V., & Li, G. (2021). T cell antigen discovery. Nature Methods, 18(8), 873–880.

Jost, L. (2006). Entropy and diversity. Oikos , *113*(2), 363–375.

Katayama, Y., & Kobayashi, T. J. (2022). Comparative study of repertoire classification methods reveals data efficiency of k -mer feature extraction. Frontiers in Immunology, 13, 797640.

Kendall, E. K., Olaker, V. R., Kaelber, D. C., Xu, R., & Davis, P. B. (2022). Association of SARS-CoV-2 Infection With New-Onset Type 1 Diabetes Among Pediatric Patients From 2020 to 2021. JAMA Network Open, 5(9), e2233014.

Komech, E. A., Pogorelyy, M. V., Egorov, E. S., Britanova, O. V., Rebrikov, D. V., Bochkova, A. G., Shmidt, E. I., Shostak, N. A., Shugay, M., Lukyanov, S., Mamedov, I. Z., Lebedev, Y. B., Chudakov, D. M., & Zvyagin, I. V. (2018). CD8+ T cells with characteristic T cell receptor beta motif are detected in blood and expanded in synovial fluid of ankylosing spondylitis patients. Rheumatology , 57(6), 1097–1104.

Krischer, J. P., Liu, X., Lernmark, Å., Hagopian, W. A., Rewers, M. J., She, J.-X., Toppari, J., Ziegler, A.-G., Akolkar, B., & TEDDY Study Group. (2022). Predictors of the Initiation of Islet Autoimmunity and Progression to Multiple Autoantibodies and Clinical Diabetes: The TEDDY Study. Diabetes Care, 45(10), 2271–2281.

Krishna, C., Chowell, D., Gönen, M., Elhanati, Y., & Chan, T. A. (2020). Genetic and environmental determinants of human TCR repertoire diversity. Immunity & Ageing: I & A, 17(1), 26.

Krogvold, L., Edwin, B., Buanes, T., Frisk, G., Skog, O., Anagandula, M., Korsgren, O., Undlien, D., Eike, M. C., Richardson, S. J., Leete, P., Morgan, N. G., Oikarinen, S., Oikarinen, M., Laiho, J. E., Hyöty, H., Ludvigsson, J., Hanssen, K. F., & Dahl-Jørgensen, K. (2015). Detection of a low-grade enteroviral infection in the islets of langerhans of living patients newly diagnosed with type 1 diabetes. Diabetes, 64(5), 1682–1687.

Lagattuta, K. A., Kang, J. B., Nathan, A., Pauken, K. E., Jonsson, A. H., Rao, D. A., Sharpe, A. H., Ishigaki, K., & Raychaudhuri, S. (2022). Repertoire analyses reveal T cell antigen receptor sequence features that influence T cell fate. Nature Immunology, 23(3), 446–457.

Lambert, A. P., Gillespie, K. M., Thomson, G., Cordell, H. J., Todd, J. A., Gale, E. A. M., & Bingley, P. J. (2004). Absolute risk of childhood-onset type 1 diabetes defined by human leukocyte antigen class II genotype: a population-based study in the United Kingdom. The Journal of Clinical Endocrinology and Metabolism, 89(8), 4037–4043.

Lefranc, M.-P., Giudicelli, V., Ginestoux, C., Jabado-Michaloud, J., Folch, G., Bellahcene, F., Wu, Y., Gemrot, E., Brochet, X., Lane, J., Regnier, L., Ehrenmann, F., Lefranc, G., & Duroux, P. (2009). IMGT, the international ImMunoGeneTics information system. Nucleic Acids Research, 37(Database issue), D1006–D1012.

Liu, M., Goo, J., Liu, Y., Sun, W., Wu, M. C., Hsu, L., & He, Q. (2022). TCR-L: an analysis tool for evaluating the association between the T-cell receptor repertoire and clinical phenotypes. BMC Bioinformatics, 23(1), 152.

Liu, S., Bradley, P., & Sun, W. (2023a). Neural network models for sequence-based TCR and HLA association prediction. PLoS Computational Biology, 19(11), e1011664.

Liu, S., Bradley, P., & Sun, W. (2023b). Neural Network Models for Sequence-Based TCR and HLA Association Prediction. In bioRxiv (p. 2023.05.25.542327). 10.1101/2023.05.25.542327

Liu, X., Zhang, W., Zhao, M., Fu, L., Liu, L., Wu, J., Luo, S., Wang, L., Wang, Z., Lin, L., Liu, Y., Wang, S., Yang, Y., Luo, L., Jiang, J., Wang, X., Tan, Y., Li, T., Zhu, B., … Lu, Q. (2019). T cell receptor β repertoires as novel diagnostic markers for systemic lupus erythematosus and rheumatoid arthritis. Annals of the Rheumatic Diseases, 78(8), 1070–1078.

Machiela, M. J., & Chanock, S. J. (2015). LDlink: a web-based application for exploring population-specific haplotype structure and linking correlated alleles of possible functional variants. *Bioinformatics (Oxford*, England), 31(21), 3555–3557.

Majumder, P., Lee, J. T., Rahmberg, A. R., Kumar, G., Mi, T., Scharer, C. D., & Boss, J. M. (2020). A super enhancer controls expression and chromatin architecture within the MHC class II locus. The Journal of Experimental Medicine, 217(2), jem.20190668.

Ma, K.-Y., Schonnesen, A. A., He, C., Xia, A. Y., Sun, E., Chen, E., Sebastian, K. R., Guo, Y.-W., Balderas, R., Kulkarni-Date, M., & Jiang, N. (2021). High-throughput and high-dimensional single-cell analysis of antigen-specific CD8+ T cells. Nature Immunology, 22(12), 1590–1598.

Marcou, Q., Mora, T., & Walczak, A. M. (2018). High-throughput immune repertoire analysis with IGoR. Nature Communications, 9(1), 561.

Marzinotto, I., Pittman, D. L., Williams, A. J. K., Long, A. E., Achenbach, P., Schlosser, M., Akolkar, B., Winter, W. E., Lampasona, V., & participating laboratories. (2023). Islet Autoantibody Standardization Program: interlaboratory comparison of insulin autoantibody assay performance in 2018 and 2020 workshops. Diabetologia, 66(5), 897–912.

Masuda, H., Atsumi, T., Fujisaku, A., Shimizu, C., Yoshioka, N., & Koike, T. (2007). Acute onset of type 1 diabetes accompanied by acute hepatitis C: the potential role of proinflammatory cytokine in the pathogenesis of autoimmune diabetes. Diabetes Research and Clinical Practice, 75(3), 357–361.

May, D. H., Woodhouse, S., Howie, B., & Robins, H. S. (2024). A Catalog of the Public T-cell Response to Cytomegalovirus. In bioRxiv (p. 2024.05.08.593237). 10.1101/2024.05.08.593237

Mayer-Blackwell, K., Schattgen, S., Cohen-Lavi, L., Crawford, J. C., Souquette, A., Gaevert, J. A., Hertz, T., Thomas, P. G., Bradley, P., & Fiore-Gartland, A. (2021). TCR meta-clonotypes for biomarker discovery with tcrdist3 enabled identification of public, HLA-restricted clusters of SARS-CoV-2 TCRs. eLife, *10*. 10.7554/eLife.68605

Mhanna, V., Bashour, H., Lê Quý, K., Barennes, P., Rawat, P., Greiff, V., & Mariotti-Ferrandiz, E. (2024). Adaptive immune receptor repertoire analysis. Nature Reviews Methods Primers, 4(1), 1–25.

Mhanna, V., Fourcade, G., Barennes, P., Quiniou, V., Pham, H. P., Ritvo, P.-G., Brimaud, F., Gouritin, B., Churlaud, G., Six, A., Mariotti-Ferrandiz, E., & Klatzmann, D. (2021). Impaired activated/memory regulatory T cell clonal expansion instigates diabetes in NOD mice. Diabetes, 70(4), 976–985.

Miho, E., Yermanos, A., Weber, C. R., Berger, C. T., Reddy, S. T., & Greiff, V. (2018). Computational Strategies for Dissecting the High-Dimensional Complexity of Adaptive Immune Repertoires. Frontiers in Immunology, 9, 224.

Min-ChunYeh, Chuang, H.-C., Weng, S.-F., Hsu, C.-H., Huang, C.-L., Lin, Y.-P., Lin, Y.-Y., & Hsieh, Y.-S. (2023). Newly diagnosed type 1 diabetes mellitus in a human immunodeficiency virus-infected patient with antiretroviral therapy-induced immune reconstitution inflammatory syndrome: a case report. BMC Infectious Diseases, 23(1), 619.

Minervina, A., Pogorelyy, M., & Mamedov, I. (2019). T-cell receptor and B-cell receptor repertoire profiling in adaptive immunity. Transplant International: Official Journal of the European Society for Organ Transplantation, 32(11), 1111–1123.

Mitchell, A. M., Baschal, E. E., McDaniel, K. A., Fleury, T., Choi, H., Pyle, L., Yu, L., Rewers, M. J., Nakayama, M., & Michels, A. W. (2023). Tracking DNA-based antigen-specific T cell receptors during progression to type 1 diabetes. Science Advances, 9(49), eadj6975.

Mitchell, A. M., Baschal, E. E., McDaniel, K. A., Simmons, K. M., Pyle, L., Waugh, K., Steck, A. K., Yu, L., Gottlieb, P. A., Rewers, M. J., Nakayama, M., & Michels, A. W. (2022). Temporal development of T cell receptor repertoires during childhood in health and disease. JCI Insight, 7(18). 10.1172/jci.insight.161885

Mitchell, A. M., & Michels, A. W. (2020). T cell receptor sequencing in autoimmunity. *Journal of Life Sciences (Westlake Village*, Calif.), 2(4), 38–58.

Nagafuchi, Y., Nakano, M., Lagattuta, K. A., Ota, M., Hatano, H., Takahashi, H., Itamiya, T., Inokuchi, H., Raychaudhuri, S., Okamura, T., Fujio, K., & Ishigaki, K. (2025). T cell plasticity in systemic lupus erythematosus revealed by large-scale T cell receptor repertoire and transcriptome studies. In medRxiv (p. 2025.01.06.24319648). 10.1101/2025.01.06.24319648

Nagafuchi, Y., Ota, M., Hatano, H., Inoue, M., Kobayashi, S., Okubo, M., Sugimori, Y., Nakano, M., Yamada, S., Yoshida, R., Tsuchida, Y., Iwasaki, Y., Shoda, H., Okada, Y., Yamamoto, K., Ishigaki, K., Okamura, T., & Fujio, K. (2022). Control of naive and effector CD4 T cell receptor repertoires by rheumatoid-arthritis-risk HLA alleles. Journal of Autoimmunity, 133, 102907.

Naik, R. G., Beckers, C., Wentwoord, R., Frenken, A., Duinkerken, G., Brooks-Worrell, B., Schloot, N. C., Palmer, J. P., & Roep, B. O. (2004). Precursor frequencies of T-cells reactive to insulin in recent onset type 1 diabetes mellitus. Journal of Autoimmunity, 23(1), 55–61.

Nakayama, M., Abiru, N., Moriyama, H., Babaya, N., Liu, E., Miao, D., Yu, L., Wegmann, D. R., Hutton, J. C., Elliott, J. F., & Eisenbarth, G. S. (2005). Prime role for an insulin epitope in the development of type 1 diabetes in NOD mice. Nature, 435(7039), 220–223.

Nakayama, M., & Michels, A. W. (2021). Using the T cell receptor as a biomarker in type 1 diabetes. Frontiers in Immunology, 12, 777788.

Niu, J., Jia, Q., Ni, Q., Yang, Y., Chen, G., Yang, X., Zhai, Z., Yu, H., Guan, P., Lin, R., Song, Z., Li, Q.- J., Hao, F., Zhong, H., & Wan, Y. (2015). Association of CD8(+) T lymphocyte repertoire spreading with the severity of DRESS syndrome. Scientific Reports, 5, 9913.

Noble, J. A., & Erlich, H. A. (2012). Genetics of type 1 diabetes. Cold Spring Harbor Perspectives in Medicine, 2(1), a007732.

Noble, J. A., & Valdes, A. M. (2011). Genetics of the HLA region in the prediction of type 1 diabetes. Current Diabetes Reports, 11(6), 533–542.

Noble, J. A., Valdes, A. M., Cook, M., Klitz, W., Thomson, G., & Erlich, H. A. (1996). The role of HLA class II genes in insulin-dependent diabetes mellitus: molecular analysis of 180 Caucasian, multiplex families. American Journal of Human Genetics, 59(5), 1134–1148.

O’Donnell, T. J., Kanduri, C., Isacchini, G., Limenitakis, J. P., Brachman, R. A., Alvarez, R. A., Haff, I. H., Sandve, G. K., & Greiff, V. (2024). Reading the repertoire: Progress in adaptive immune receptor analysis using machine learning. Cell Systems, 15(12), 1168–1189.

Ortega, M. R., Pogorelyy, M. V., Minervina, A. A., Thomas, P. G., Walczak, A. M., & Mora, T. (2024). Learning predictive signatures of HLA type from T-cell repertoires. bioRxiv.org: The Preprint Server for Biology, 2024.01.25.577228.

Ostmeyer, J., Christley, S., Toby, I. T., & Cowell, L. G. (2019). Biophysicochemical motifs in T cell receptor sequences distinguish repertoires from tumor-infiltrating lymphocytes and adjacent healthy tissue. Cancer Research. 10.1158/0008-5472.CAN-18-2292

Pak, C. Y., Eun, H. M., McArthur, R. G., & Yoon, J. W. (1988). Association of cytomegalovirus infection with autoimmune type 1 diabetes. The Lancet, 2(8601), 1–4.

Pathiraja, V., Kuehlich, J. P., Campbell, P. D., Krishnamurthy, B., Loudovaris, T., Coates, P. T. H., Brodnicki, T. C., O’Connell, P. J., Kedzierska, K., Rodda, C., Bergman, P., Hill, E., Purcell, A. W., Dudek, N. L., Thomas, H. E., Kay, T. W. H., & Mannering, S. I. (2015). Proinsulin-specific, HLA-DQ8, and HLA-DQ8-transdimer-restricted CD4+ T cells infiltrate islets in type 1 diabetes. Diabetes, *64*(1), 172–182.

Pavlović, M., Scheffer, L., Motwani, K., Kanduri, C., Kompova, R., Vazov, N., Waagan, K., Bernal, F. L. M., Costa, A. A., Corrie, B., Akbar, R., Al Hajj, G. S., Balaban, G., Brusko, T. M., Chernigovskaya, M., Christley, S., Cowell, L. G., Frank, R., Grytten, I., … Sandve, G. K. (2021). The immuneML ecosystem for machine learning analysis of adaptive immune receptor repertoires. Nature Machine Intelligence, 3(11), 936–944.

Perry, D. J., Shapiro, M. R., Chamberlain, S. W., Kusmartseva, I., Chamala, S., Balzano-Nogueira, L., Yang, M., Brant, J. O., Brusko, M., Williams, M. D., McGrail, K. M., McNichols, J., Peters, L. D., Posgai, A. L., Kaddis, J. S., Mathews, C. E., Wasserfall, C. H., Webb-Robertson, B.-J. M., Campbell-Thompson, M., … Brusko, T. M. (2023). A genomic data archive from the Network for Pancreatic Organ donors with Diabetes. Scientific Data, 10(1), 323.

Pociot, F., & Lernmark, Å. (2016). Genetic risk factors for type 1 diabetes. The Lancet, 387(10035), 2331–2339.

Pogorelyy, M. V., Kirk, A. M., Adhikari, S., Minervina, A. A., Sundararaman, B., Vegesana, K., Brice, D. C., Scott, Z. B., & Thomas, P. G. (2024). TIRTL-seq: Deep, quantitative, and affordable paired TCR repertoire sequencing. In Immunology (No. biorxiv;2024.09.16.613345v1). bioRxiv. https://www.biorxiv.org/content/10.1101/2024.09.16.613345v1

Pugliese, A. (2017). Autoreactive T cells in type 1 diabetes. The Journal of Clinical Investigation, 127(8), 2881–2891.

Ramsauer, H., Schäfl, B., Lehner, J., Seidl, P., Widrich, M., Adler, T., Gruber, L., Holzleitner, M., Pavlović, M., Sandve, G. K., Greiff, V., Kreil, D., Kopp, M., Klambauer, G., Brandstetter, J., & Hochreiter, S. (2020). Hopfield Networks is All You Need. In arXiv [cs.NE]. arXiv. http://arxiv.org/abs/2008.02217

Rényi, A. (1961). On measures of entropy and information. Proceedings of the Fourth Berkeley Symposium on Mathematical Statistics and Probability, Volume 1: Contributions to the Theory of Statistics, *4*, 547–562.

Robertson, C. C., Inshaw, J. R. J., Onengut-Gumuscu, S., Chen, W.-M., Santa Cruz, D. F., Yang, H., Cutler, A. J., Crouch, D. J. M., Farber, E., Bridges, S. L., Jr, Edberg, J. C., Kimberly, R. P., Buckner, J. H., Deloukas, P., Divers, J., Dabelea, D., Lawrence, J. M., Marcovina, S., Shah, A. S., … Rich, S. S. (2021). Fine-mapping, trans-ancestral and genomic analyses identify causal variants, cells, genes and drug targets for type 1 diabetes. Nature Genetics, 53(7), 962–971.

Robins, H. S., Campregher, P. V., Srivastava, S. K., Wacher, A., Turtle, C. J., Kahsai, O., Riddell, S. R., Warren, E. H., & Carlson, C. S. (2009). Comprehensive assessment of T-cell receptor beta-chain diversity in alphabeta T cells. Blood, 114(19), 4099–4107.

Rognes, T., Scheffer, L., Greiff, V., & Sandve, G. K. (2022). CompAIRR: ultra-fast comparison of adaptive immune receptor repertoires by exact and approximate sequence matching. Bioinformatics , 38(17), 4230–4232.

Ross, J. J., Wasserfall, C. H., Bacher, R., Perry, D. J., McGrail, K., Posgai, A. L., Dong, X., Muir, A., Li, X., Campbell-Thompson, M., Brusko, T. M., Schatz, D. A., Haller, M. J., & Atkinson, M. A. (2021). Exocrine pancreatic enzymes are a serological biomarker for type 1 diabetes staging and pancreas size. Diabetes, 70(4), 944–954.

Sakaue, S., Gurajala, S., Curtis, M., Luo, Y., Choi, W., Ishigaki, K., Kang, J. B., Rumker, L., Deutsch, A. J., Schönherr, S., Forer, L., LeFaive, J., Fuchsberger, C., Han, B., Lenz, T. L., de Bakker, P. I. W., Okada, Y., Smith, A. V., & Raychaudhuri, S. (2023). Tutorial: a statistical genetics guide to identifying HLA alleles driving complex disease. Nature Protocols. 10.1038/s41596-023-00853-4

Schmidt-Barbo, P., Kalweit, G., Naouar, M., Paschold, L., Willscher, E., Schultheiß, C., Märkl, B., Dirnhofer, S., Tzankov, A., Binder, M., & Kalweit, M. (2024). Detection of disease-specific signatures in B cell repertoires of lymphomas using machine learning. PLoS Computational Biology, 20(7), e1011570.

Schneider-Hohendorf, T., Wünsch, C., Falk, S., Raposo, C., Rubelt, F., Mirebrahim, H., Asgharian, H., Schlecht, U., Mattox, D., Zhou, W., Dawin, E., Pawlitzki, M., Lauks, S., Jarius, S., Wildemann, B., Havla, J., Kümpfel, T., Schrot, M.-C., Ringelstein, M., … Schwab, N. (2024). Broader anti-EBV TCR repertoire in multiple sclerosis: disease specificity and treatment modulation. *Brain: A Journal of Neurology*, awae244.

Seay, H. R., Yusko, E., Rothweiler, S. J., Zhang, L., Posgai, A. L., Campbell-Thompson, M., Vignali, M., Emerson, R. O., Kaddis, J. S., Ko, D., Nakayama, M., Smith, M. J., Cambier, J. C., Pugliese, A., Atkinson, M. A., Robins, H. S., & Brusko, T. M. (2016). Tissue distribution and clonal diversity of the T and B cell repertoire in type 1 diabetes. JCI Insight, 1(20), e88242.

Shannon, C. E. (1948). A mathematical theory of communication. The Bell System Technical Journal, 27(3), 379–423.

Shapiro, M. R., Dong, X., Perry, D. J., McNichols, J. M., Thirawatananond, P., Posgai, A. L., Peters, L. D., Motwani, K., Musca, R. S., Muir, A., Concannon, P., Jacobsen, L. M., Mathews, C. E., Wasserfall, C. H., Haller, M. J., Schatz, D. A., Atkinson, M. A., Brusko, M. A., Bacher, R., & Brusko, T. M. (2023). Human immune phenotyping reveals accelerated aging in type 1 diabetes. JCI Insight, 8(17). 10.1172/jci.insight.170767

Shomuradova, A. S., Vagida, M. S., Sheetikov, S. A., Zornikova, K. V., Kiryukhin, D., Titov, A., Peshkova, I. O., Khmelevskaya, A., Dianov, D. V., Malasheva, M., Shmelev, A., Serdyuk, Y., Bagaev, D. V., Pivnyuk, A., Shcherbinin, D. S., Maleeva, A. V., Shakirova, N. T., Pilunov, A., Malko, D. B., … Efimov, G. A. (2020). SARS-CoV-2 epitopes are recognized by a public and diverse repertoire of human T cell receptors. Immunity, 53(6), 1245–1257.e5.

Sidhom, J.-W., Benjamin Larman, H., Pardoll, D. M., & Baras, A. S. (2021). DeepTCR is a deep learning framework for revealing sequence concepts within T-cell repertoires. In Nature Communications (Vol. 12, Issue 1). 10.1038/s41467-021-21879-w

Sidhom, J.-W., Oliveira, G., Ross-MacDonald, P., Wind-Rotolo, M., Wu, C. J., Pardoll, D. M., & Baras, A. S. (2022). Deep learning reveals predictive sequence concepts within immune repertoires to immunotherapy. Science Advances, 8(37), eabq5089.

Simpson, E. H. (1949). Measurement of Diversity. Nature, 163(4148), 688–688.

Skowera, A., Ladell, K., McLaren, J. E., Dolton, G., Matthews, K. K., Gostick, E., Kronenberg-Versteeg, D., Eichmann, M., Knight, R. R., Heck, S., Powrie, J., Bingley, P. J., Dayan, C. M., Miles, J. J., Sewell, A. K., Price, D. A., & Peakman, M. (2015). β-cell-specific CD8 T cell phenotype in type 1 diabetes reflects chronic autoantigen exposure. Diabetes, 64(3), 916–925.

Slabodkin, A., Sollid, L. M., Sandve, G. K., Robert, P. A., & Greiff, V. (2023). Weakly supervised identification and generation of adaptive immune receptor sequences associated with immune disease status. In bioRxiv (p. 2023.09.24.558823). 10.1101/2023.09.24.558823

Smith, C. J., Strausz, S., FinnGen, Spence, J. P., Ollila, H. M., & Pritchard, J. K. (2024). Haplotype analysis reveals pleiotropic disease associations in the HLA region. medRxiv: The Preprint Server for Health Sciences, 2024.07.29.24311183.

Snyder, T. M., Gittelman, R. M., Klinger, M., May, D. H., Osborne, E. J., Taniguchi, R., Zahid, H. J., Kaplan, I. M., Dines, J. N., Noakes, M. T., Pandya, R., Chen, X., Elasady, S., Svejnoha, E., Ebert, P., Pesesky, M. W., De Almeida, P., O’Donnell, H., DeGottardi, Q., … Robins, H. S. (2020). Magnitude and Dynamics of the T-Cell Response to SARS-CoV-2 Infection at Both Individual and Population Levels. medRxiv : The Preprint Server for Health Sciences. 10.1101/2020.07.31.20165647

Sokal, R., & Michener, C. (1958). A statistical method for evaluating systematic relationships. University of Kansas Science Bulletin. https://www.semanticscholar.org/paper/A-statistical-method-for-evaluating-systematic-Sokal-Michener/0db093335bc3b9445fa5a1a5526d634921d7b59a?sort=relevance&queryString=upgma

Stadinski, B. D., Cleveland, S. B., Brehm, M. A., Greiner, D. L., Huseby, P. G., & Huseby, E. S. (2023). I-Ag7 β56/57 polymorphisms regulate non-cognate negative selection to CD4+ T cell orchestrators of type 1 diabetes. Nature Immunology, 24(4), 652–663.

Strom, T. B. (2009). Can childhood viral infection protect from type 1 diabetes? The Journal of Clinical Investigation, 119(6), 1458–1461.

Sundararajan, M., Taly, A., & Yan, Q. (2017). Axiomatic Attribution for Deep Networks. In D. Precup & Y. W. Teh (Eds.), Proceedings of the 34th International Conference on Machine Learning (Vol. 70, pp. 3319–3328). PMLR.

Taguchi, M., Ihana-Sugiyama, N., Shiojiri, D., Izumi, K., Kobayashi, M., Kodani, N., Bouchi, R., Ohsugi, M., Tanabe, A., Ueki, K., & Kajio, H. (2023). New-onset type 1 diabetes and Graves’ disease after antiretroviral therapy in a patient with human immunodeficiency virus infection. Journal of Diabetes Investigation, 14(3), 489–493.

Textor, J., Buytenhuijs, F., Rogers, D., Gauthier, È. M., Sultan, S., Wortel, I. M. N., Kalies, K., Fähnrich, A., Pagel, R., Melichar, H. J., Westermann, J., & Mandl, J. N. (2023). Machine learning analysis of the T cell receptor repertoire identifies sequence features of self-reactivity. Cell Systems, 14(12), 1059–1073.e5.

Tickotsky, N., Sagiv, T., Prilusky, J., Shifrut, E., & Friedman, N. (2017). McPAS-TCR: a manually curated catalogue of pathology-associated T cell receptor sequences. Bioinformatics , 33(18), 2924– 2929.

Tran, M. T., Lim, J. J., Loh, T. J., Mannering, S. I., Rossjohn, J., & Reid, H. H. (2024). A structural basis of T cell cross-reactivity to native and spliced self-antigens presented by HLA-DQ8. The Journal of Biological Chemistry, 300(9), 107612.

T, R. R., Demerdash, O. N. A., & Smith, J. C. (2024). TCR-H: explainable machine learning prediction of T-cell receptor epitope binding on unseen datasets. Frontiers in Immunology, 15, 1426173.

Valkiers, S., Van Houcke, M., Laukens, K., & Meysman, P. (2021). ClusTCR: a python interface for rapid clustering of large sets of CDR3 sequences with unknown antigen specificity. Bioinformatics , 37(24), 4865–4867.

van Lummel, M., van Veelen, P. A., Zaldumbide, A., de Ru, A., Janssen, G. M. C., Moustakas, A. K., Papadopoulos, G. K., Drijfhout, J. W., Roep, B. O., & Koning, F. (2012). Type 1 diabetes-associated HLA-DQ8 transdimer accommodates a unique peptide repertoire. The Journal of Biological Chemistry, 287(12), 9514–9524.

Volta, U., Tovoli, F., & Caio, G. (2011). Clinical and immunological features of celiac disease in patients with Type 1 diabetes mellitus. Expert Review of Gastroenterology & Hepatology, 5(4), 479–487.

Võsa, U., Claringbould, A., Westra, H.-J., Bonder, M. J., Deelen, P., Zeng, B., Kirsten, H., Saha, A., Kreuzhuber, R., Yazar, S., Brugge, H., Oelen, R., de Vries, D. H., van der Wijst, M. G. P., Kasela, S., Pervjakova, N., Alves, I., Favé, M.-J., Agbessi, M., … Franke, L. (2021). Large-scale cis- and trans-eQTL analyses identify thousands of genetic loci and polygenic scores that regulate blood gene expression. Nature Genetics, 53(9), 1300–1310.

Vujović, M., Marcatili, P., Chain, B., Kaplinsky, J., & Andresen, T. L. (2023). Signatures of T cell immunity revealed using sequence similarity with TCRDivER algorithm. Communications Biology, 6(1), 357.

Wasserfall, C., Montgomery, E., Yu, L., Michels, A., Gianani, R., Pugliese, A., Nierras, C., Kaddis, J. S., Schatz, D. A., Bonifacio, E., & Atkinson, M. A. (2016). Validation of a rapid type 1 diabetes autoantibody screening assay for community-based screening of organ donors to identify subjects at increased risk for the disease. Clinical and Experimental Immunology, 185(1), 33–41.

Weber, A., Pélissier, A., & Rodríguez Martínez, M. (2024). T-cell receptor binding prediction: A machine learning revolution. *Immunoinformatics (Amsterdam*, Netherlands), 15(100040), 100040.

Weber, C. R., Rubio, T., Wang, L., Zhang, W., Robert, P. A., Akbar, R., Snapkov, I., Wu, J., Kuijjer, M. L., Tarazona, S., Conesa, A., Sandve, G. K., Liu, X., Reddy, S. T., & Greiff, V. (2022). Reference-based comparison of adaptive immune receptor repertoires. Cell Reports Methods, 2(8), 100269.

Weber, D. A., Evavold, B. D., & Jensen, P. E. (1996). Enhanced dissociation of HLA-DR-bound peptides in the presence of HLA-DM. *Science (New York*, N.Y.), 274(5287), 618–620.

Widrich, M., Schäfl, B., Pavlović, M., Ramsauer, H., Gruber, L., Holzleitner, M., Brandstetter, J., Sandve, G. K., Greiff, V., Hochreiter, S., & Klambauer, G. (2020). Modern Hopfield networks and attention for immune repertoire classification. In bioRxiv (pp. 18832–18845). bioRxiv. 10.1101/2020.04.12.038158

Williams, C. L., Fareed, R., Mortimer, G. L. M., Aitken, R. J., Wilson, I. V., George, G., Gillespie, K. M., Williams, A. J. K., BOX Study Group, & Long, A. E. (2022). The longitudinal loss of islet autoantibody responses from diagnosis of type 1 diabetes occurs progressively over follow-up and is determined by low autoantibody titres, early-onset, and genetic variants. Clinical and Experimental Immunology, 210(2), 151–162.

Williams, M. D., Bacher, R., Perry, D. J., Grace, C. R., McGrail, K. M., Posgai, A. L., Muir, A., Chamala, S., Haller, M. J., Schatz, D. A., Brusko, T. M., Atkinson, M. A., & Wasserfall, C. H. (2021). Genetic Composition and Autoantibody Titers Model the Probability of Detecting C-Peptide Following Type 1 Diabetes Diagnosis. Diabetes, 70(4), 932–943.

Yu, X., Pan, M., Ye, J., Hathaway, C. A., Tworoger, S. S., Lea, J., & Li, B. (2024). Quantifiable TCR repertoire changes in prediagnostic blood specimens among patients with high-grade ovarian cancer. *Cell Reports*. Medicine, 5(7), 101612.

Zahid, H. J., Taniguchi, R., Ebert, P., Chow, I.-T., Gooley, C., Lv, J., Pisani, L., Rusnak, M., Elyanow, R., Takamatsu, H., Zhou, W., Greissl, J., Robins, H., & Carlson, J. M. (2024). Large-scale statistical mapping of T-cell receptor*β*sequences to Human Leukocyte Antigens. In bioRxiv (p. 2024.04.01.587617). 10.1101/2024.04.01.587617

Zander, R., Kasmani, M. Y., Chen, Y., Topchyan, P., Shen, J., Zheng, S., Burns, R., Ingram, J., Cui, C., Joshi, N., Craft, J., Zajac, A., & Cui, W. (2022). Tfh-cell-derived interleukin 21 sustains effector CD8+ T cell responses during chronic viral infection. Immunity, 55(3), 475–493.e5.

Zaslavsky, M. E., Craig, E., Michuda, J. K., Sehgal, N., Ram-Mohan, N., Lee, J.-Y., Nguyen, K. D., Hoh, R. A., Pham, T. D., Röltgen, K., Lam, B., Parsons, E. S., Macwana, S. R., DeJager, W., Drapeau, E. M., Roskin, K. M., Cunningham-Rundles, C., Anthony Moody, M., Haynes, B. F., … Boyd, S. D. (2024). Disease diagnostics using machine learning of immune receptors. In bioRxiv (p. 2022.04.26.489314). 10.1101/2022.04.26.489314

Zdinak, P. M., Trivedi, N., Grebinoski, S., Torrey, J., Martinez, E. Z., Martinez, S., Hicks, L., Ranjan, R., Makani, V. K. K., Roland, M. M., Kublo, L., Arshad, S., Anderson, M. S., Vignali, D. A. A., & Joglekar, A. V. (2024). De novo identification of CD4+ T cell epitopes. Nature Methods, 21(5), 846–856.

Zhou, Z., Reyes-Vargas, E., Escobar, H., Rudd, B., Rockwood, A. L., Delgado, J. C., He, X., & Jensen, P. E. (2016). Type 1 diabetes associated HLA-DQ2 and DQ8 molecules are relatively resistant to HLA-DM mediated release of invariant chain-derived CLIP peptides. European Journal of Immunology, 46(4), 834–845.

Ziegler, A.-G., Bonifacio, E., & BABYDIAB-BABYDIET Study Group. (2012). Age-related islet autoantibody incidence in offspring of patients with type 1 diabetes. Diabetologia, 55(7), 1937– 1943.

