## Supplementary Material for "Identification of a type 1 diabetes-associated T cell receptor repertoire signature from the human peripheral blood"

### Supplementary Note

#### Assessment of sequencing depth

A separate set of replicate experiments were conducted on cohort 1 to gauge the sequencing depth and precision of sequencing technology. The replicate experiment consisted of 7 shallow sequenced replicates, 38 deep sequenced replicates, and one shallow and deep sequenced replicate. The clones are primarily defined as CDR3β sequences, unless explicitly stated otherwise. The clonal frequencies were merged for identical CDR3β AA sequences. The dataset was divided into 2 sections, where repertoires used in shallow vs shallow sequencing comparison were considered **Dataset A** and repertoires used in shallow vs deep sequencing comparison were considered **Dataset B**.

First, to assess the reproducibility of our experiments, we employed Pearson correlation (r) and Morisita-Horn (MH) similarity index, which incorporate both clonal overlap and clonal frequencies in their assessments, on shallow and deep sequenced technical replicates in cohort 1 (see **Methods**). Our results demonstrated high Pearson correlation and MH index values of shallow replicates with (i) shallow replicates ($r = 0.99 \pm 0.01$ and MH index = $0.92 \pm 0.12$ for 7 replicates, **Dataset A**) and (ii) deep replicates ($r = 0.98 \pm 0.03$ and MH index = $0.87 \pm 0.16$ for 38 replicates, **Dataset B**) (Fig. S2). Additionally, we sequenced two shallow and two deep replicates for one individual and observed high clonal overlap among the various combinations of shallow-shallow, shallow-deep, and deep-deep sequenced replicates ($r > 0.98$ and MH index $> 0.82$ for all six possible combinations). Taken together, we demonstrate high technical reproducibility and adequate sequencing depth to capture the clonal diversity of TCRs.

#### Robustness assessment of HLA-based CDR3 restriction based on different clinical groups

We first conducted MMLR-MANOVA tests on the T1D, FDR, and CTRL repertoire sets individually to quantify the association between each HLA site and CDR3β position. These analyses revealed predominantly class II HLA (DQA1, DQB1, and DRB1 genes)-based restrictions on CDR3β amino acid positions in each clinical group, consistent with the observations from cohort 1 (Fig. S5, Fig. S6). Therefore, we further conducted a robustness analysis of CDR3 phenotype and CDR3 risk score based on two main objectives, which includes whether CDR3 phenotypes obtained from specific clinical group (i) show overlap with other clinical statuses and (ii) demonstrate the similar association pattern (T1D>FDR>SDR>CTRL), as observed for cohort 1.

We observed 183, 251 and 68 significant CDR3 phenotypes for T1D, FDR and CTRL clinical groups, respectively using LR test between positional CDR3β frequencies of each AA and HLA risk score. CDR3 phenotypes calculated for each clinical group had significant overlap with each other, for example, the overlap of CDR3 phenotypes was around 63% (43 AAs) for T1D (183 AAs) with respect to CTRL (68 AAs) and 52% (95 AAs) for FDR (251 AAs) with respect to T1D (183 AAs). The number of significant CDR3 phenotypes was proportional to the number of repertoires in each set. Therefore, it is expected to have fewer CDR3 phenotypes for the smaller repertoire set (e.g., only 68 phenotypes were observed for the CTRL group with 182 repertoires).

We further calculated CDR3 risk score for each repertoire using CDR3 phenotypes obtained from each clinical group. We observed that CDR3 risk score shows the same association with each clinical groups as observed in cohort 1, where average CDR3 risk score was highest for the T1D repertoires followed by FDR, SDR and CTRL repertoires (T1D>FDR>SDR>CTRL), respectively, across different lengths, irrespective of which clinical group was used to calculate the CDR3 phenotypes (Fig. S8). Moreover, similar to observation from cohort 1, The CDR3 risk score showed positive correlation with HLA risk score for repertoires from T1D (~0.4%), FDR (~0.46%) and CTRL (~0.54%) clinical groups (Fig. S5, Fig. S6).

#### Assessment of factors influencing CDR3 risk score in T1D

We hypothesized that observed difference in CDR3 risk score among different clinical groups can arise from two possibilities: (i) the count of CDR3β sequences containing CDR3 phenotype was different among repertoires of different clinical groups or (ii) the effect sizes of the CDR3 phenotype affect the overall CDR3 risk score. To address the first possibility, we examined the CDR3β sequences in each repertoire containing at least one CDR3 phenotype (calculated on cohort 1) and observed that there were no significant differences in the percentage of CDR3β sequences across clinical groups (ranging from average of ~90.4% for length 12 to ~99.7% for length 18; Table S7).

The availability of large cohorts resulted in more significant CDR3 phenotypes ($p$-value$<0.05$), which in turn led to a higher percentage of CDR3β sequences containing these CDR3 phenotypes in cohort 1. For the second possibility, we shuffled the effect size of the T1D-associated CDR3β AAs for T1D clinical group and recalculated the CDR3 risk score (Fig. S8D). The expected trend of CDR3 risk score (T1D>FDR>SDR>CTRL) was lost after shuffling the effect sizes, highlighting its importance for the CDR3 phenotypes. This demonstrates that although almost all CDR3β sequences contained at least one CDR3 phenotypes, the risk observed for T1D in TCR cohorts was mainly observed from the effect sizes of these T1D-associated CDR3β AA, in other words how strongly these CDR3β AAs associated with HLA risk score.

#### Role of positive and negative effect sizes of CDR3 phenotypes

The T1D-associated CDR3 phenotypes exhibited both positive and negative effect sizes, indicating that these T1D-associated position specific CDR3β AAs correlate both positively and negatively with the HLA risk score. Therefore, we further assessed the importance of the positively and negatively correlated CDR3 phenotypes in cohort 1. As expected, CDR3 risk score based on positively (positive effect sizes)- and negatively-associated CDR3 phenotype (negative effect sizes) exhibited a positive correlation with HLA risk score (Fig. S5). The correlation of HLA and CDR3 risk scores obtained from either positive or negative effect sizes of CDR3 phenotypes was marginally different from the correlation obtained from the cumulative effect sizes. CDR3 risk scores based on both positive and negative effect sizes were also grouped based on high-risk HLA allele types (DR3 and DR4) for each clinical group, which showed an association with high-risk HLA allele types. The CDR3 risk score for DRX/X (non-risk allele type) HLA individuals was almost consistently lower than for the individuals containing at least one T1D-associated risk allele DR3 or DR4 (Fig. S7). It is important to note that we observed a positive correlation with HLA risk score and higher CDR3 risk score for T1D compared to CTRL repertoires (Fig. S5) for CDR3 risk score obtained from the negatively-associated CDR3 phenotypes (although in negative numbering scale). The negatively-associated CDR3 phenotypes can be utilized to identify CDR3β motifs that were less frequent in T1D repertoires, as observed in subsequent analyses.

#### Analysis of CDR3β motifs obtained from the HLA-based restriction analysis

In the consensus positively-associated HLA-motif (pHLA-motif; [LVSICY][FLWHY][FMWNSIEHY][FWPDVIEHY][FWDEHY]), aromatic and hydrophobic residues were present in almost all positions; whereas, negatively charged residues were predominantly present at the C-terminus of the motif. The C-terminal residues were also more redundant across different lengths, even when additional positions were included for longer CDR3β lengths (Extended Data Fig. 4). Similarly, a consensus negatively-associated HLA-motif (nHLA-motif; [KR][AGHIKMTV][ADKRT][GKQRT][GIKLTV]) was also derived from the negatively-associated CDR3 phenotype (Extended Data Fig. 5). Negatively-associated CDR3 phenotypes had relatively fewer observable recurring patterns across different CDR3β lengths (see **Methods**). However, positively-charged residues were predominant in almost all positions in the motif, showing conserved biophysical compatibility among HLA alleles and restricted TCR repertoire (*165*).

It is important to note that pHLA- and nHLA-motif scores have varying frequency within a repertoire and should not be compared with each other. In summary, we successfully identified HLA-associated CDR3β motifs exhibiting gradual overrepresentation or diminution across different clinical groups in cohort 1.

#### Performance of the conventionally considered high-risk HLA alleles in classification of immune repertoires

The HLA DR3 and DR4 alleles were strongly associated with a high-risk of T1D. Consequently, we utilized an individual's HLA type to evaluate the predictability of their clinical statuses with regards to T1D. By considering the presence of at least one DR3 or DR4 allele as a predictor of T1D, we achieved an accuracy of 57.1%, (sensitivity of 75.6% and specificity of 48.2%) in cohort 1 and 80.1% (sensitivity of 82.3% and specificity of 71.9%) in cohort 2 and 3 (Extended Data Fig. 7A). We also employed stricter criteria, requiring both alleles to be either DR3 or DR4 as a predictor of T1D. This led to an accuracy improvement to 72.4%. However, we observed a significant decrease in sensitivity (32.1%) while specificity increased substantially to 91.8%. Whereas cohort 2 and 3 showed accuracy of 47.5% (sensitivity of 33.8% and specificity of 97.2%). Approximately 85% of the repertoires in cohort 1 carried at least one HLA DRX allele, and the majority of these repertoires were present in non-T1D clinical groups. This explains the higher specificity observed with the stricter criteria. Indeed, our results aligned closely with the documented knowledge that the presence of two T1D risk-associated HLA alleles (i.e., DR3/3, DR4/4, DR3/4) associates with a higher incidence of T1D compared to one or no copies of these risk alleles (e.g., DR3/X, DR4/X, DRX/X) (*166*, *167*). However, such heterozygous individuals carrying HLA DR3/DR4 were relatively few in number (n=211 with 61.1% of those belonging to the T1D group), and among the 1149 individuals with one or no allele containing DR3/DR4, 276 (24%) had T1D leading to lower sensitivity.

#### Age-based confounding factors reduction using sample weights

We derived age-dependent sample weights to balance data distribution across age groups and disease classes. We first computed age bins and assigned each sample to an appropriate age group, then calculated sample weights for each sample based on the distribution of age groups and disease states. These sample weights were designed to correct for over-representation by down-weighting samples from highly represented groups and up-weighting those from underrepresented groups, effectively normalizing the contribution of each sample to the overall loss function. The pseudo-code for the process is given below:

```
total_n_samples = 1298 # total number of samples in cohort 1
age_groups = bin_ages(ages, n_bins=10)
contingency_table = create_contingency_table(disease_states, age_group)
for cell in contingency_table:
      cell    =    n_samples_in_age_group    *    n_samples_in_disease_state    /
(total_n_samples * cell)

# For (age_group, disease) combinations with no samples
replace_infs_with_1(contingency_table)

# sum of sample weights is equal to the number of samples
weights = get_weights_based_on_disease_and_age_group(data, contingency_table)
```

### Supplementary Tables

**Table S1 | Statistical Overview of the McPAS and VDJdb Databases (as of April, 2024).**

**Table S2 | Distribution of pathology (McPAS) and antigen species (VDJdb)-associated CDR3β sequences across clinical groups present in cohort 1.** The normalized frequency of the CDR3β clones was calculated by dividing the clonal frequency of the overlapping CDR3β clones in a repertoire by the total sum of clonal frequencies of all CDR3β clones in the same repertoire. Values are presented as mean ± standard deviation and p-values were obtained from Kruskal–Wallis test and adjusted for multiple testing using the Benjamini-Hochberg procedure. Significant values where normalized frequencies are higher for T1D compared to other clinical groups are highlighted in violet.

**Table S3 | Unique V-gene family-CDR3β centroids assigned to either MHC class I or class II alleles.** Values with FDR < 0.2 that are overrepresented (red) or underrepresented (blue) in the T1D clinical group are highlighted.

**Table S4 | P-values obtained from the MMLR-MANOVA tests for the whole cohort 1 and each clinical group (T1D, FDR and SDR).** P-values were adjusted for multiple testing using the Benjamini-Hochberg procedure.

**Table S5 | The odds ratios for T1D risk based on DRB1 and DQB1 HLA alleles** (sourced from https://github.com/immunogenomics/cdr3-QTL/tree/main/data/genotype).

**Table S6 | Observed CDR3 phenotypes and their effect sizes for Cohort 1 and for each clinical group (T1D, FDR, CTRL).** The significant CDR3 phenotypes (p-value<0.05) are highlighted in blue.

**Table S7 | The average percentage of CDR3β sequences containing at least one CDR3 phenotype in each clinical group and for different CDR3β sequence lengths.**

**Table S8 | Calculation of positively- (pHLA) and negatively- (nHLA) associated HLA-motifs by combining the IMGT-numbered positions from CDR3β sequence lengths L13 to L16.**

**Table S9 | Performance of the different methods utilized in the current study on cohort 1, and held-out test cohorts 2 and 3.** Note: some methods, such as HLA risk score and different motif-based approaches, may not involve direct training on cohort 1.

**Table S10 | Performance matrices for each split in statistical classification method using public clones.** The best-performing encoding is highlighted in violet.

**Table S11 | T1D-associated V-gene/CDR3/J-gene clones obtained from statistical classification method using public clones.**

#### Metadata information

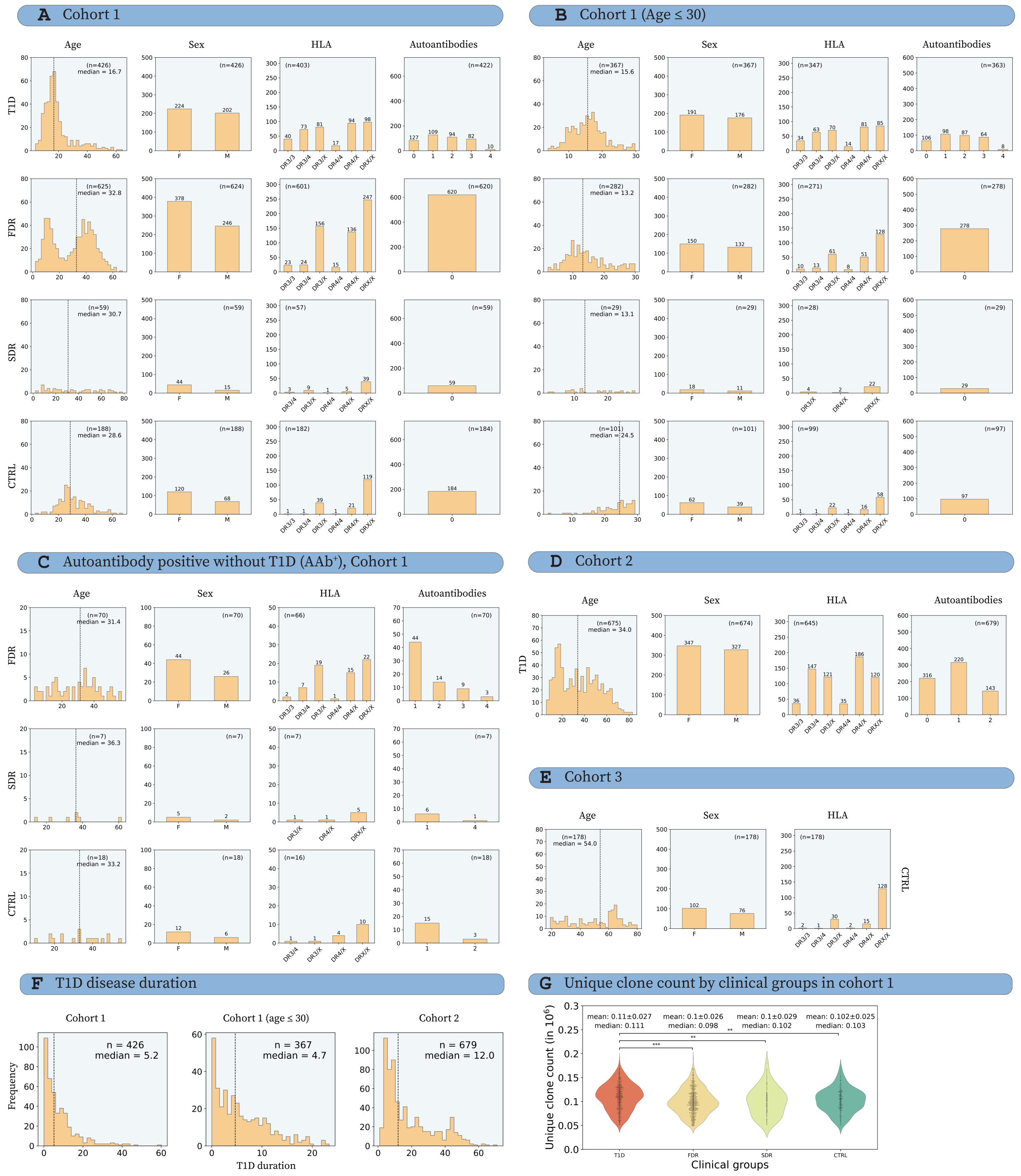


**Fig. S1** | **The metadata information for the TCRβ repertoires.** The metadata information includes age, sex, human leukocyte antigen (HLA) and autoantibody distribution in the **(A)** cohort 1, **(B)** cohort 1 filtered based on age (age ≤ 30 years), **(C)** "AAb+" repertoires in cohort 1, which are at risk of developing T1D (i.e., contains significant concentration of one or more autoantibodies but not classified under T1D clinical group), **(D)** cohort 2 (contains only T1D repertoires) and **(E)** cohort 3 (contains only control repertoires). The autoantibody information was not available for the TCRβ repertoires in cohort 3. "AAb+" repertoires, although part of cohort 1, were kept as a separate category and excluded from all analyses unless specified. **(F)** The duration (in years) for T1D clinical group in: cohort 1, cohort 1 with age ≤ 30 years and cohort 2, respectively. "n" is the number of datapoints considered in the respective metadata information plot. The median line was plotted for all age-related metadata information along with the median value. **(G)** The number of unique clones observed in cohort 1, when grouped by clinical groups, shows minimal differences in both mean and median values. p-values for pairwise testing were calculated using two tailed Mann-Whitney U tests and p-values were adjusted between clinical groups. All p-values were adjusted for multiple testing using the Benjamini–Hochberg method. p-values were described as * for [0.01,0.05], ** for [0.001,0.01] and *** for <0.001 and no stars plotted for non-significant values. Relates to Fig. 1.

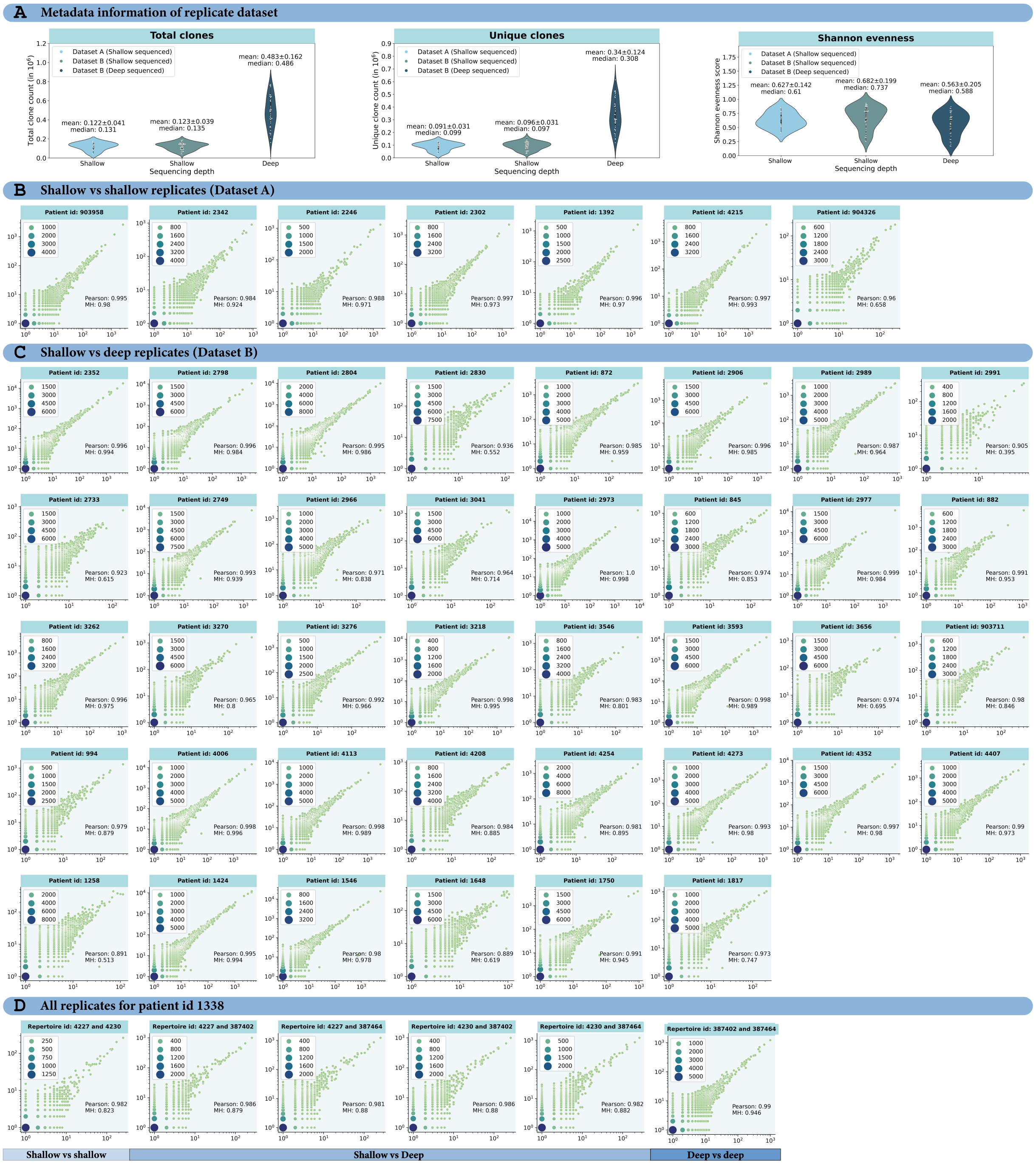


**Fig. S2 | High overlap observed between the technical replicates of shallow vs shallow and shallow vs deep sequenced repertoires demonstrate high reproducibility of the experiments.** Technical replicates are defined as two samples obtained from the blood of the same patient, while clones are defined as unique CDR3β sequences. **(A)** metadata information (total clones, unique clones and shannon evenness) of the technical replicate dataset, where repertoires used in shallow vs shallow sequencing comparison were considered **Dataset A** and repertoires used in shallow vs deep sequencing comparison were considered **Dataset B.** To assess the reproducibility of our experiments, we employed Pearson correlation (r) and Morisita-Horn (MH) similarity index, which incorporate both clonal overlap and clonal frequencies in their assessments (annotated on each plot). Scatterplots of sequence counts of one replicate versus the other replicate of the same patient were presented for **(B)** shallow vs shallow sequenced repertoires and **(C)** shallow vs deep sequenced repertoires. **(D)** patient id 1338 that had two shallow and two deep sequenced repertoires. Therefore, all combinations of shallow and deep sequenced repertoires were plotted. The results demonstrated high Pearson correlation and MH index values of shallow replicates with (i) shallow replicates (r = 0.99 ± 0.01 and MH index = 0.92 ± 0.12 for 7 replicates) and (ii) deep replicates (r = 0.98 ± 0.03 and MH index = 0.87 ± 0.16 for 38 replicates) . Additionally, two shallow and two deep sequenced replicates for patient id 1338 observed high clonal overlap among the various combinations of shallow-shallow, shallow-deep, and deep-deep sequenced replicates ($r > 0.98$ and MH index > 82 for all six possible combinations). Taken together, we demonstrate high technical reproducibility and adequate sequencing depth to capture the clonal diversity of TCRs. Relates to Fig. 1.

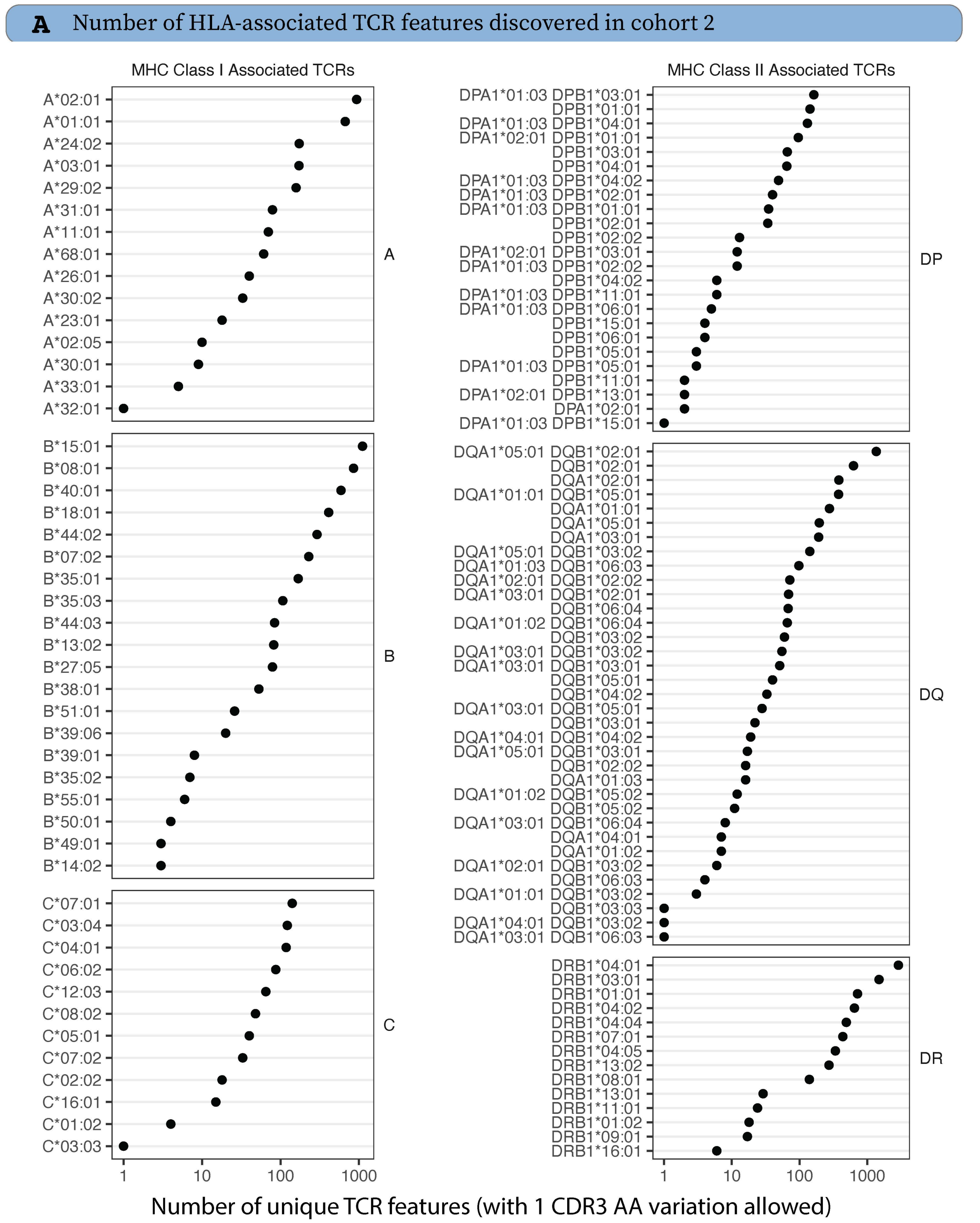


**Fig. S3** | **Number of HLA-associated TCR features discovered in cohort 2**. A TCR was hypothesized as HLA-associated if its V-family constrained exact CDR3 match or edit-distance 1 CDR3 match was overrepresented in repertoires of persons with a particular HLA allele or haplotype (p-value < 1e-8 and prevalence within HLA-matched group > 5%, and prevalence in HLA-mismatched repertoires < 10%. Relates to Fig. 3.

**A** Odds ratio and unadjusted p-value of CDR3β sequences for each HLA allele

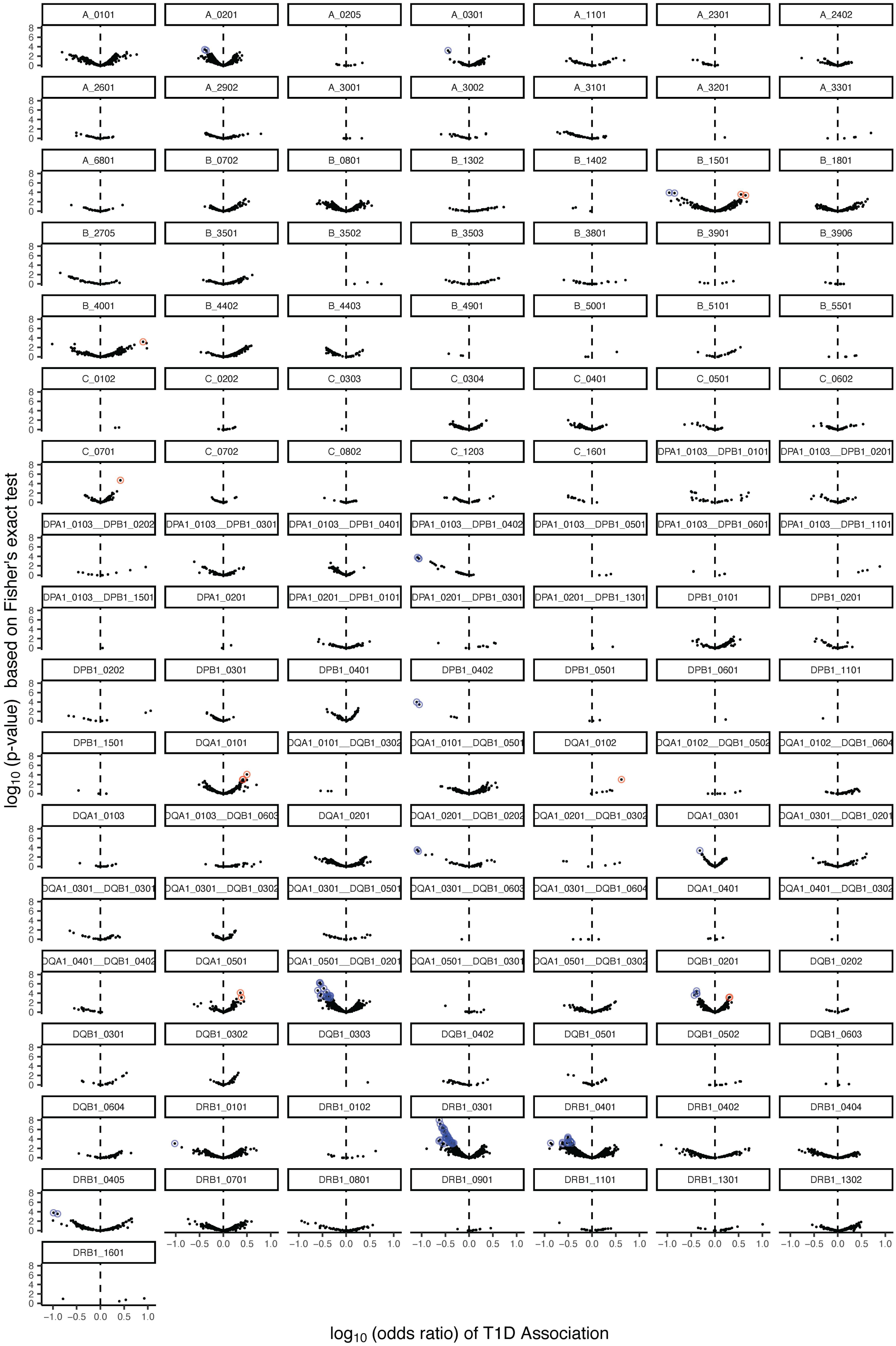

**Fig. S4** | **HLA-conditioned enrichment or depletion of TCRs based on T1D clinical status.** TCRs were identified as HLA-associated based on odds ratio of detection in cohort 2. Subsequently we tabulated detection of these TCRs and their v-family constrained one mutation distance neighbors among participants expressing the predicted restricting allele in cohort 1 and tested for enrichment or depletion based on T1D clinical status (as detailed in Table S3). Volcano plots show odds ratio of T1D association and unadjusted negative log10 p-values. TCRs with statistically significant T1D-association (Fisher's exact test, false discovery rate < .2) are shown as red (T1D-enriched) or blue (T1D-depleted) circles. Relates to Fig. 3.

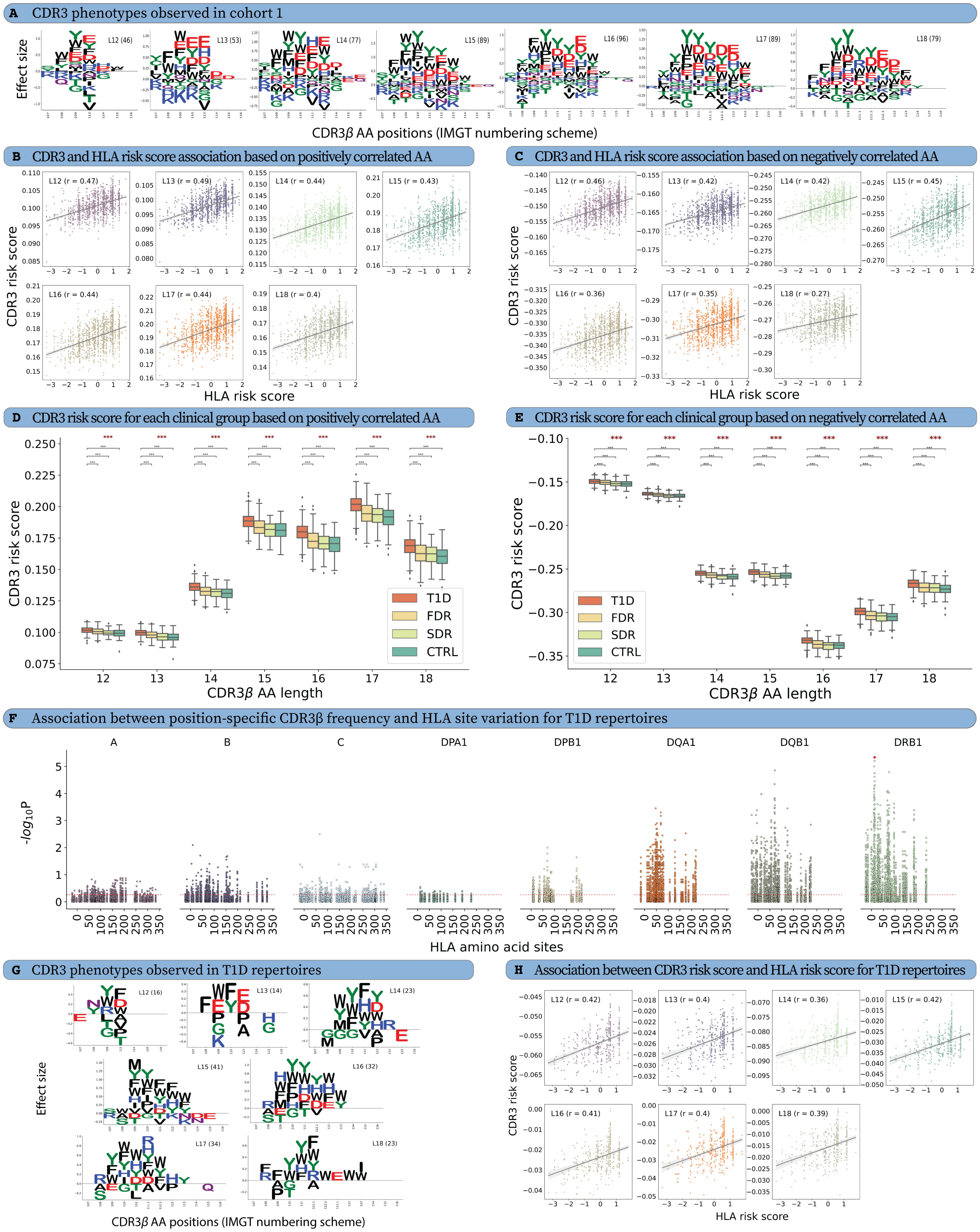


**Fig. S5** | **Assessment of positively- and negatively-associated CDR3 phenotypes obtained from cohort 1 and HLA-based restriction of CDR3β sequences in T1D clinical group of cohort 1. (A)** The sequence logo plot of CDR phenotypes and their effect sizes (coefficients), obtained from the LR analysis of each CDR3β length (L12-L18), using cohort 1. In cohort 1, there were 304 positive and 225 negative significant ($p-value \leq 0.05$) associations present between CDR3 phenotypes and HLA risk score, for all CDR3β lengths. We calculated the correlation with HLA risk score when CDR3 risk score was calculated using **(B)**

positively-associated CDR3 phenotypes and **(C)** negatively-associated CDR3 phenotypes. CDR3 risk score calculated using **(D)** positively-associated and **(E)** negatively-associated CDR3 phenotypes also showed association with clinical groups and observed the expected trend, where T1D>FDR>SDR>CTRL. **(F)** The MANOVA test p-values from the MMLR analysis of the 402 T1D repertoires, where all CDR3β positions were plotted for each HLA site containing mutation(s). The p-values (≤0.05) above the red line were significant. The lowest p-value observed for cohort 1 (HLA-DRB1 site 13 with CDR3β position 111 for L15) is mapped onto the MANOVA test p-values and highlighted in red color. **(G)** The logo plot represents CDR3 phenotypes and their effect size (coefficient) obtained from the LR analysis using T1D clinical group in cohort 1. **(H)** The correlation between HLA risk score and CDR3 risk scores calculated for T1D repertoires, for each CDR3β length (L12-L18). The correlation (r) values were shown for each length in appropriate plots. Multiple testing was performed using Kruskal–Wallis test (denoted with red stars) and p-values were adjusted between different CDR3β lengths. p-values for pairwise testing were calculated using two tailed Mann-Whitney U tests and p-values were adjusted between clinical groups. All p-values were adjusted for multiple testing using the Benjamini–Hochberg method. p-values were described as * for [0.01,0.05], ** for [0.001,0.01] and *** for <0.001 and no stars plotted for non-significant values. Relates to Fig. 3.

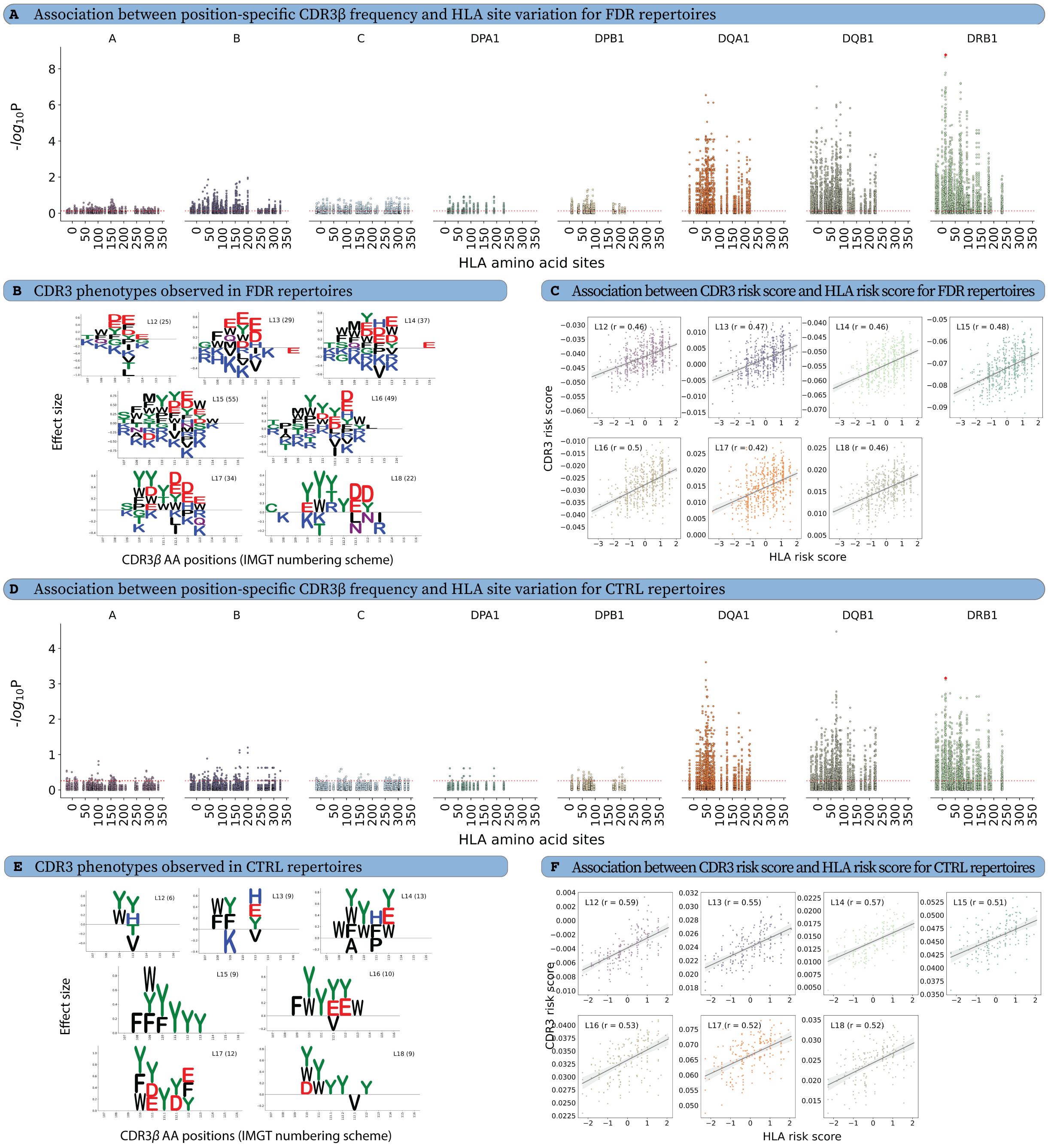


**Fig. S6 | Assessment of HLA-based restriction of CDR3β sequences in FDR (first degree relatives) and CTRL (control) groups of cohort 1.** The MANOVA test p-values from the MMLR analysis of the **(A)** 601 FDR and **(D)** 182 CTRL repertoires, where all CDR3β positions were plotted for each HLA site containing mutation(s). The p−values (≤0.05) above the red line were significant. The logo plots represent CDR3 phenotypes and their effect sizes (coefficient) obtained from the LR analysis using **(B)** FDR and **(E)** CTRL repertoires. The correlation between HLA risk score and CDR3 risk scores calculated for **(C)** FDR and **(F)** CTRL repertoires, for each CDR3β length (L12-L18). The correlation (r) values were shown for each length in appropriate plots. The lowest p-value observed for cohort 1 (HLA-DRB1 site 13 with CDR3β position 111 for L15) is mapped onto the MANOVA test p-values and highlighted in red color. Relates to Fig. 3.

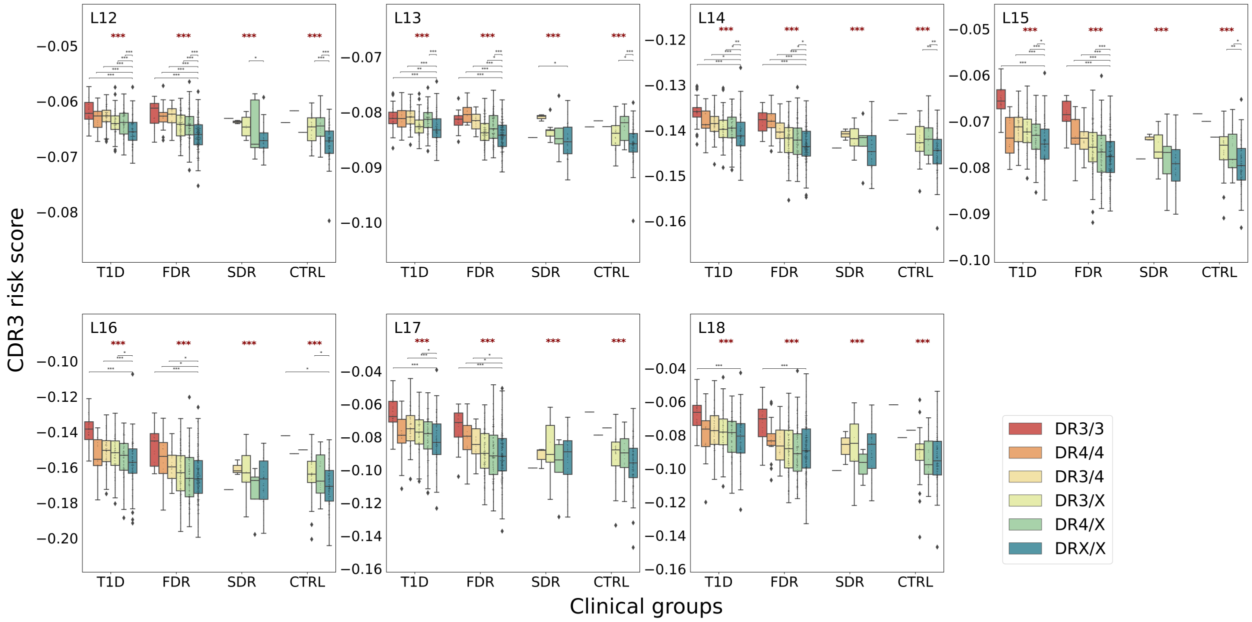


**Fig. S7 | CDR3 risk score shows association with conventional T1D-linked high-risk HLA alleles (DR3 and DR4) at different CDR3β lengths within clinical groups, in cohort 1.** The CDR3 risk score was higher in individuals containing at least one high-risk HLA allele regardless of clinical groups. In the figure, multiple testing was performed using Kruskal–Wallis test (denoted with red stars) and p-values were adjusted between different clinical groups. p-values for pairwise testing were calculated using two tailed Mann-Whitney U tests and p-values were adjusted between different high-risk HLA alleles. All p-values were adjusted for multiple testing using the Benjamini–Hochberg method. p-values were described as * for [0.01,0.05], ** for [0.001,0.01] and *** for <0.001 and no stars plotted for non-significant values. Relates to Fig. 3.

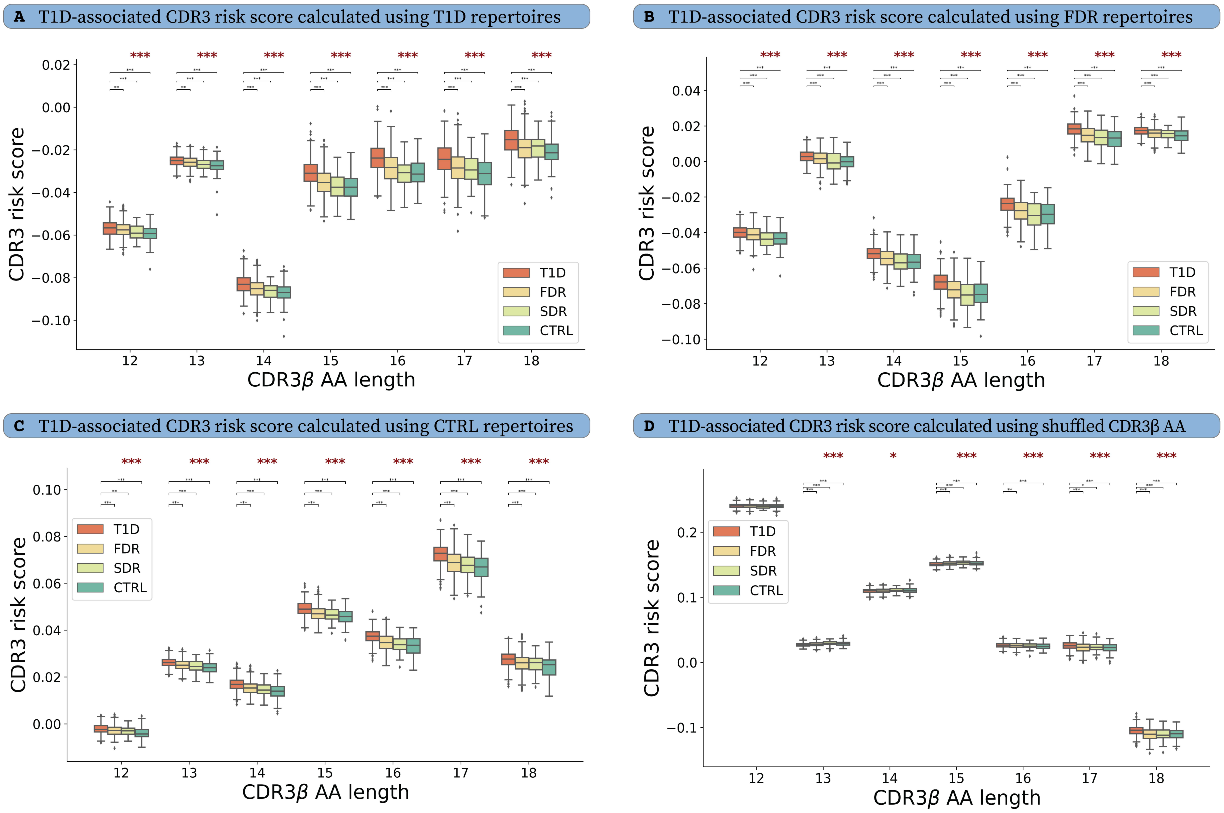


**Fig. S8 | The association between CDR3 risk scores and clinical groups exhibits a high level of consistency and robustness, suggesting a strong and reliable association.** The CDR3 phenotypes obtained from each clinical group were further used to calculate the CDR3 risk score for all clinical groups. Further, CDR3 risk score was clustered based on clinical groups, where CDR3 phenotypes were identified using **(A)** T1D **(B)** FDR and **(C)** CTRL clinical groups. **(D)** As a validation test, we shuffled the effect sizes (LR coefficients) of the CDR3 phenotypes obtained from cohort 1 to calculate the CDR3 risk score. The observed trend, where T1D>FDR>SDR>CTRL was lost upon shuffling the effect sizes of CDR3 phenotypes. In the figure, multiple testing was performed using Kruskal–Wallis test (denoted with red stars) and p-values were adjusted between different CDR3β lengths. p-values for pairwise testing were calculated using two tailed Mann-Whitney U tests and p-values were adjusted between different clinical groups. All p-values were adjusted for multiple testing using the Benjamini–Hochberg method. p-values were described as * for [0.01,0.05], ** for [0.001,0.01] and *** for <0.001 and no stars plotted for non-significant values. Relates to Fig. 3.

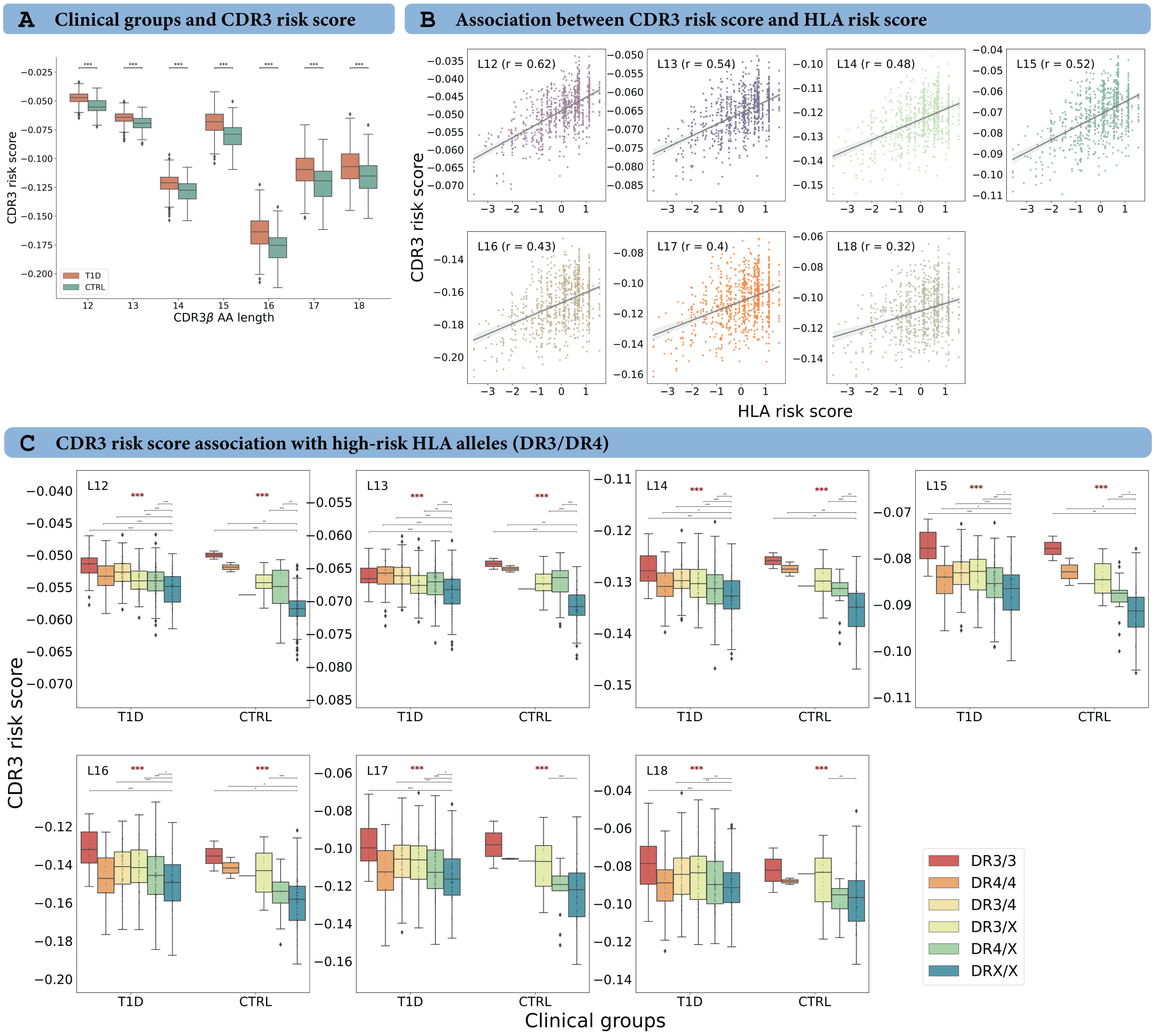


**Fig. S9 | Assessment of CDR3 risk score calculated on cohort 2 and 3 based on the CDR3 phenotypes obtained from cohort 1.** The CDR3 risk score calculated on cohort 2 and 3 **(A)** showed association with T1D and CTRL clinical groups, where T1D>CTRL; and **(B)** showed positive correlation with HLA risk score for different CDR3β length (L12-L18). **(C)** CDR3 risk score also showed association with presence of high-risk HLA alleles (DR3 and DR4) in cohort 2 and 3, when clustered by clinical groups. In figure **panel C**, multiple testing was performed using Kruskal–Wallis test (denoted with red stars) and p-values were adjusted between different clinical groups. p-values for pairwise testing were calculated using two tailed Mann-Whitney U tests and p-values were adjusted between different high-risk HLA alleles (and between clinical groups in **panel A**). All p-values were adjusted for multiple testing using the Benjamini–Hochberg method. p-values were described as * for [0.01,0.05], ** for [0.001,0.01] and *** for <0.001 and no stars plotted for non-significant values. Relates to Fig. 3.

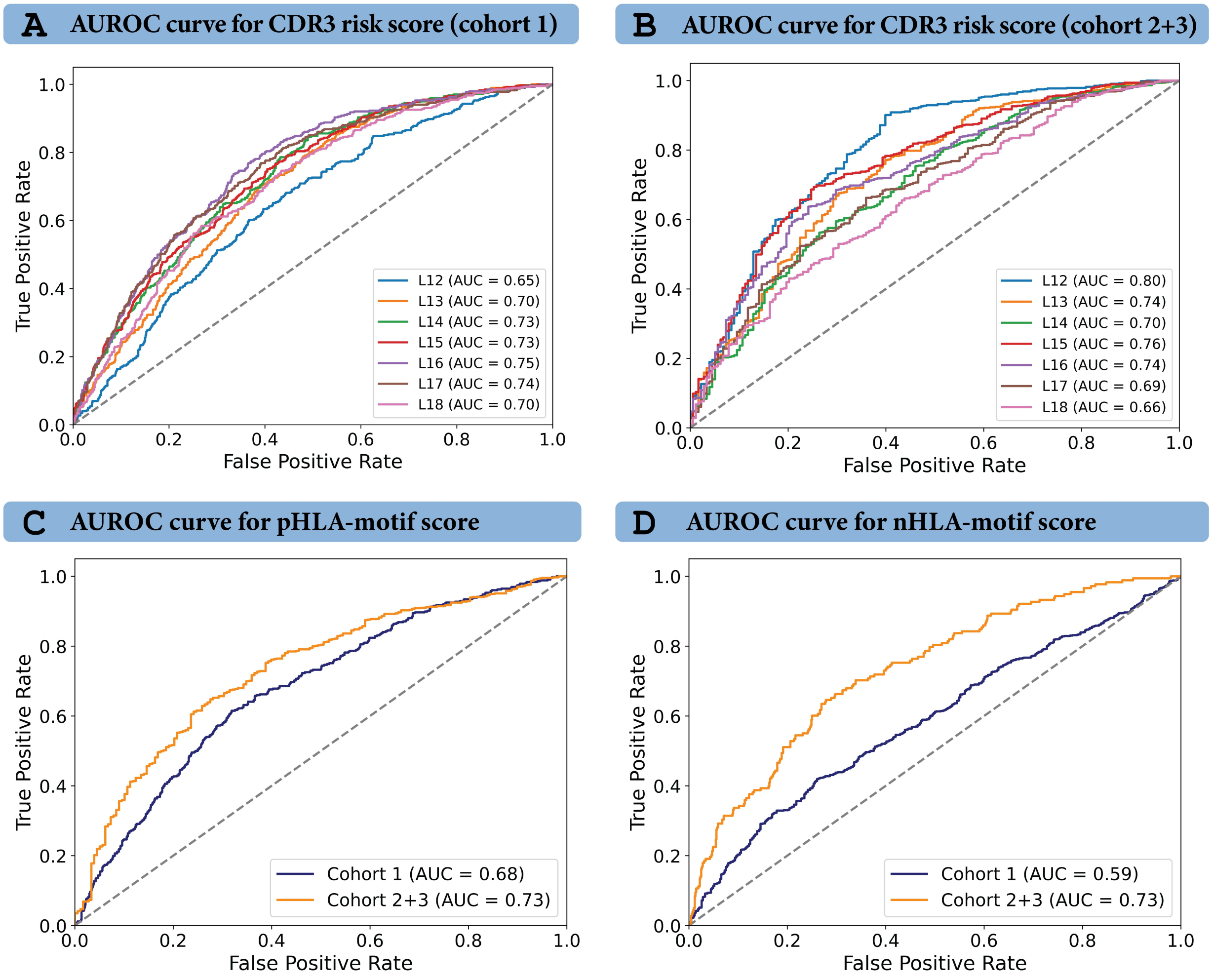


**Fig. S10 | CDR3 risk scores and HLA-associated motif scores derived from the TCR repertoire (TCR features) in the HLA-based restriction study demonstrate good performance in classification of T1D and non-T1D repertoires.** The AUROC curves for classification were plotted for **(A)** CDR3 risk score calculated for each CDR3β length (L12-L18) for cohort 1, **(B)** CDR3 risk score calculated for cohort 2 and 3 using the CDR3 phenotypes obtained from cohort 1, **(C)** pHLA-motif score and (D) nHLA-motif score. Relates to Fig. 4.

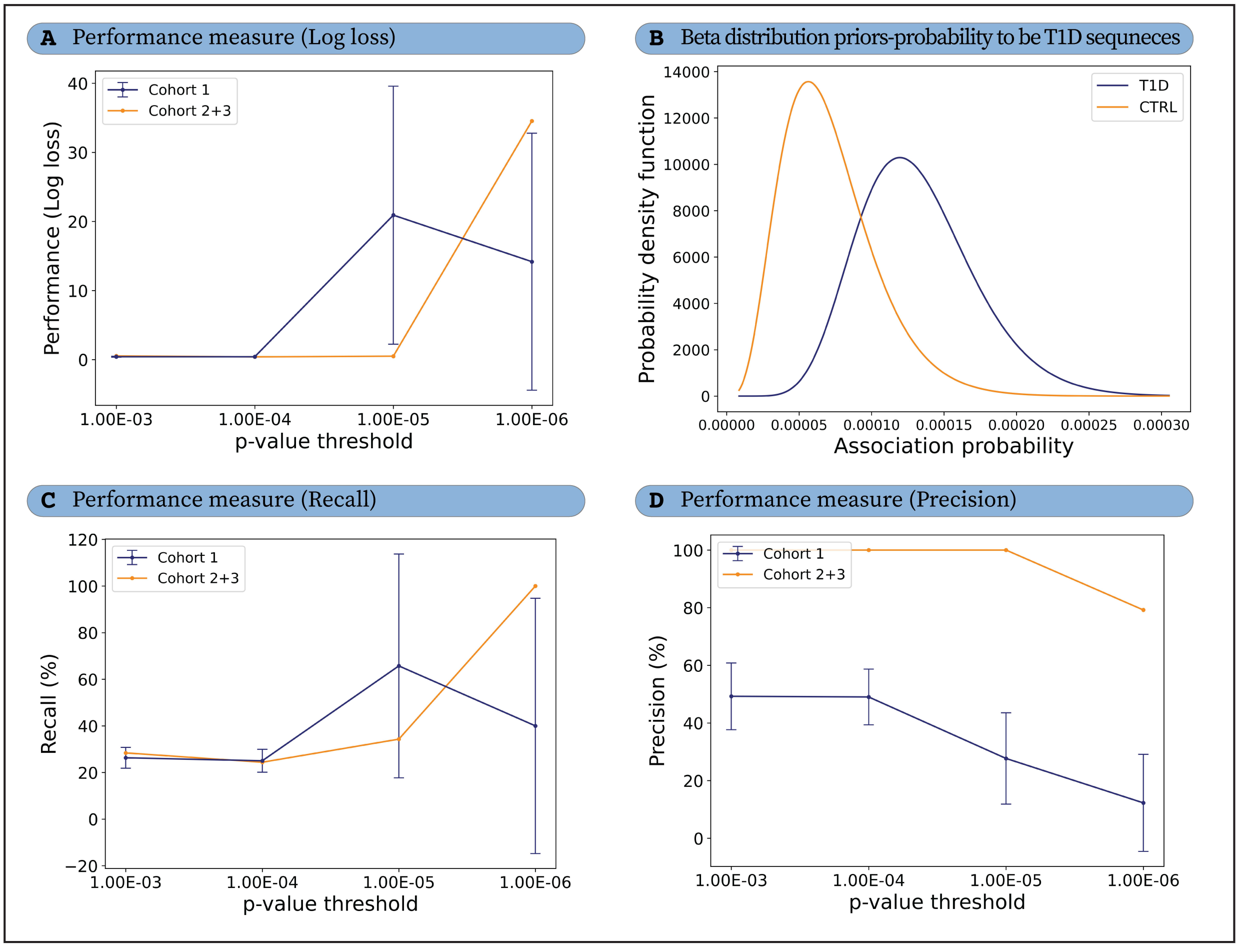


**Fig. S11** | **The performance of the statistical classification framework based on public clones demonstrated high variability in the results on the training cohort 1 and test cohorts 2+3, at different selection thresholds. (A)** The classification of TCR repertoires was optimized using the Log loss function. **(B)** The beta-distribution priors for T1D and CTRL classes demonstrated poor class separation. **(C)** The recall for the model was low for all threshold values. **(D)** The precision for the model was low for cohort 1 but unexpectedly high for cohort 2+3 (potentially due to high sequencing depth in cohort 3). Relates to Fig. 4.

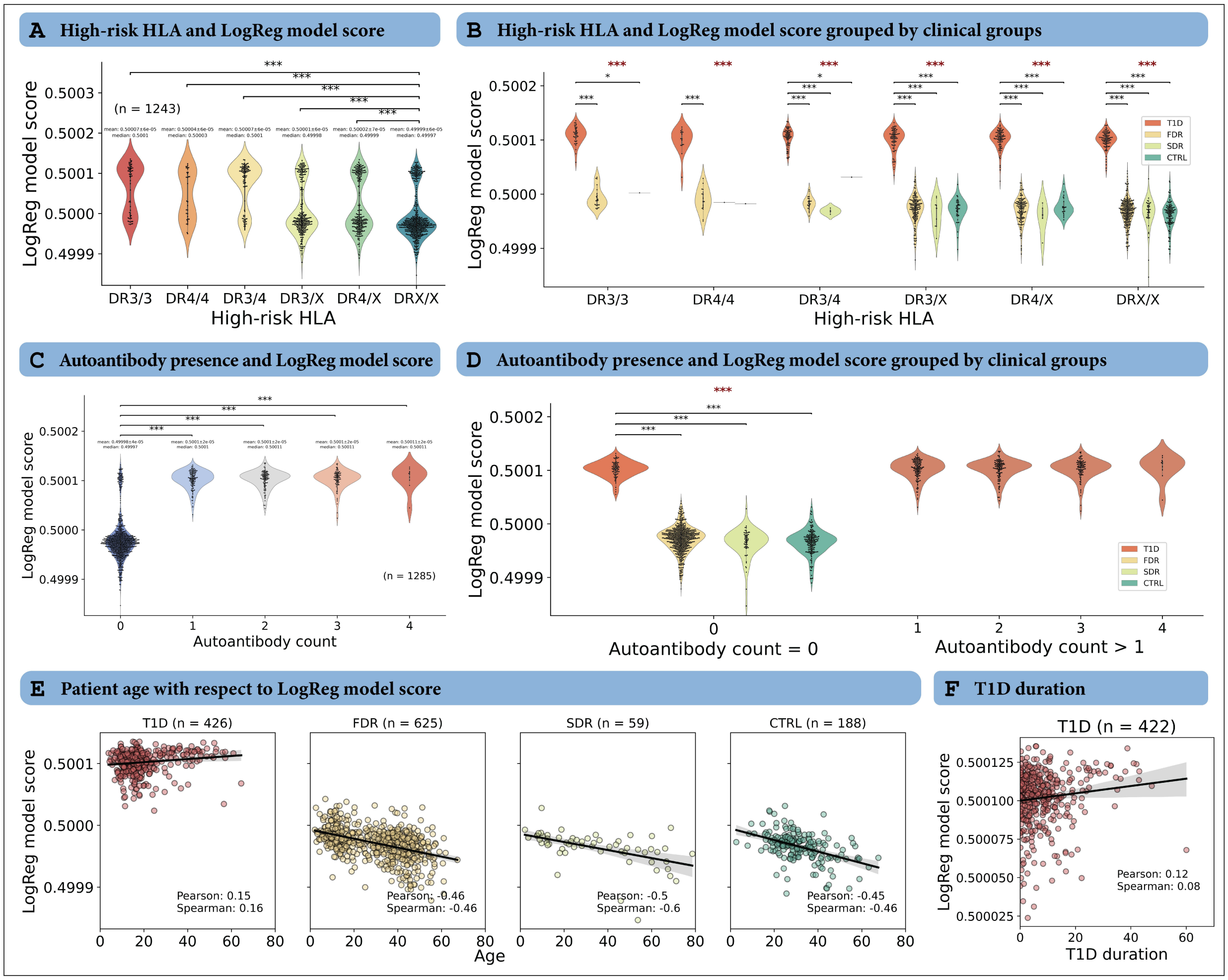


**Fig. S12 | The logistic regression model exhibits signs of overfitting on cohort 1. (A)** The logistic regression (LogReg) model scores in repertoires containing high-risk HLA alleles (DR3 or DR4) exhibited a significant difference compared to other HLA types (DRX/X). Notably, all combinations of high-risk HLA alleles formed two distinct clusters. The p-values in the figure were calculated with respect to the DRX/X HLA type. **(B)** The model score was plotted for each high-risk HLA type and grouped by the clinical groups. The model score was significantly high for T1D repertoires in all high-risk HLA combinations and did not show expected observation of T1D>FDR>SDR>CTRL. Here, pairwise testing was performed with respect to the T1D clinical group. **(C)** The LogReg model score also had significantly low values when no autoantibody were present. The p-values were calculated with respect to no presence of autoantibodies. **(D)** The LogReg model score was high for autoantibody count >0. However, in case of no presence of autoantibody, LogReg model score was significantly higher for the T1D repertoires and did not follow the expected trend of T1D>FDR>SDR>CTRL. **(E)** Interestingly, the LogReg model score plotted against the age of the patient, showed a positive correlation for T1D group and negative correlation for remaining clinical groups. **(F)** The T1D duration also showed a positive correlation with LogReg model score (pearson correlation: 0.27 and spearman correlation: 0.17). In the figure, the p-values for the multiple testing were performed using Kruskal–Wallis test (denoted with red stars). Similarly, p-values for pairwise testing were calculated using two tailed Mann-Whitney U test. p-values were described as * for [0.01,0.05], ** for [0.001,0.01] and *** for <0.001 and no stars plotted for non-significant values. All p-values were adjusted for multiple testing using the Benjamini–Hochberg method. Relates to Fig. 4.

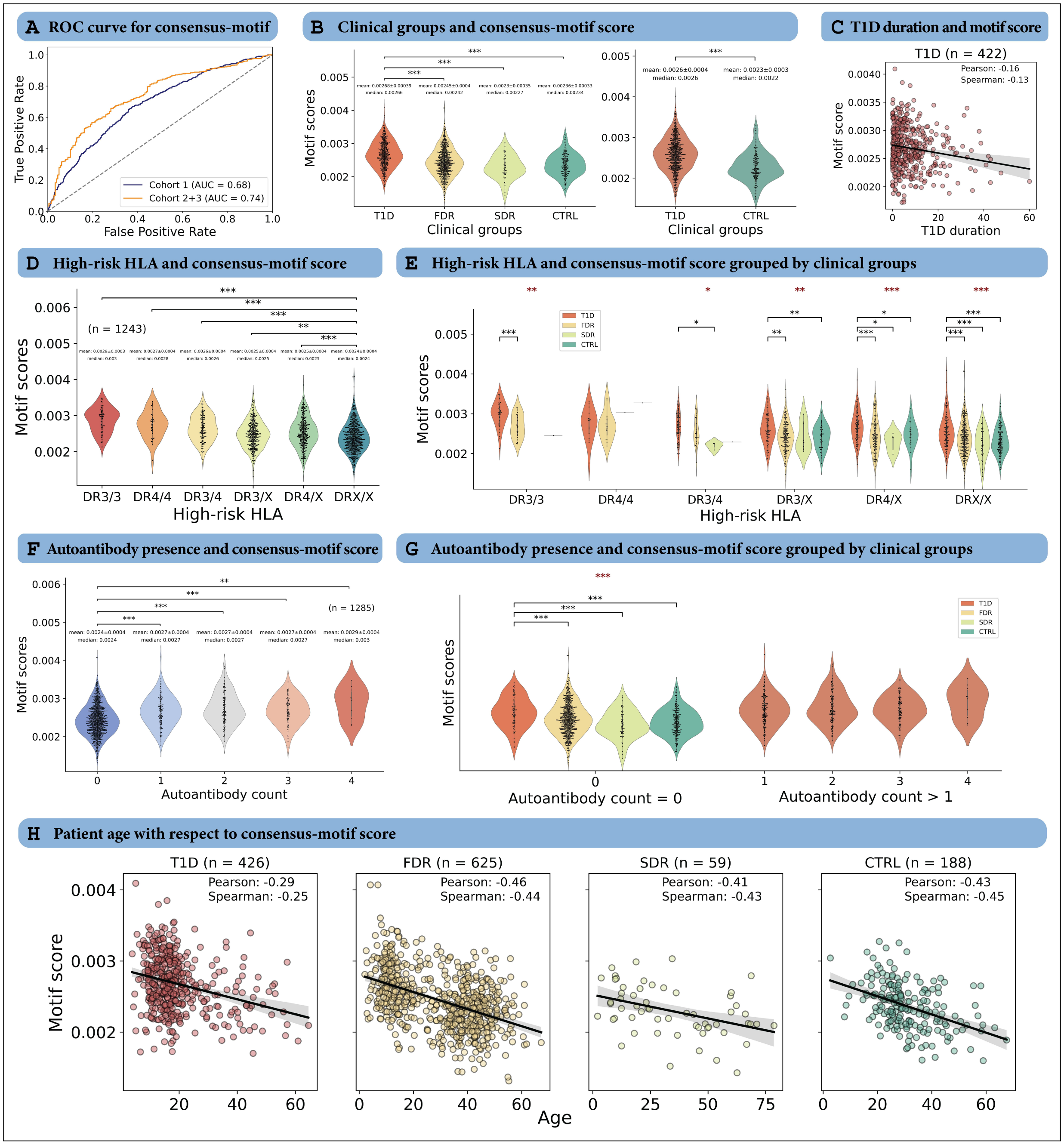


**Fig. S13** | **The assessment of the consensus-motif, derived from the intersection of pHLA-motif and DeepRC-motif, shows positive association with the clinical groups, high-risk HLA alleles (DR3/DR4) and autoantibody presence. (A)** consensus-motif score was used for classification, where AUROC was 0.68 for cohort 1 and 0.74 for cohort 2+3. **(B)** The consensus-motif score was higher for T1D repertoires in cohort 1, following the expected trend of T1D > FDR > CTRL. A similar pattern was also observed in cohorts 2 and 3. The consensus-motif score represents the number of CDR3β sequences containing both pHLA-motif and DeepRC-motif normalized by total number of CDR3β sequences per repertoire, where higher consensus-motif score signifies higher risk of T1D. **(C)** The correlation between T1D duration and consensus-motif score shows a pearson correlation of -0.16 and spearman correlation of -0.13. **(D)** The consensus-motif score was significantly higher in the high-risk HLA type (DR3 or DR4) individuals than other HLA types (DRX/X). The p-values in the figure were calculated with respect to the DRX/X HLA type. **(E)** The consensus-motif score was plotted for each high-risk HLA type and grouped by the clinical groups. The consensus-motif score observed the expected trend T1D>FDR>CTRL in all cases irrespective of high-risk HLA allele type. Here, pairwise testing was performed with respect to the T1D clinical group. **(F)** The consensus-motif score also showed positive association with the number of autoantibodies present. The p-values were calculated with respect to no presence of autoantibodies. **(G)** The consensus-motif score was high for autoantibody count >0. However, in case of no presence of autoantibody, consensus-motif score followed the expected trend where T1D>FDR>CTRL. **(H)** The consensus-motif score was plotted against age of the patient for different clinical groups (for cohort 1). Each clinical group showed negative correlation with the age of the patient. In the figure, the p-values for the multiple testing were performed using Kruskal–Wallis test (denoted with red stars). Similarly, p-values for pairwise testing were calculated using two tailed Mann-Whitney U tests. p-values were described as * for [0.01,0.05], ** for [0.001,0.01] and *** for <0.001 and no stars plotted for non-significant values. All p-values were adjusted for multiple testing using the Benjamini–Hochberg method. Relates to Fig. 3.

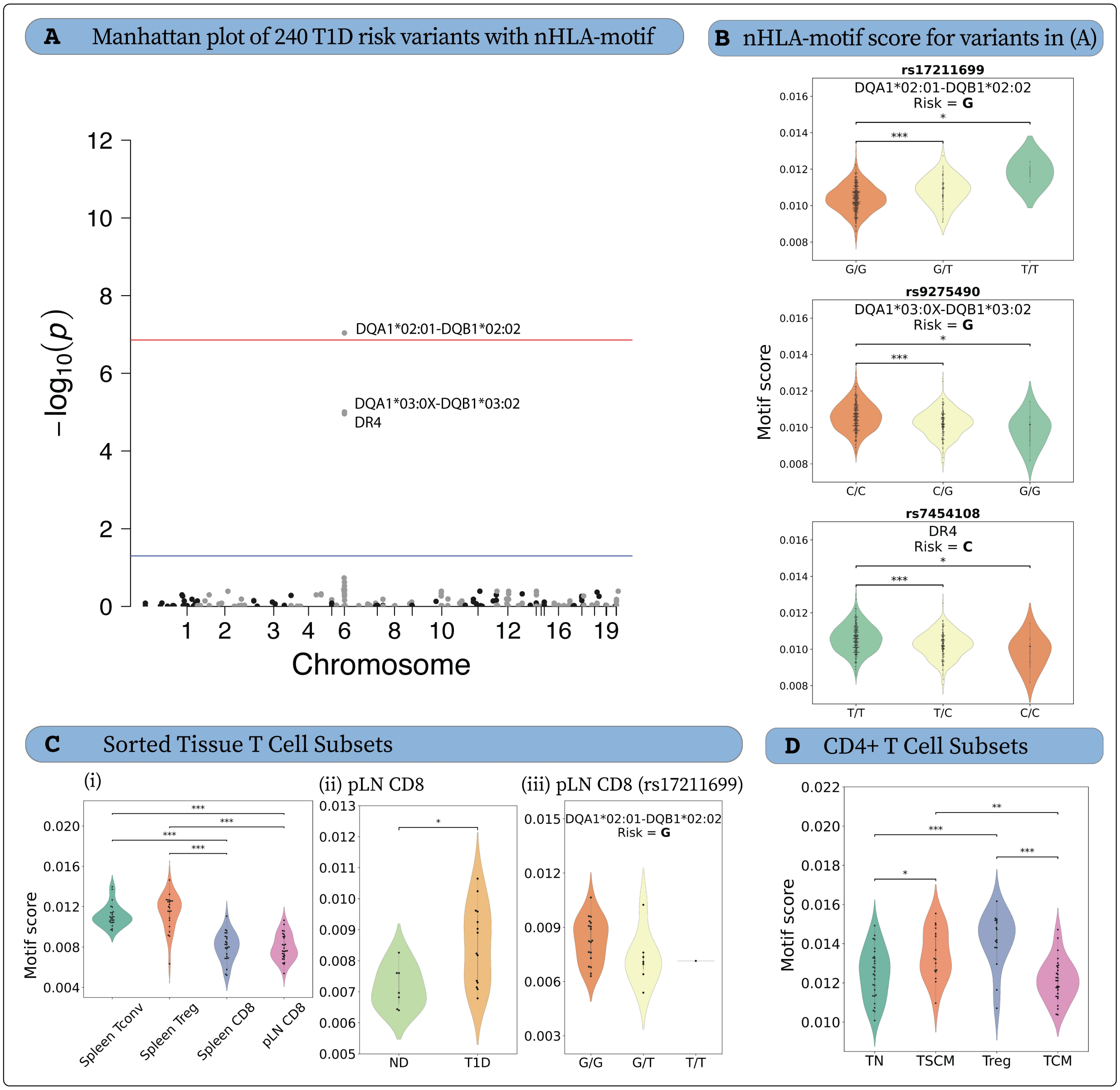


**Fig. S14** | **Negatively-associated HLA-motif (nHLA-motif) enriched in carriers of T1D protective genetics in bulk peripheral blood and carriers of T1D risk genetics in pancreatic lymph node CD8$^+$ T cells.** (**A**) Manhattan plot of 240 T1D risk variants versus nHLA-motif score in bulk peripheral blood. Linear regression assuming additive genotypic effect with age, sex, T1D status, predicted probability of CMV infection, and 10 multidimensional scaling components as covariates. Benjamini-Hochberg false discovery rate threshold at p=1.38e-7 (red line) and a conventional threshold of p = 0.05 (blue line). **(B)** Violin plots of nHLA-motif score in European ancestry non-diabetic (ND) subjects, according to significantly associated variants in **(A)**. Tagged HLA genotype and risk allele annotated above each plot. (**C**) nHLA-motif score across sorted T cell subsets in spleen and pancreatic lymph node (pLN). **(i)** Mixed-effects analysis with Tukey's multiple comparisons test. (**ii**) pLN CD8$^+$ T cell nHLA-motif score according to T1D status. (**iii**) pLN CD8$^+$ T cell nHLA-motif score according to DQA1*02:01-DQB1*02:02 tag SNP. **(D)** nHLA-Motif score across sorted CD4+ T cell subsets, where mixed-effects analysis was done with Tukey's multiple comparisons test. Linear regression with age, sex, diabetes status, and 10 multidimensional scaling components as covariates. In all violin plots, p-values for pairwise testing were calculated using two tailed Mann-Whitney U tests and p-values were adjusted between different clinical groups using the Benjamini–Hochberg method. p-values were described as * for [0.01,0.05], ** for [0.001,0.01] and *** for <0.001 and no stars plotted for non-significant values. Relates to Fig. 5.
