## Supplementary figures and images for "Identification of a type 1 diabetes-associated T cell receptor repertoire signature from the human peripheral blood"

### Extended_data_figure_1.png

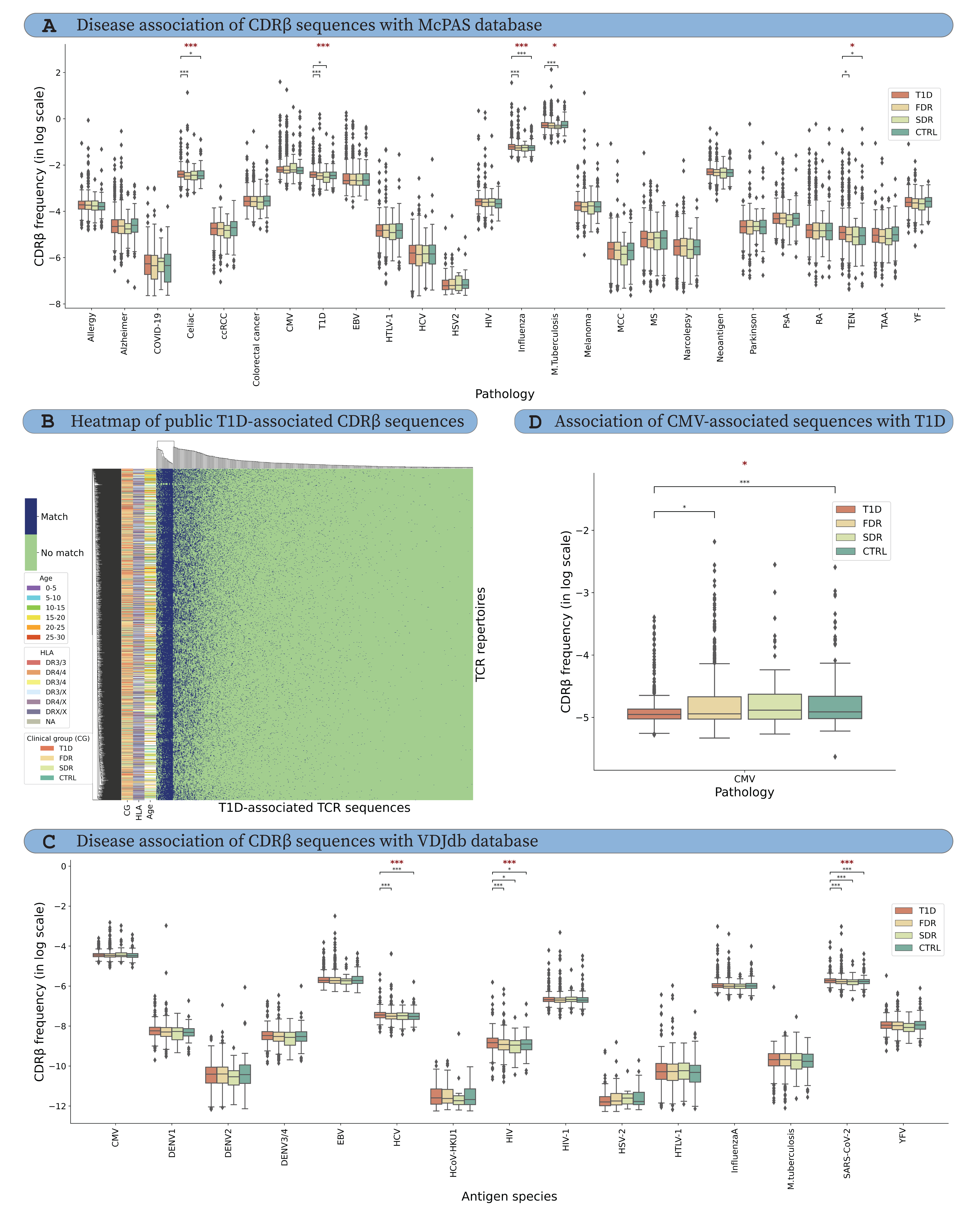

### Extended_data_figure_2.png

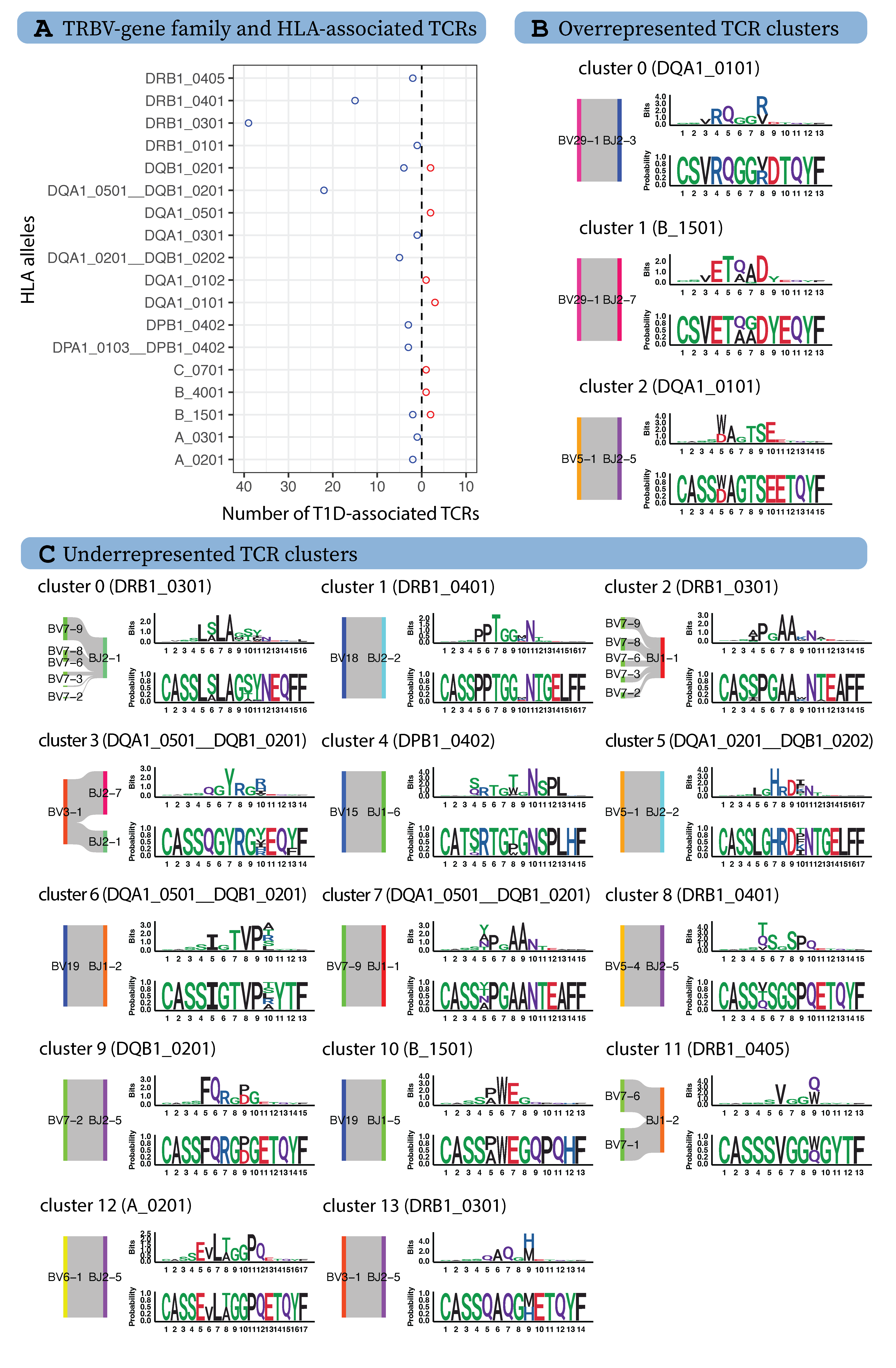

### Extended_data_figure_3.png

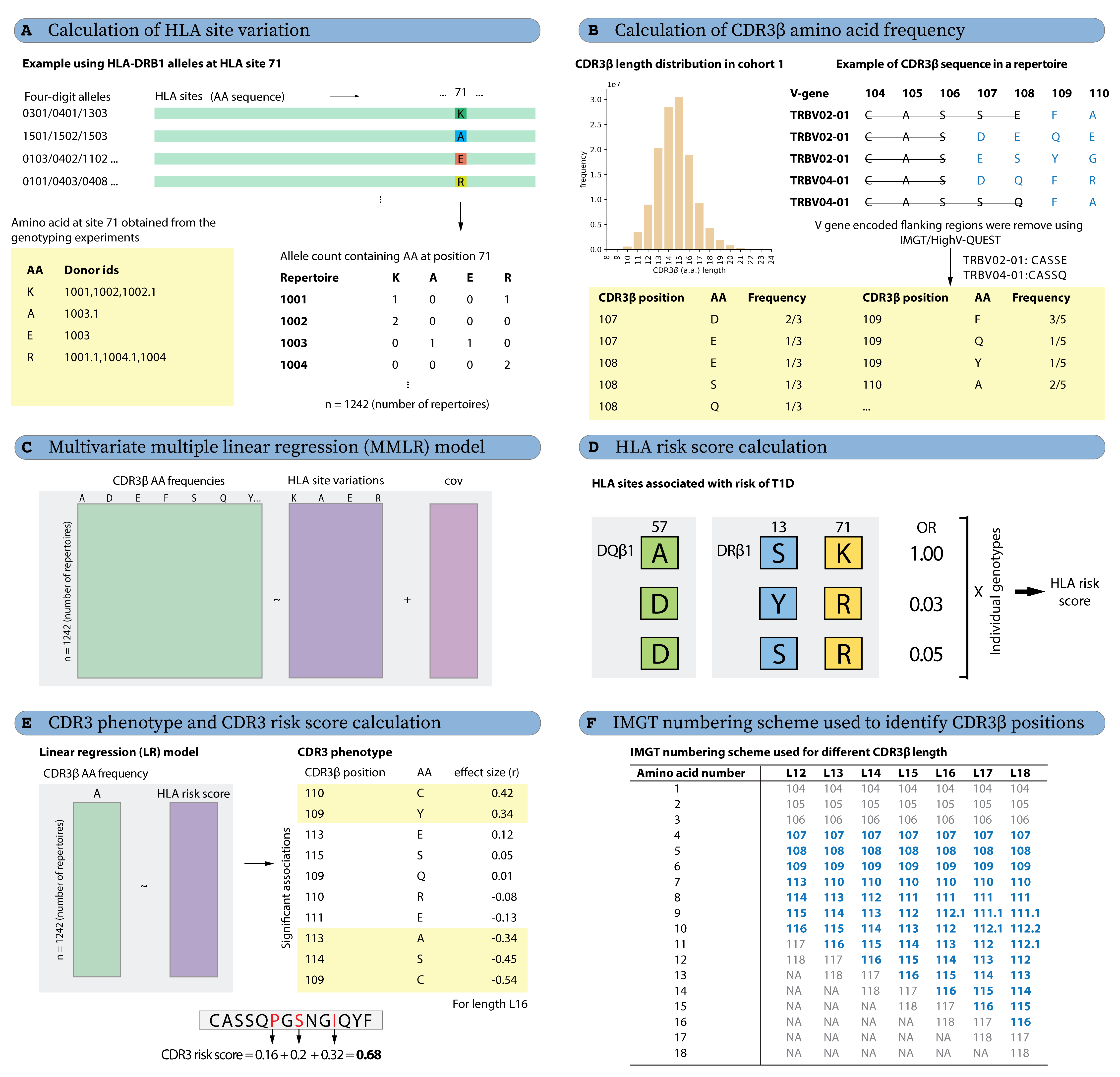

### Extended_data_figure_4.png

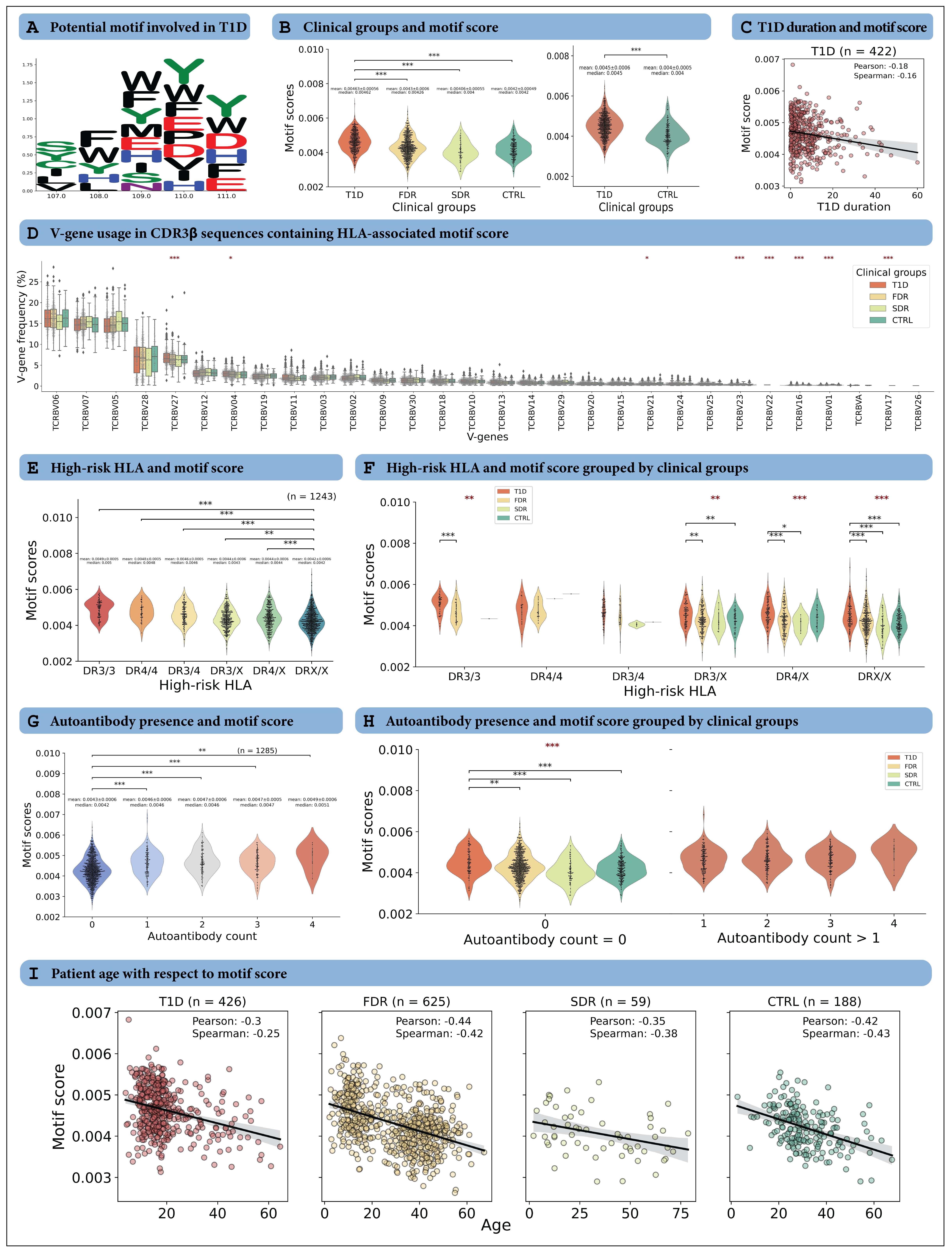

### Extended_data_figure_5.png

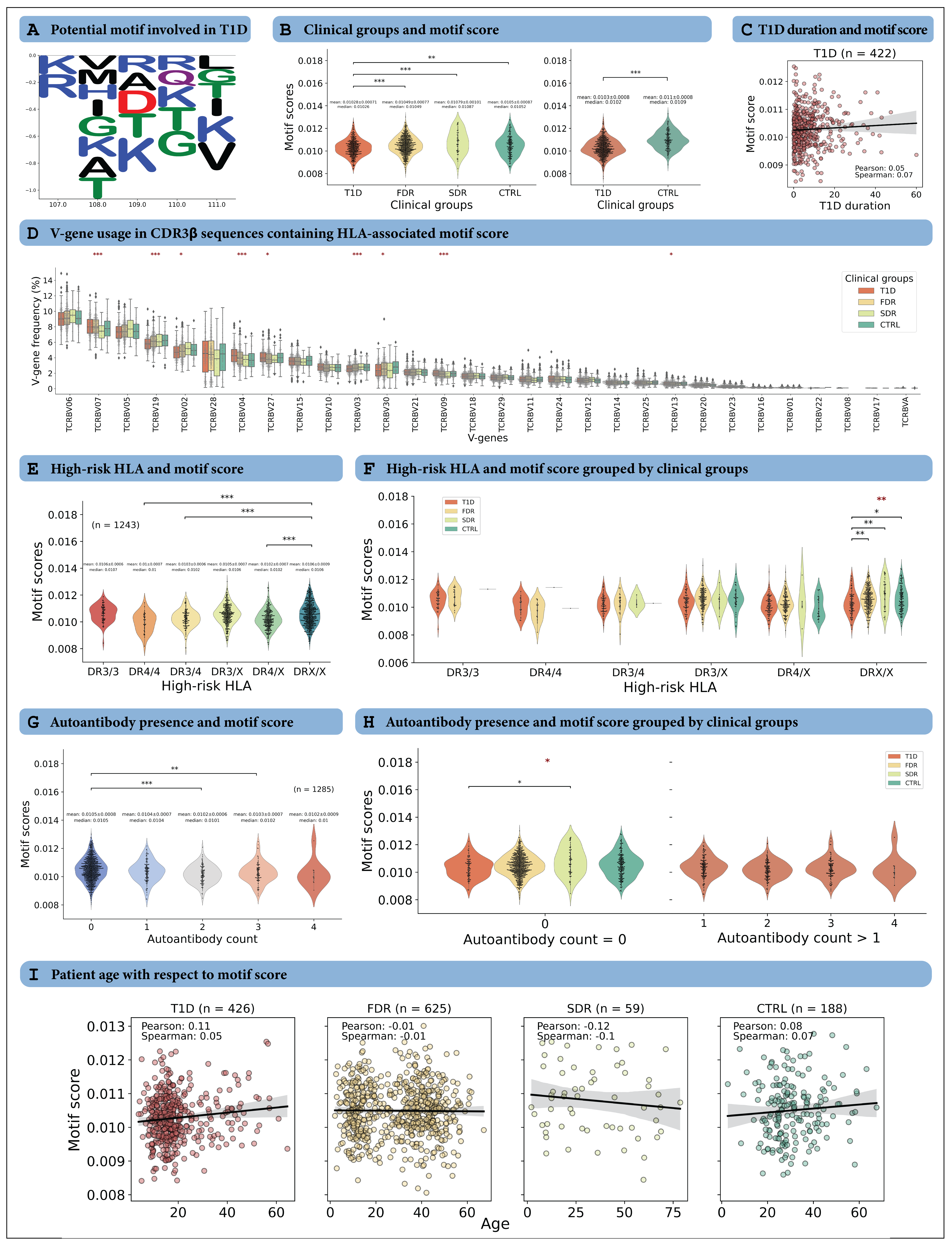

### Extended_data_figure_6.png

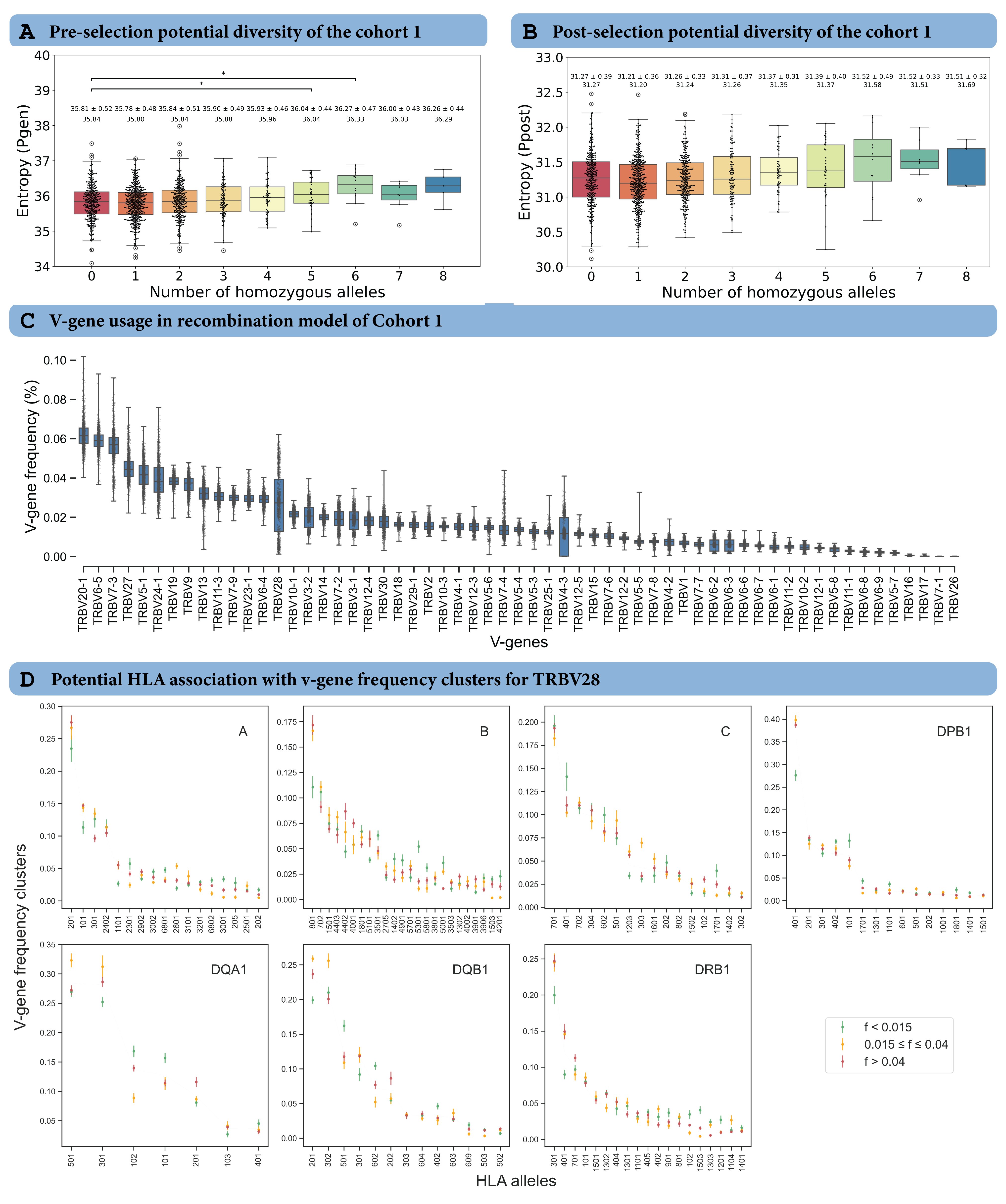

### Extended_data_figure_7.png

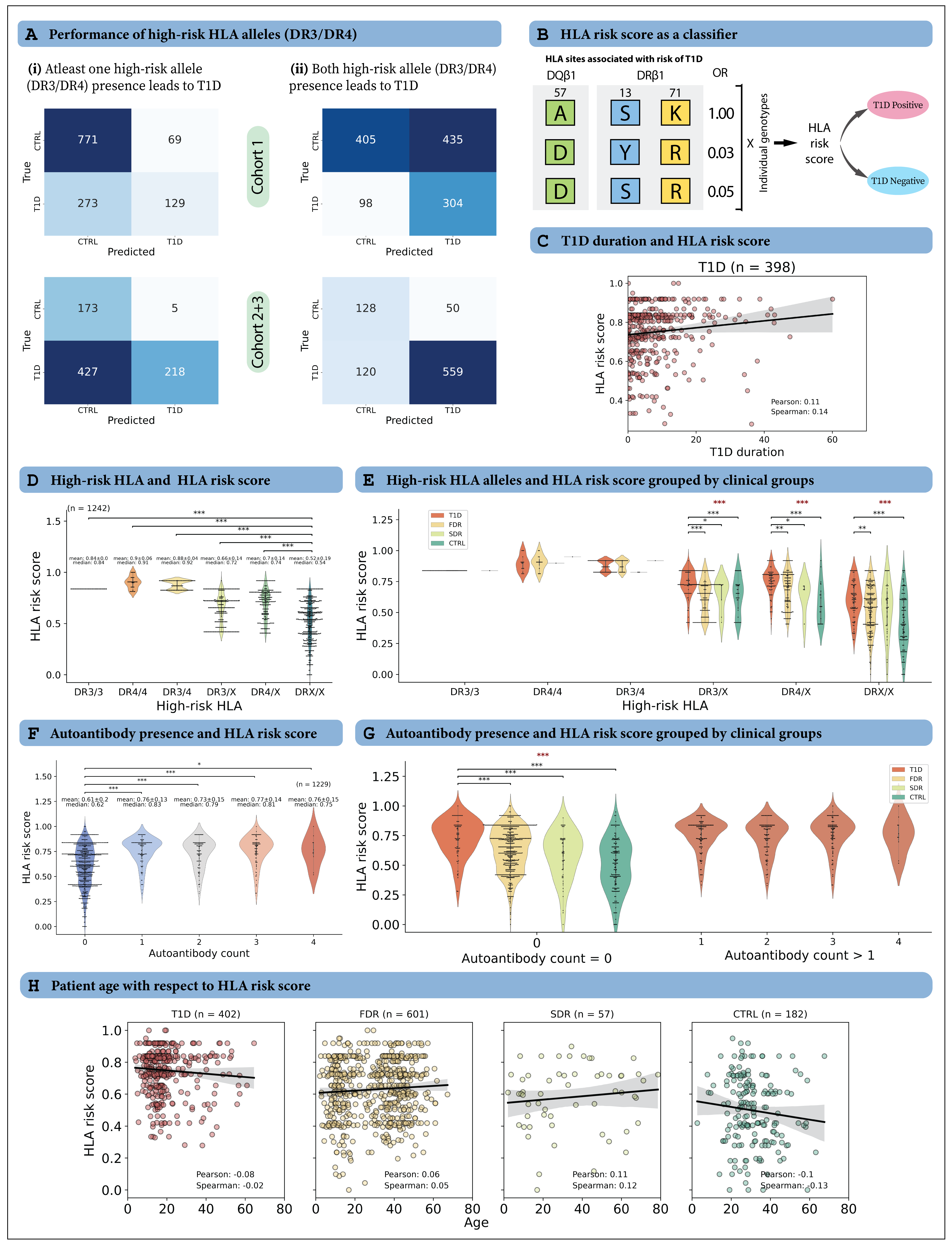

### Extended_data_figure_8.png

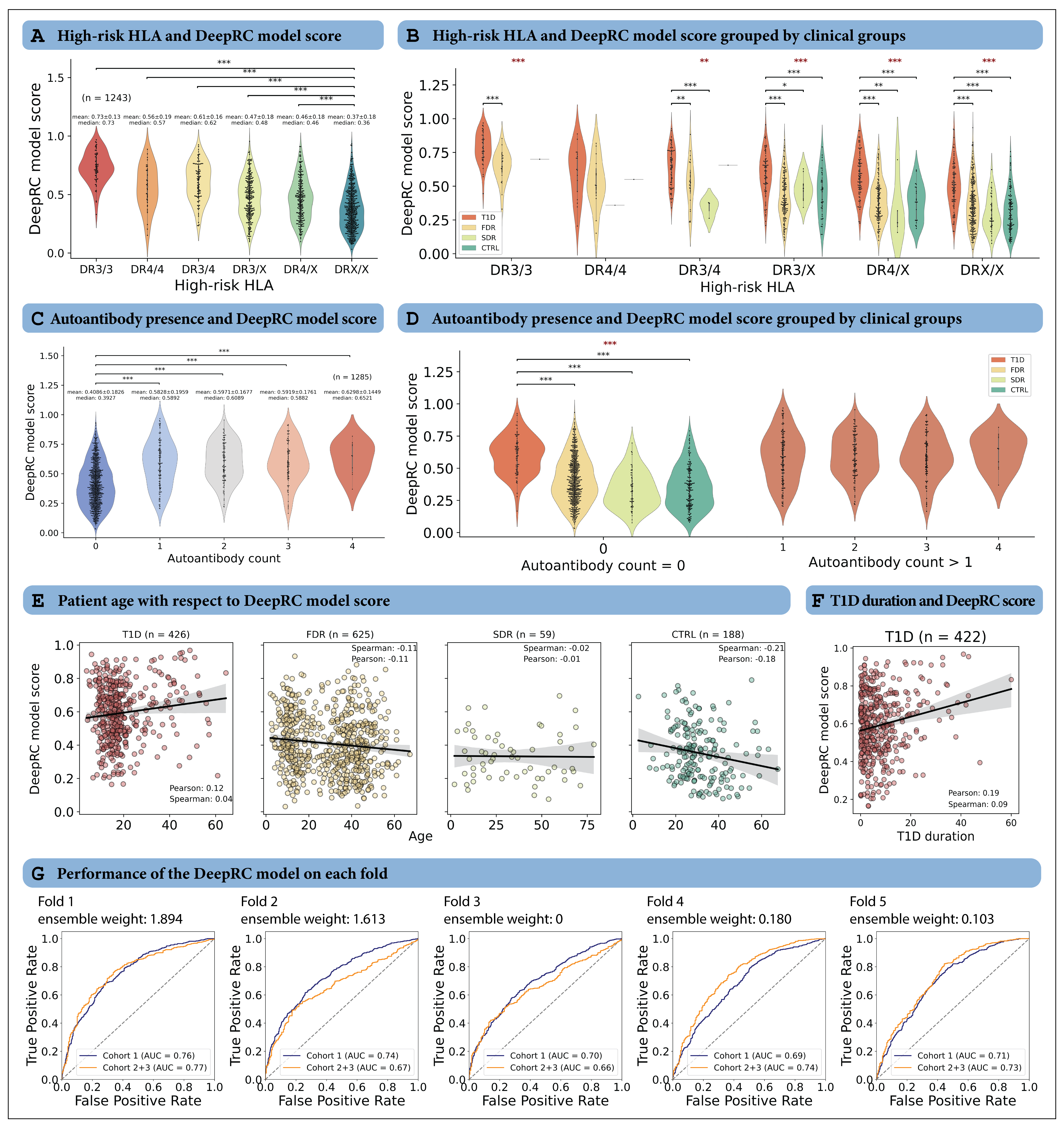

### Extended_data_figure_9.png

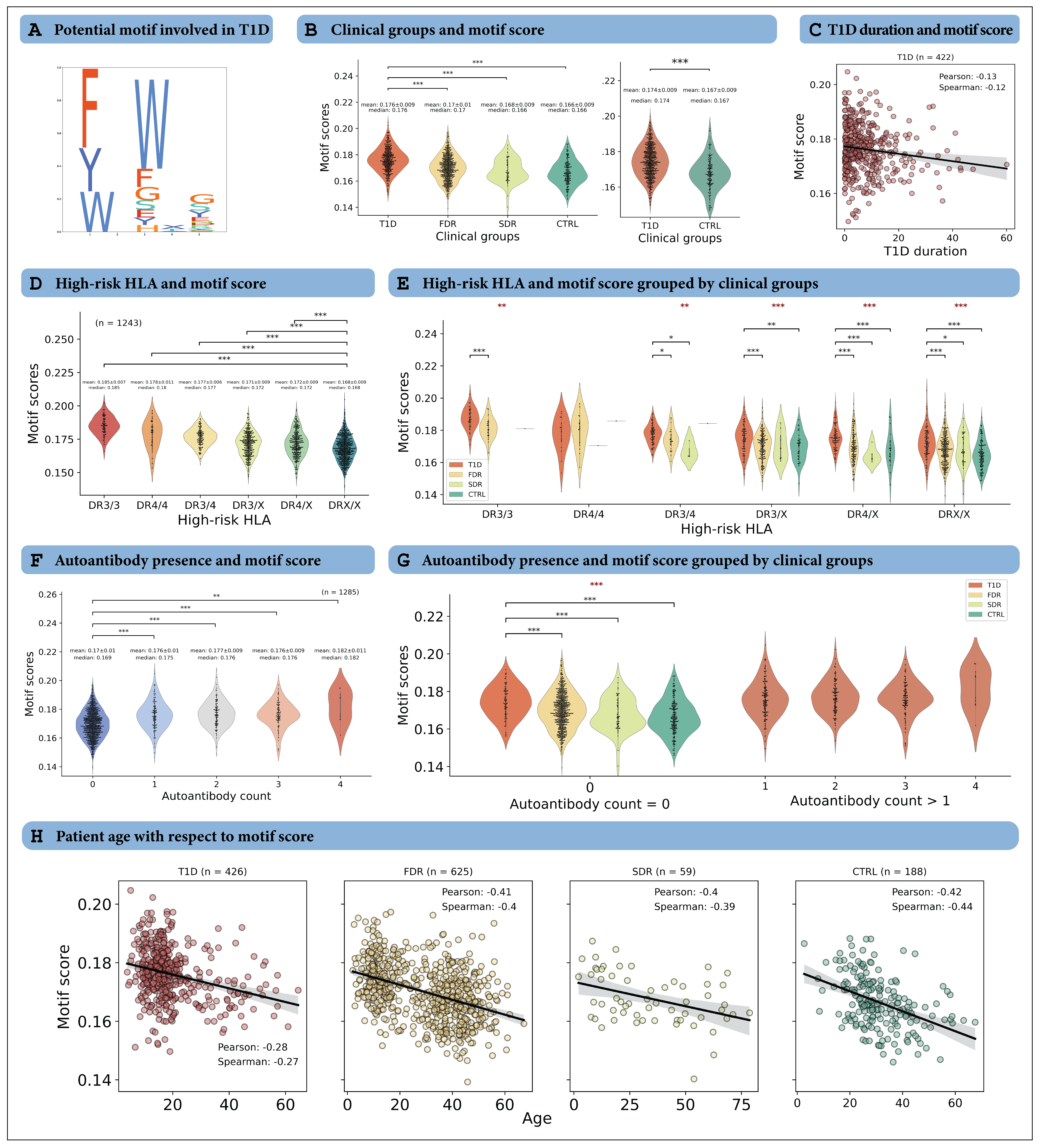

### Extended_data_figure_10.png

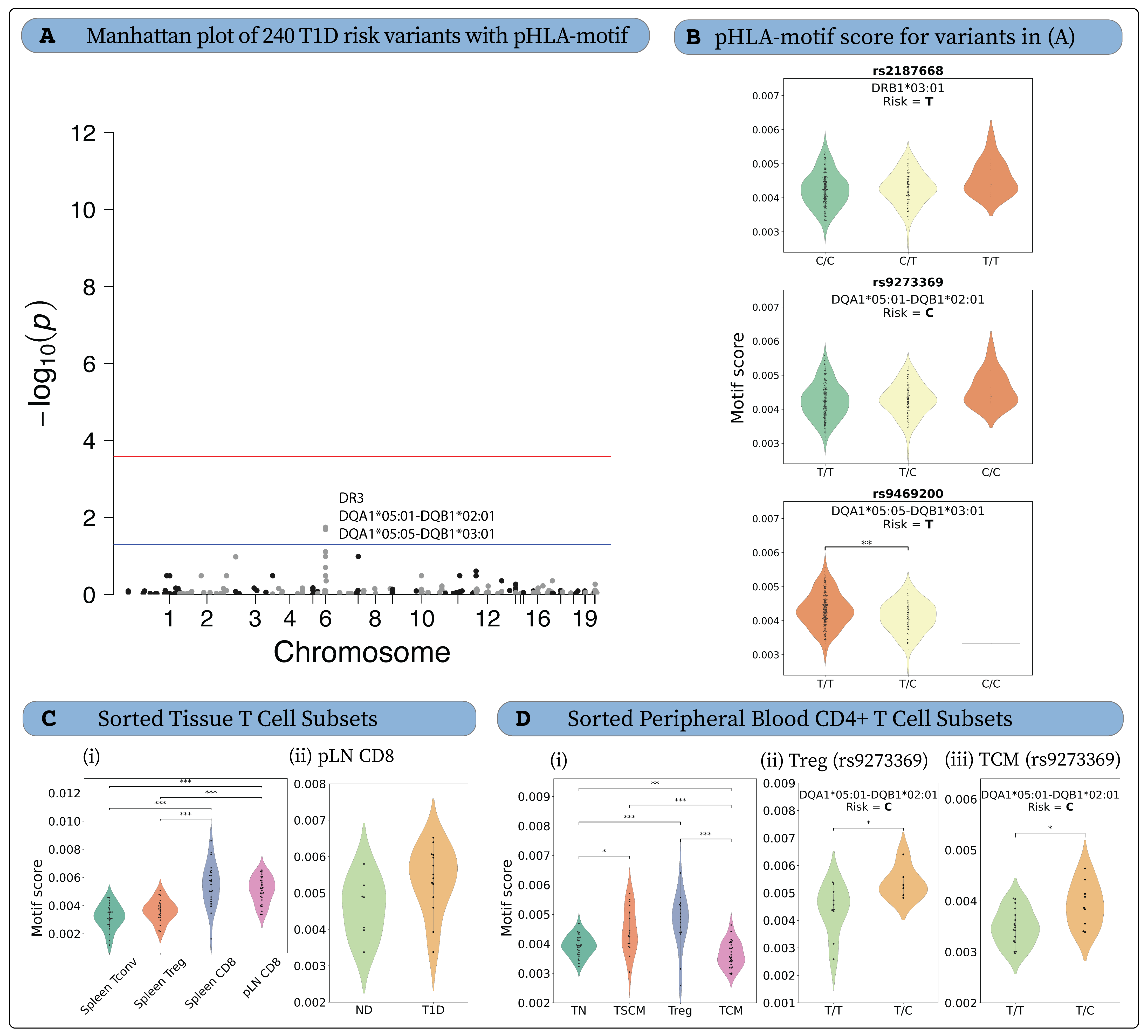
